# Somatic mutation and selection at epidemiological scale

**DOI:** 10.1101/2024.10.30.24316422

**Authors:** Andrew R. J. Lawson, Federico Abascal, Pantelis A. Nicola, Stefanie V. Lensing, Amy L. Roberts, Georgios Kalantzis, Adrian Baez-Ortega, Natalia Brzozowska, Julia S. El-Sayed Moustafa, Dovile Vaitkute, Belma Jakupovic, Ayrun Nessa, Samuel Wadge, Anna L. Paterson, Doris M. Rassl, Raul E. Alcantara, Laura O’Neill, Sara Widaa, Siobhan Austin-Guest, Matthew D. C. Neville, Moritz J. Przybilla, Wei Cheng, Maria Morra, Lucy Sykes, Matthew Mayho, Nicole Müller-Sienerth, Nick Williams, Diana Alexander, Luke M. R. Harvey, Thomas Clarke, Alex Byrne, Jamie R. Blundell, Matthew D. Young, Krishnaa T. A. Mahbubani, Kourosh Saeb-Parsy, Hilary C. Martin, Michael R. Stratton, Peter J. Campbell, Raheleh Rahbari, Kerrin S. Small, Iñigo Martincorena

## Abstract

As we age, many tissues become colonised by microscopic clones carrying somatic driver mutations (^1–10^. Some of these clones represent a first step towards cancer whereas others may contribute to ageing and other diseases. However, our understanding of the clonal landscapes of human tissues, and their impact on cancer risk, ageing and disease, remains limited due to the challenge of detecting somatic mutations present in small numbers of cells. Here, we introduce a new version of nanorate sequencing (NanoSeq)^11^, a duplex sequencing method with error rates <5 errors per billion base pairs, which is compatible with whole-exome and targeted gene sequencing. Deep sequencing of polyclonal samples with single-molecule sensitivity enables the simultaneous detection of mutations in large numbers of clones, yielding accurate somatic mutation rates, mutational signatures and driver mutation frequencies in any tissue. Applying targeted NanoSeq to 1,042 non-invasive samples of oral epithelium and 371 samples of blood from a twin cohort, we found an unprecedentedly rich landscape of selection, with 49 genes under positive selection driving clonal expansions in the oral epithelium, over 62,000 driver mutations, and evidence of negative selection in some genes. The high number of positively selected mutations in multiple genes provides high-resolution maps of selection across coding and non-coding sites, a form of in vivo saturation mutagenesis. Multivariate regression models enable mutational epidemiology studies on how carcinogenic exposures and cancer risk factors, such as age, tobacco or alcohol, alter the acquisition and selection of somatic mutations. Accurate single-molecule sequencing has the potential to unveil the polyclonal landscape of any tissue, providing a powerful tool to study early carcinogenesis, cancer prevention and the role of somatic mutations in ageing and disease.

## Introduction

In the past decade, studies using progressively more advanced sequencing techniques have begun to unravel the patterns of somatic mutation in human tissues. They have revealed that somatic mutations accumulate linearly with age in a tissue-specific manner ^3,12,13^, largely due to endogenous mutational processes but also influenced by mutagen exposures ^9^, germline variation ^14^ and disease states ^15,16^. Unexpectedly, these studies have also revealed that, as we age, our tissues become progressively colonised by myriad clones carrying positively selected driver mutations. By middle age, tissues such as blood, skin, oesophagus, endometrium, bladder, liver, and lung, are patchworks of mutant clones ^1–10^. These clones provide a window into the earliest stages of cancer development and may contribute to other diseases. However, most of these clones are microscopic and methods to detect them, such as laser microdissection ^17^ or single-cell in vitro expansion ^18–21^, are low throughput, which has limited our understanding of this phenomenon to a minority of tissues and small cohorts of individuals.

An alternative approach is error-corrected bulk sequencing ^22^. A particularly accurate strategy is duplex sequencing, which combines information from both strands of each original DNA molecule to eliminate sequencing errors and amplification errors ^23,24^. Theoretically, duplex sequencing error rates should approximate the polymerase error rate squared (10^−8^ to 10^−10^ errors per base pair [bp]) ^25^. However, error rates are typically higher (∼10^−7^) due to the copying of errors between strands by DNA polymerases used for end-repair or A-tailing during library preparation ^11^. We have previously described NanoSeq, a duplex sequencing protocol that avoids this issue by using restriction enzyme fragmentation without end repair, and dideoxy-nucleotides (ddBTPs) during A-tailing, achieving error rates <5×10^−9^ errors/bp in single DNA molecules ^11^. Since this rate is two orders of magnitude lower than the mutation burden of normal adult cells (∼10^−7^) ^18^, mutations are accurately detected in single molecules of DNA, enabling the quantification of mutation burdens and signatures in any tissue, including post-mitotic and polyclonal tissues. However, the original NanoSeq protocol could not be used for driver discovery, as the use of restriction enzymes resulted in partial coverage of the human genome.

### Accurate full-genome single-molecule sequencing

To develop a version of NanoSeq with full genome representation, we replaced restriction enzyme digestion by two alternative sequence-agnostic forms of blunt-ended genome fragmentation: (1) sonication followed by exonuclease digestion of overhangs, and (2) enzymatic fragmentation with a commercial enzyme mix in a reaction buffer optimized specifically for the NanoSeq workflow to eliminate any potential for the transfer of errors between strands. As in the original version of NanoSeq ^11^, ddBTPs are used to prevent the extension of single-stranded nicks and quantitative PCR followed by a library bottleneck is used to ensure optimal duplicate rates to maximise cost efficiency. Following optimisation, these protocols yielded full-genome duplex coverage with similar library yields and error rates to the original NanoSeq protocol (**Supplementary Note 1**).

To compare the error rates of these protocols to standard duplex sequencing, we used cord blood DNA. Neonatal blood has just ∼60-80 somatic mutations per cell (∼10^−8^ mutations/bp), providing a useful negative control. Both new versions of NanoSeq (sonication and enzymatic) yielded mutation loads and mutational spectra consistent with prior knowledge from single-cell derived colonies and restriction-enzyme NanoSeq (**Fig. 1a,b**) ^11^. In contrast, standard duplex sequencing (with end repair and without ddBTPs), using sonication or enzymatic fragmentation, yielded error rates around 1.5×10^−7^ and 4×10^−8^ errors/bp, respectively. Since many errors in standard duplex sequencing are caused by fixing DNA damage, error rates can be higher in highly damaged DNA. To study this, we applied these protocols to two adult pancreas biopsies fixed in formalin for 3 and 17 days prior to paraffin embedding. Error rates in standard duplex sequencing increased ∼10-fold in these samples, whereas the mutation loads from both versions of NanoSeq were comparable to those from a control biopsy (snap-frozen not formalin-fixed) (**Fig. 1c**). This raises the possibility of using NanoSeq on more heavily damaged sources of DNA. The changes introduced here to prevent the copying of errors between strands could also be applied to other error-corrected sequencing methods ^26,27^.

**Figure 1.**
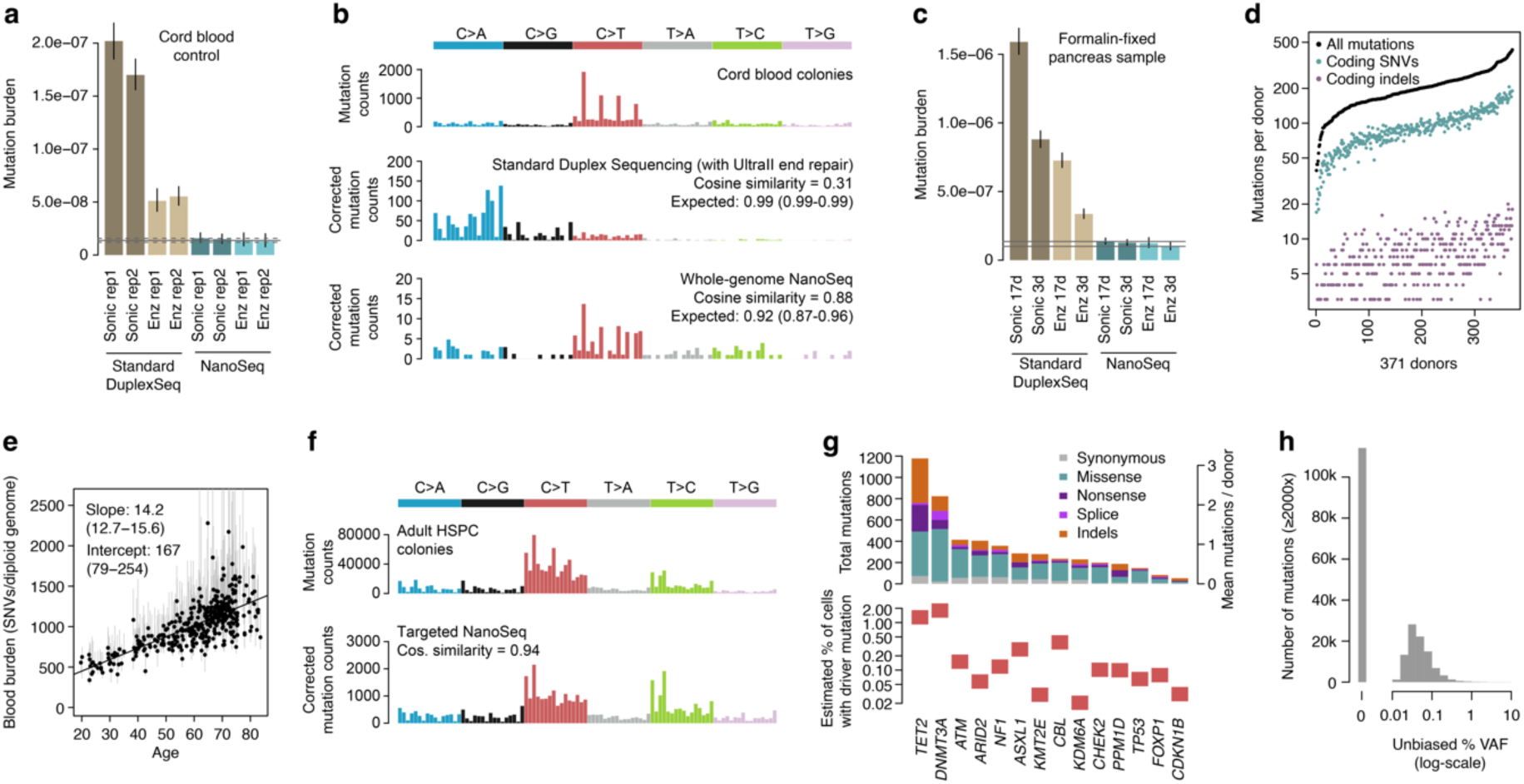
Technical and biological validation of Targeted NanoSeq. **a**, Genome-wide SNV burden estimates for cord blood granulocytes, sequenced using four different fragmentation and library preparation protocols (*sonic* refers to sonication and *enz* to enzymatic fragmentation). Error bars show Poisson 95% confidence intervals (CIs). Horizontal lines denote the observed burden (solid) and 95% Poisson CIs (dashed) for cord blood granulocytes sequenced by restriction-enzyme NanoSeq^11^. Duplex Sequencing and NanoSeq burdens are corrected for missed embryonic mutations, as described in ^11^. **b**, Trinucleotide mutational spectra of single-cell derived cord blood colonies from a previous study^65^ (top), and cord blood granulocytes sequenced using standard duplex sequencing (middle) and whole-genome NanoSeq (bottom). Duplex sequencing and NanoSeq spectra are corrected by the ratio of genomic to observed trinucleotide frequencies. Cosine similarity 95% CIs are calculated by drawing 1,000 random samples from each observed profile, as described in^11^. **c**, Genome-wide mutation burden estimates for adult pancreas samples formalin-fixed for 3 days (3d) or 17 days (17d), and sequenced using four different protocols. Horizontal lines denote the burden and the associated Poisson 95% CIs in a matching fresh-frozen sample. **d**, Numbers of total mutations, coding SNVs and coding indels identified in whole blood samples from 371 donors using targeted NanoSeq. **e**, Linear regression of genome-wide SNV burdens (estimated using targeted NanoSeq) for whole blood samples from 371 donors against donor age. Points and their associated error bars represent the point estimates and 95% Poisson bootstrapping CIs of passenger mutation burdens for each sample. Slope and intercept of the fitted model (point estimates and 95% CIs) are indicated. One sample was excluded due to the ratio between upper and lower confidence limits being >5. **f**, Trinucleotide mutational spectra of adult haematopoietic stem and progenitor cell (HSPC) colonies from a previous study ^65^ and whole blood samples sequenced using targeted NanoSeq (corrected by the ratio of genomic to observed trinucleotide frequencies). **g**, Mutation counts for each coding mutation consequence (top) and estimated mutant cell fractions (bottom) for 14 genes under significant positive selection in blood. Mutant cell fractions are shown for individuals aged 65-85 whose blood samples were not selected on the basis of their oral epithelium results. **h**, Distribution of (log_10_-scaled) unbiased VAFs for mutations with sequencing depth ≥2000× identified in 371 whole blood samples using targeted NanoSeq. Unbiased VAFs were calculated from read bundles not used for duplex variant calling.

### Somatic mutation and selection in polyclonal tissues

Combining these new protocols with bait capture (a common step in duplex sequencing, ^28^), targeted NanoSeq can be used to accurately quantify somatic mutation rates, mutational signatures and driver landscapes in any tissue, regardless of clonality. Unlike traditional bulk sequencing, which only detects mutations over a certain variant allele fraction (VAF) (typically >1%), accurate single-molecule sequencing can potentially detect mutations present at any cell fraction, even in single cells, with a detection probability proportional to the frequency of the mutation in the cell population. In highly polyclonal samples, where the number of clones is larger than the duplex depth, most mutations are seen in just one molecule, providing an efficient way to simultaneously study driver mutations in hundreds or thousands of clones with a single DNA sequencing library.

Our first studies of somatic mutation in normal skin and oesophagus revealed a rich landscape of mutation and selection but were limited to fewer than 10 individuals ^2,3^. To investigate how mutation landscapes vary across the population in a similar squamous tissue we chose oral epithelium, which can be collected non-invasively using buccal swabs. Here we describe the mutation landscape of oral epithelium across 1,042 individuals, by applying targeted NanoSeq to buccal swabs with a panel of 239 cancer genes (bait footprint 0.9 Mb) (**Methods**, **Extended Data Fig. 1,2**, **Extended Data Table 1**). Buccal swabs were sequenced to an average depth of 665 duplex coverage (dx) per sample (693,208 dx cumulative coverage across all samples, requiring around 21,000× raw coverage per sample). From 371 donors, we also applied targeted NanoSeq to archival blood samples (cumulative 250,947 dx).

### Single-molecule mutation landscape of blood

Analysis of the blood data further validates the technology. Targeted NanoSeq confirmed the mutation rate and the trinucleotide spectrum of blood known from previous whole-genome NanoSeq and whole-genome sequencing of haematopoietic stem cell colonies (**Fig. 1d-f**). To detect genes under positive selection in blood we then applied dNdScv ^29^, an implementation of dN/dS (the ratio of nonsynonymous to synonymous mutation rates) that corrects for sequence composition, trinucleotide mutation rates, and variable rates across genes. This identified 14 genes under significant positive selection (**Fig. 1g, Extended Data Fig. 3**, **Extended Data Table 3**), all known clonal haematopoiesis drivers ^30,31^. Hotspot dN/dS analyses also identified evidence of selection on several additional drivers, including *JAK2*, *MYD88*, *SF3B1*, *SRSF2, GNB1* and *STAT3* (**Extended Data Table 4**, **Supplementary Note 2**).

Despite the modest size of the dataset (371 samples, 676 mean dx), we found 4,406 non-synonymous mutations in these driver genes (11.9 per donor on average), including 800 mutations in *DNMT3A* and 1,104 in *TET2*. 95% of mutations detected were called by just one molecule, 99% had unbiased VAFs under 1%, and 90% under 0.1% (**Fig. 1h**, **Methods)**. In contrast, a recent study of clonal haematopoiesis in over 200,000 individuals using standard sequencing (only sensitive to clones with >1% VAF) found 0.029 and 0.012 *DNMT3A* and *TET2* mutations per donor, an approximately 100-200-fold lower yield of driver mutations per sample. Overall, these results exemplify the power of accurate single-molecule sequencing to measure somatic mutation rates, mutational spectra and clonal selection landscapes of highly polyclonal samples.

### A rich driver landscape in oral epithelium

Unlike in blood, knowledge of somatic mutation and selection in solid tissues has been limited to small cohorts. This is due to the microscopic size of most clones, requiring non-standard sequencing strategies, and the difficulty of collecting large numbers of invasive samples of healthy tissue compatible with these technologies. Accurate single-molecule sequencing of non-invasive samples provides a means of overcoming these limitations to study somatic mutation and selection at population scale.

Buccal swabs were collected by post from 1,042 volunteers in the TwinsUK registry ^32^: median age 68 (range 21-91), 79% female, 37% smokers, including 332 pairs of twins (214 identical/monozygotic, 118 non-identical/dizygotic) (**Extended Data Fig. 1**). A protocol designed to reduce saliva and blood contamination was used, with methylation and mutation analyses confirming a mean epithelial fraction >90% (**Extended Data Fig. 2**, **Methods**). Across donors, we found 341,682 mutations, including 160,708 coding single-nucleotide variants (SNVs) and 29,333 coding indels (**Fig. 2a**). Calculation of mutation burdens per sample revealed that mutations in oral epithelium accumulate linearly with age, with estimated rates of ∼18.0 SNVs/cell/year (95% confidence interval [CI_95%_]: 16.7-19.4) and ∼2.0 indels/cell/year (CI_95%_: 1.7-2.4) (**Fig. 2b,c**). Since these rates are extrapolated from genic sequences, which often have lower mutation rates than intergenic sequences, we performed whole-genome NanoSeq on 16 samples to study the distribution of mutations across the genome. This revealed a genome-wide rate for oral epithelium of ∼23 SNVs/cell/year (**Extended Data Fig. 4**, **Supplementary Note 3**).

**Figure 2.**
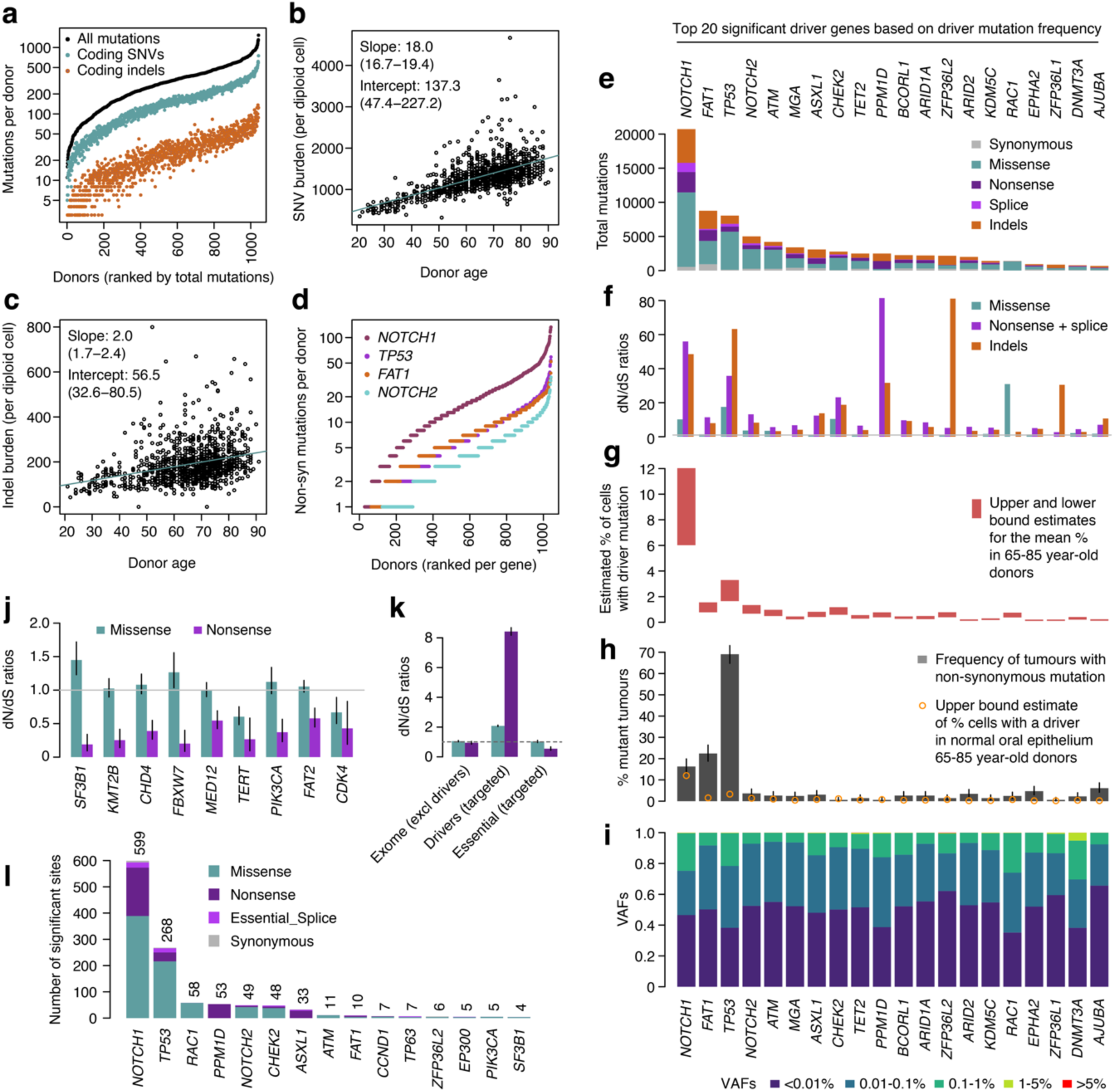
Driver landscape of oral epithelium in 1,042 donors. **a**, Numbers of total mutations, coding SNVs and coding indels identified in oral epithelium samples from 1,042 donors using targeted NanoSeq. **b, c**, Linear regressions of genome-wide (**b**) SNV and (**c**) indel burdens in oral epithelium (estimated using targeted NanoSeq) against donor age. Points and their associated error bars represent the point estimates and 95% Poisson bootstrapping CIs of passenger mutation burdens for each sample. Slope and intercept of the fitted model (point estimates and 95% CIs) are indicated. **d**, Numbers of non-synonymous mutations per donor in oral epithelium for genes *NOTCH1*, *TP53*, *FAT1* and *NOTCH2*. Mutation counts are ordered independently for each gene. **e-i**, For the top 20 significant driver genes based on driver mutation frequency, panels show (**e**) mutation counts per mutation consequence category, (**f**) dN/dS ratios per mutation consequence category (horizontal line indicates neutral dN/dS=1, only categories with significant dN/dS ratios are shown for each gene), (**g**) estimated mutant cell percentages (upper and lower bounds for the mean across donors aged 65-85), (**h**) percentage of tumours carrying a non-synonymous mutation (with error bars denoting 95% binomial confidence intervals), and (**i**) distribution of unbiased mutation VAFs. **j**, dN/dS ratios for missense and nonsense mutations in genes under significant negative selection. Error bars denote 95% CIs; horizontal line indicates neutral dN/dS=1. **k**, Global dN/dS ratios for missense and nonsense mutations across non-driver genes, targeted driver genes, and 17 targeted essential genes. Error bars and horizontal line denote the same quantities as in **j**. **l**, Numbers of amino acid changes under significant positive selection based on site-level dN/dS (site-wide or under restricted hypothesis testing of known cancer hotspots), grouped by gene and mutation consequence category. Counts of significant amino acid changes per gene are shown above each bar.

By sequencing large numbers of clones simultaneously, these data captured an unprecedentedly rich landscape of selection. We found 49 genes under significant positive selection by dNdScv, with a total of 90,000 non-synonymous mutations, of which ∼62,000 are estimated to be driver mutations (**Fig. 2d-i**, **Extended Data Fig. 5**, **Extended Data Table 3**, **Supplementary Note 2)**. Of these 49 genes, at least 21 are significantly mutated in head and neck cancers from TCGA (The Cancer Genome Atlas) (**Supplementary Note 2**). The highest numbers of non-synonymous mutations per gene were found in *NOTCH1* (n=20,185), *TP53* (n=7,942), *FAT1* (n=7,826), and *NOTCH2* (n=4,733), with *NOTCH1*, *TP53*, *CHEK2*, *PPM1D* and *RAC1* showing the highest dN/dS ratios (**Fig. 2e,f**). Reassuringly, the commonest drivers in oral epithelium matched those reported in skin and oesophagus ^2–4^, consistent with these tissues being similar epithelia composed mostly of keratinocytes.

Duplex sequencing is normally applied to small gene panels ^23,28^, but our use of a library quantification step followed by a bottleneck on the library complexity, simplifies the use of panels of any size as well as whole-exome capture or whole-genome sequencing. To showcase this and to ensure that major drivers are not being missed by our 239-gene panel, we performed exome-wide NanoSeq on 12 samples to a total duplex coverage of 1,024 dx. This analysis reidentified *NOTCH1*, *TP53*, *PPM1D*, *RAC1* and *ZFP36L2*, suggesting that our targeted gene panel already includes the commonest driver genes in oral epithelium. We also found a significant excess of indels in the keratin gene *KRT15*, which rather than selection is likely the result of a hypermutation process known to affect highly expressed lineage-defining genes ^33,34^.

Of all mutations in the buccal data, 95.5% were found in only one duplex molecule, and ∼90% had unbiased VAFs under 0.1%. These VAFs seem consistent with the microscopic size of most clones reported in skin and oesophagus ^2,3,7^, and emphasise the need of single-molecule sensitivity to study solid tissues in bulk. Analysis of matched blood samples confirmed that the signal of selection on *DNMT3A*, *TET2* and *FOXP1* (**Fig. 2e**) derives from contaminating blood cells, whereas the signals in other genes are dominated by oral epithelium (**Extended Data Fig. 3c**, **Methods**). This includes *PPM1D* and *ASXL1*, which are drivers in both oral epithelium and blood.

Aggregating duplex VAFs (**Methods**), we estimate that, in donors aged 65-85, the average fraction of cells carrying a driver mutation is ∼10% for *NOTCH1*, ∼3% for *TP53*, ∼1% for *NOTCH2*, *CHEK2* and *ATM*, and <1% for other driver genes (**Fig. 2g**). Whereas *NOTCH1* dominates the driver landscape in oral epithelium, we note that its frequency is several fold lower than in normal oesophagus ^3,35^. The frequency of mutations in *NOTCH1* and *TP53* in normal epithelium also contrasts with their frequencies in TCGA head and neck squamous carcinomas of 16% and 69%, respectively (**Fig. 2h**, **Methods**). The similar frequency of *NOTCH1* driver mutations in oral cancer and normal oral epithelium suggests that *NOTCH1* mutations lead to benign clonal expansions at no greater risk of transformation than *NOTCH1*-wildtype cells. In contrast, *TP53* and most other driver genes found under selection in oral epithelium appear enriched in squamous carcinomas consistent with a genuine tumorigenic role of these mutations. We note, however, that detailed comparisons are limited by the number of cancers sequenced to date, compared to the thousands of normal clones that can be assayed simultaneously in a small number of individuals.

### Negative selection on essential genes

The high number of mutations detected per gene also provides unprecedented power to detect negative selection (i.e. genes with a depletion of non-synonymous mutations: dN/dS<1). Previous studies have shown that negative selection is much weaker in somatic evolution than in germline evolution, with only a small minority of coding mutations exome-wide being lost through negative selection in somatic tissues and cancers ^29,36^. However, previous studies could not rule out the possibility of strong negative selection in a small minority of genes, particularly in essential haploinsufficient genes.

Using new one-sided negative selection tests in dNdScv, we found 9 genes under significant negative selection in our targeted panel, mostly driven by selection against truncating SNVs (dN/dS<1 for nonsense and essential splice site mutations) (**Fig. 2j**, **Extended Data Table 3**). This includes three essential genes from CRISPR screens (*SF3B1*, *CHD4* and *CDK4*) (**Methods**). *PIK3CA*, *SF3B1* and *TERT* showed interesting patterns of negative selection against truncating mutations and positive selection on activating hotspot mutations (coding in *PIK3CA* and *SF3B1*, and promoter in *TERT*), suggesting that these genes are both essential genes in wild-type oral epithelial cells, and drivers upon activating mutations.

Aggregating mutations across gene sets, the global dN/dS ratios across the 49 driver genes were high (dN/dS ∼2 for missense, ∼8 for truncating), whereas global dN/dS ratios on truncating mutations across 17 genes essential in CRISPR screens and targeted in this study showed clear negative selection (dN/dS = 0.69, CI_95%_ 0.61-0.78) (**Fig. 2k**). In contrast, exome-wide dN/dS ratios, excluding the genes above, were consistent with a largely neutral accumulation of missense and truncating somatic mutations in most genes.

### In vivo saturation mutagenesis

To formalise the analysis of recurrent mutation hotspots, we used site-level dN/dS models (*sitednds*, **Methods**). Thanks to the high number of driver mutations, we found 1,220 amino acid changes with evidence of significant positive selection (q-value<0.01), including 599 in *NOTCH1* and 268 in *TP53* (**Fig. 2l**, **Methods**). Restricting hypothesis testing to known cancer hotspots added several oncogenes to the list of positively selected genes in buccal swabs, including *PIK3CA*, *ERBB2*, *KRAS*, and *HRAS* (**Extended Data Table 4**, **Methods**).

The high number of mutations per gene provides an opportunity to start building high-resolution maps of selection across sites for major driver genes (**Fig. 3a-c**), a form of in vivo saturation mutagenesis. The distribution of coding mutations across sites in *TP53* mirrors the distribution observed by aggregating thousands of cancer exomes and genomes from the COSMIC (Catalogue of Somatic Mutations in Cancer) database ^37^ (**Fig. 3a**, **Methods**). In *TP53*, we found nearly as many mutations as in 44,000 cancer exomes and genomes from COSMIC, and for several other driver genes, the number of mutations reported here far outweighs all previously observed mutations from cancer studies (**Fig. 3b**).

**Figure 3.**
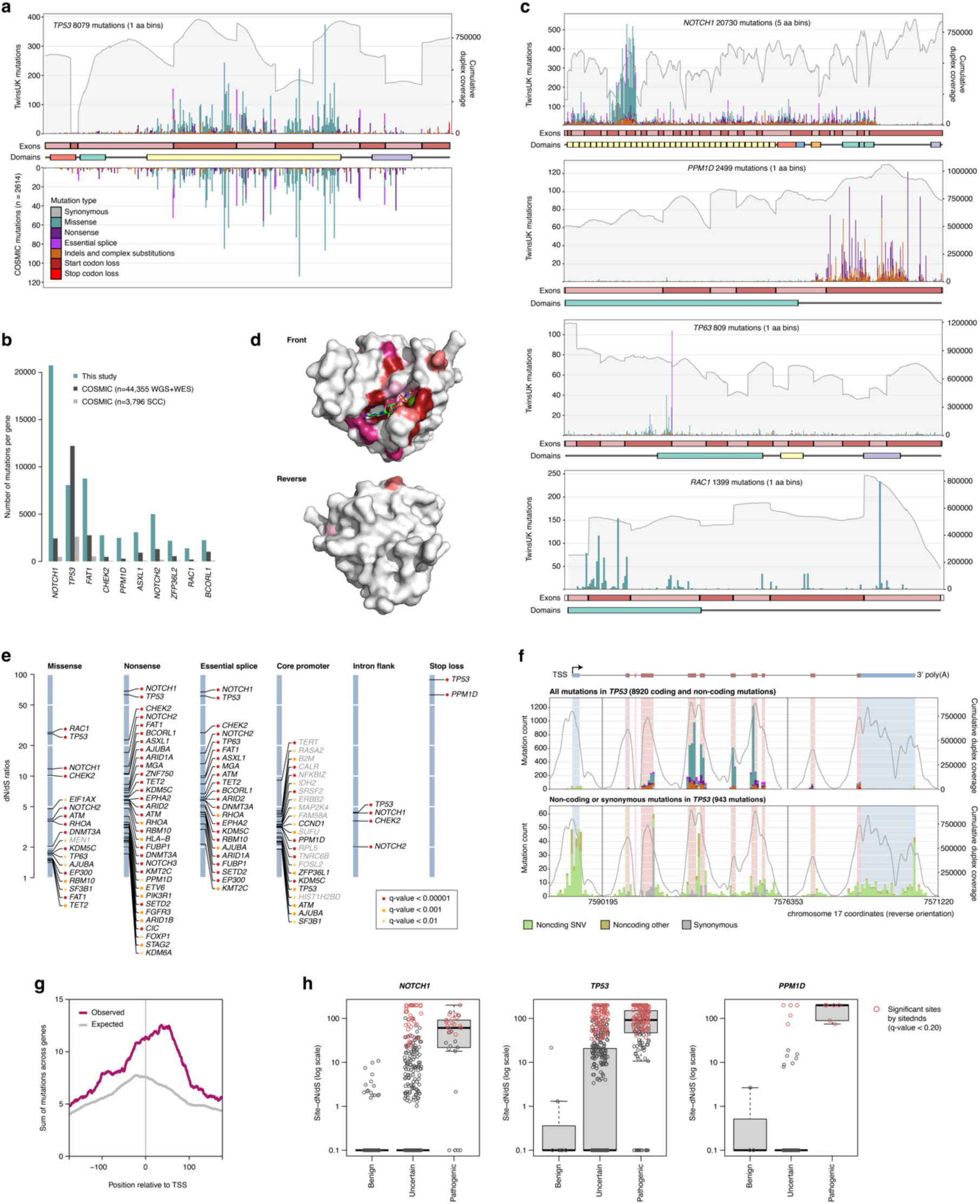
In vivo saturation mutagenesis in oral epithelium. **a**, Mutation histogram for *TP53*. The *x*-axis represents coordinates along the coding sequence. Exons and protein domains are indicated along the axis. The *y*-axis represents number of mutations, either in the 1,042 TwinsUK oral epithelium samples used in this study (top) or in squamous cell carcinoma from the COSMIC database. Mutations are coloured according to mutation consequence category. Grey shading indicates cumulative duplex coverage across TwinsUK buccal swab samples. **b**, Numbers of mutations per gene found in this study and in the COSMIC catalogue (obtained from across all whole-genome sequencing [WGS] and whole-exome sequencing [WES] studies or only squamous cell carcinoma [SCC] WGS and WES studies), for a selection of driver genes. **c**, Mutation histograms for *NOTCH1*, *PPM1D*, *TP63* and *RAC1*. Elements are as indicated in **a**; COSMIC mutations not shown. **d**, Diagrams of the 3-dimensional structure of Rac1, showing the clustering of sites under significant positive selection around the GDP/GTP binding pocket. Residues with site-level dN/dS *q*-value < 0.01 are coloured. Shading intensity denotes degree of significance. **e**, dN/dS ratios for driver sites under significant positive selection based on the *withingenednds* method. Driver sites are classified into six groups according to mutation consequence. Labels in grey indicate genes not identified as significant by gene-level dN/dS analyses. **f**, Mutation histograms for *TP53*, including all mutations (top) and synonymous or non-coding mutations only (bottom). The *x*-axis represents genomic coordinates along the gene body, with coding exons (red) and UTRs (blue) indicated by the gene diagram on top and the shading within each histogram. The grey line denotes cumulative duplex coverage across TwinsUK buccal swab samples. Coding mutation counts are coloured according to mutation consequence as indicated in **a**. **g**, Observed and expected (*withingenednds*) density of mutations as a function of position relative to the transcription start site (TSS), aggregated across all targeted genes. **h**, Distribution of site-level dN/dS ratios for sites annotated in ClinVar as benign, pathogenic or of uncertain significance, in *NOTCH1*, *TP53* and *PPM1D*. Significant sites are shown in red.

The distribution of mutations along each gene revealed a diversity of selection patterns (**Fig. 3a,c**) (see **Supplementary Note 4** for an extended description). *TP53* shows strong selection on missense mutations in the DNA binding domain and on truncating mutations throughout the gene, which mirrors the pattern from thousands of cancer genomes. *NOTCH1* shows a characteristic clustering of missense mutations in EGF repeats 8-12, predicted to disrupt binding to Notch1 ligands Jagged and Delta ^3^. Interestingly, truncating mutations are subject to much weaker selection in the last exon of *NOTCH1* (dN/dS for nonsense mutations 68.4 across the gene, 6.9 in the last exon), which likely reflects their inability to trigger nonsense-mediated decay. *RAC1* shows a classical oncogene pattern, with strong selection on activating missense hotspots, and site-dN/dS identifying 58 missense sites under significant selection (q-value<0.01). Although these sites appear scattered along the protein primary structure, they cluster around the GDP/GTP binding pocket in the 3-dimensional structure of Rac1 (**Fig. 3d**). *PPM1D* encodes a known negative regulator of p53 and shows a characteristic pattern of recurrent nonsense SNVs and indels in the last exon, which results in the loss of a C-terminal degradation domain leading to a functional more stable isoform of the protein, hence increasing p53 suppression ^38^. Finally, *TP63* shows an unusual selection pattern with a highly recurrent essential splice hotspot predicted to lead to an alternative isoform of p63 ^39^. Additional mutation maps are shown in **Extended Data Fig. 6**.

Beyond coding mutations, we obtained high duplex coverage in exon-flanking intronic sequences, and we targeted the promoters of multiple genes (**Methods**). To test for selection on specific subsets of coding and non-coding sites within a gene, we implemented a new function in dNdScv (*withingenednds*, **Supplementary Note 2**). This revealed multiple underappreciated driver sites, including strong positive selection on mutations causing the loss of the stop codon in *TP53* and *PPM1D*, on intronic mutations near essential splice sites in *TP53*, *NOTCH1*, *CHEK2*, and *NOTCH2*, and on some synonymous sites in *TP53* and *NOTCH1* predicted to impact splicing by SpliceAI ^40^ (**Fig. 3e**, **Supplementary Note 4**). In addition, we observed suggestive clustering of mutations at the *TP53* transcription start site, the *TP53* polyadenylation signal, and at splice sites in the first non-coding exon of *TP53* (**Fig. 3f**), as well as hotspots in non-canonical but previously reported 5′ UTR sites in *TERT*, and a striking cluster of mutations upstream of *AJUBA* overlapping a CTCF binding site. These analyses also revealed a general inflation of mutations in the core promoters of many genes in the panel, suggestive of a higher background mutation rate in promoters rather than selection (**Fig. 3g**), consistent with previous reports ^41^. Despite our panel not being designed to search for non-coding cis-regulatory driver mutations, these examples show the potential of deep clonal scanning to exhaustively discover coding and non-coding driver mutations.

Variants of uncertain significance (VUS) are germline or somatic variants identified by genetic testing whose clinical relevance is unknown. Evidence of selection in cancer is starting to be used for the classification of germline and somatic variants in some genes ^42^ but is limited by the sparsity of cancer genomic datasets. To investigate whether selection in normal tissues could contribute to these efforts, we compared the distribution of site-dN/dS ratios for sites annotated in ClinVar as pathogenic, benign or of uncertain significance. Nearly all known pathogenic sites in *TP53*, *NOTCH1* and *PPM1D* have high site-dN/dS ratios, and nearly all known benign sites have low site-dN/dS ratios (**Fig. 3h)**. Looking at sites reaching or approaching significance (q-value<0.20), we find many VUS (and zero benign variants) with comparable evidence of selection to known pathogenic variants (e.g. 86 in *TP53*, 35 in *NOTCH1*, 5 in *PPM1D*) (**Extended Data Table 4**). Although deeper sequencing will be required to achieve true saturation (**Supplementary Note 4**), these results suggest that deep single-molecule sequencing of polyclonal tissues has the potential to provide in vivo saturation mutagenesis information for genes under clonal selection.

### Mutational epidemiology, mutagens and selectogens

The discovery of many clones carrying cancer-driver mutations in normal tissues has caused some confusion about their role in carcinogenesis. However, these clones are entirely compatible with a multistage model of carcinogenesis, and were in fact anticipated by some classical mathematical models (see **Supplementary Note 5** for an extended description). In the 1950s, Nordling, Armitage and Doll proposed that the rapid increase in cancer incidence with age could be explained by a model where cells acquire mutations linearly with age and 6-7 driver events are required for transformation ^43,44^. Lesser-known models with clonal expansions were proposed soon after and showed that the size and type of clonal expansion had large effects on cancer incidence. The current paradigm of carcinogenesis is that cancers emerge by somatic evolution. Both mutation and selection (clonal expansion) increase the likelihood of a cell acquiring the complement of driver changes needed for transformation. Carcinogens may thus act by inducing mutations (mutagens) or by altering selection and favouring clonal expansions (promoters ^45,46^ or selectogens ^47^) (**Supplementary Note 5**). By studying the variation in mutation and selection across 1,042 individuals, we can begin to quantify these processes.

To investigate the mode of clonal growth in oral epithelium, we first studied how the frequency of driver mutations increases with age in our cohort. Interestingly, this showed that the estimated fraction of cells carrying driver mutations increases roughly linearly with age, through the accumulation of large numbers of small clones, with the VAF of the largest clone per individual growing slowly or plateauing with age (**Fig. 4a**). Since new driver mutations occur continuously, this observation is inconsistent with models of continued clonal growth, including exponential growth, quadratic growth (expected if clones grow only at their edges) ^48^, and models predicting an acceleration of selection during ageing ^49^. Instead, the pattern seems more consistent with a model where clone sizes are constrained (by cell-intrinsic or cell-extrinsic mechanisms) (see **Supplementary Notes 5,6, Extended Data Fig. 7**). This contrasts with the pattern observed in blood, where both the driver density summed across clones and the size of the largest clone increase almost exponentially with age, consistent with previous clonal haematopoiesis studies ^50,51^. While detailed modelling of clonal dynamics is beyond the scope of this study, we note that the slower-than-expected increase in driver density with age and the small size of epithelial clones should be major barriers to carcinogenesis (**Supplementary Note 5**).

**Figure 4.**
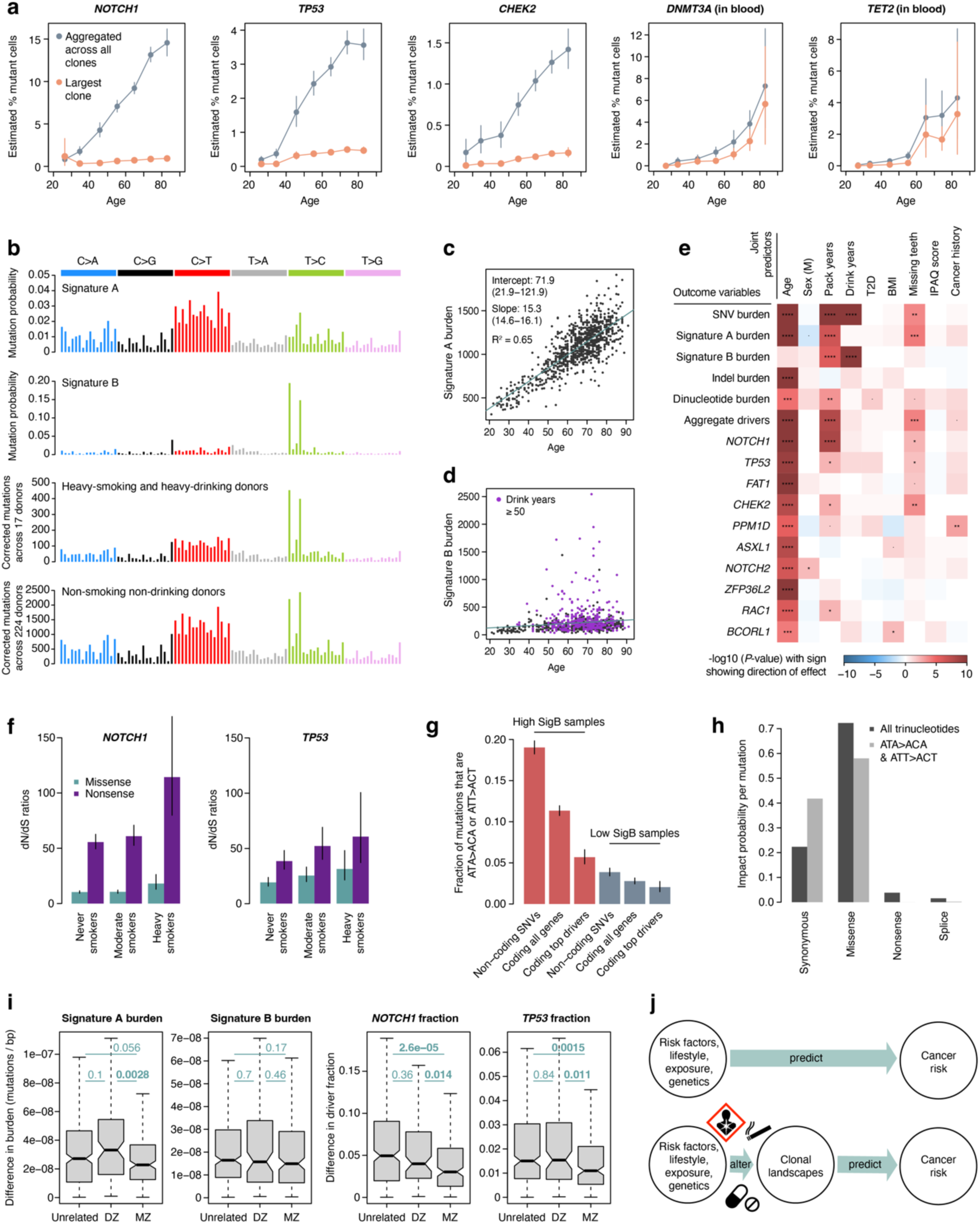
Mutational epidemiology in oral epithelium. **a**, Estimated percentage of cells carrying a mutation in each of five selected genes, as a function of donor age. Mutant cell percentages are shown for the largest clone (orange) and for all mutant clones (grey). Values for *NOTCH1*, *TP53* and *CHEK2* were calculated from oral epithelium samples, whereas values for *DNMT3A* and *TET2* were calculated from blood samples. **b**, Trinucleotide mutational spectra for (top to bottom) inferred Signatures A and B, and mutations in oral epithelium from heavy-smoking, heavy-drinking donors (*n*=17) and oral epithelium from non-smoking, non-drinking donors (*n*=224). Mutational spectra are corrected by the ratio of genomic to observed trinucleotide frequencies. **c, d**, Linear regressions of genome-wide Signature A and Signature B burdens in oral epithelium against donor age. For Signature A, the intercept and slope (point estimates and 95% CIs) and coefficient of determination (*R*^2^) of the fitted model are indicated. For Signature B, donors with a record of ≥50 drink-years are highlighted. **e**, Heatmap of associations between different measures of mutation burden, signature burden or driver density (*y*-axis) and relevant donor metadata (*x*-axis), inferred using linear mixed-effects regression models. The *P*-value of each association is indicated by both colour shading (red and blue for positive and negative associations, respectively) and asterisk labels (****: *q* < 10^−4^; ***: *q* < 10^−3^; **: *q* < 0.01; *: *q* < 0.05; •: *P* < 0.05). **f**, dN/dS ratios for missense and nonsense mutations in *NOTCH1* and *TP53*, with donors grouped by smoking status. Error bars denote 95% CIs. **g**, Fraction of ATA>ACA or ATT>ACT SNVs in samples presenting high (top 5%, red) or low (bottom 5%, blue) exposure to Signature B (SigB). Fractions are presented for non-coding SNVs, coding SNVs in all genes, and coding SNVs in top driver genes (defined as genes with >1,000 driver mutations across mutational classes with >80% estimated driver fraction). Error bars denote 95% CIs. **h**, Distribution of predicted mutation consequences for substitutions in any trinucleotide context (dark grey) and for ATA>ACA or ATT>ACT substitutions only. **i**, Distributions of pairwise differences in generalised linear model residuals for Signature A and B mutation burdens and *NOTCH1* and *TP53* driver fractions, between pairs of unrelated (age-matched) individuals, pairs of dizygotic (DZ) twins, and pairs of monozygotic (MZ) twins. The *P*-values for significantly different pairs of distributions (two-sided Mann–Whitney–Wilcoxon tests) are highlighted in bold. **j**, Non-mechanistic (top) and mechanistic (bottom) risk models connecting predictor variables to cancer risk. Mechanistic risk models can offer insight into the impact of risk factors on mutational or clonal landscapes and may be used to predict cancer risk.

Targeted NanoSeq also provides information on mutation rates and signatures across individuals. Performing signature decomposition on all 1,042 donors, we found two signatures responsible for most mutations in our data (**Fig. 4b**). Signature A (SigA) resembles a combination of COSMIC SBS5 and SBS1 (94% and 6% respectively, cosine similarity 0.90, **Methods**). SBS5 is a ubiquitous clock-like signature observed across tissues, believed to result from the occasional misrepair of the continuous DNA damage suffered by all cells ^11,12,52,53^, whereas SBS1 results from the deamination of 5-methylcytosine. SigA is largely responsible for the life-long accumulation of mutations in oral epithelium, with a slope of ∼15.3 mutations per cell per year (**Fig. 4c**, R^2^=0.65, *P*<2.2×10^−16^). Signature B (SigB) resembles COSMIC SBS16 (cosine similarity 0.975), a common signature in head and neck epithelia and liver, associated with alcohol consumption and aldehyde metabolism ^4,54,55^ (**Extended Data Fig. 8**). SigB showed extreme variation across donors, contributing low numbers to most individuals but >1,000 mutations per cell in some heavy drinkers (**Fig. 4d**, **Supplementary Note 3**).

Oral cancer risk factors may be expected to increase mutation rates or induce clonal expansions. To test for mutagenic and selectogenic effects while accounting for confounders and twin relationships, we used multivariate mixed-effect regressions using major risk factors and other relevant metadata as covariates, and different measures of mutation rate, signatures, or driver density as outcome variables (**Fig. 4e**, **Supplementary Note 7, Extended Data Fig. 9**). As expected, SNVs, indels and dinucleotide variants, as well as SigA (but not SigB), and the density of all major drivers, increased strongly with age. Interestingly, sex was not significantly associated with differences in mutation rates, signatures or driver densities when correcting for confounders. Tobacco smoking is a major oral cancer risk factor, and we found pack-years to be strongly associated with total SNVs, SigA and SigB, dinucleotide substitutions (but not indels), overall driver density across genes, *NOTCH1* driver density, and nominally significantly associated with three other drivers with lower statistical power. Alcohol consumption is another major oral cancer risk factor, and we found estimated drink-years to be strongly associated with SNV and SigB burden, but not SigA burden, consistent with the known aetiology of SigB/SBS16 (**Methods**). Poor oral health is also an oral cancer risk factor ^56^, and we found that the number of missing teeth correlated with SigA burden and overall driver density. We were unable to study the effect of oral human papillomavirus (HPV) infection in our dataset, which is an increasingly important risk factor for oral cancer particularly in younger individuals ^57^, as HPV infection history was unavailable and sequencing-based detection of HPV in our data yielded limited information (**Methods**), possibly due to the predominance of older donors in our cohort.

The strong association of alcohol consumption with SigB is believed to result from DNA damage by alcohol-derived aldehydes ^58^. However, the mechanistic basis for the association of smoking with SigB is less clear. Some analyses in our dataset suggest that smoking may increase SigB by exacerbating the mutagenic effects of alcohol consumption, consistent with epidemiological studies ^59^ (**Supplementary Note 7**). However, we cannot rule out the possibility that this association is partially caused by inaccurate self-reporting of alcohol consumption. While our models also suggest that smoking and poor oral health are significantly associated with an increase in SigA/SBS5, the mechanistic bases for these associations remain unclear. Notably, these regressions suggest that one additional year of life causes as many mutations in the oral epithelium as ∼2.8 pack-years or ∼19.1 drink-years (see **Supplementary Note 7** for caveats and interpretation).

Additional regression models can help disentangle the mutagenic or selectogenic mode of action of some risk factors. If a carcinogen acts solely as a mutagen without altering selection, driver density should increase proportionally to the increase in mutation burden, at least under some assumptions. Pure promoters or selectogens may be expected to alter clonal selection without changes in mutation rates, and dual carcinogens may be expected to alter both (see **Supplementary Note 7**). To test for selectogenic effects, we used regressions with driver density as the outcome variable correcting for the increase in mutation burden as a covariate. Using several different models suggested that most of the association between driver densities and oral cancer risk factors (pack-years, drink-years, and oral health), is explained by mutagenesis rather than selectogenesis (**Supplementary Note 7**). Putative selectogenic associations included an increase in *NOTCH1* clones with smoking (**Fig. 4f**), and *CHEK2* with poor oral health (**Extended Data Fig. 9**), as well as trends for other genes. Since these associations are only correlative and may be influenced by confounders, additional studies will be needed to confirm and extend these observations. However, these analyses show the potential of mutational epidemiology studies to illuminate the mode of action of major cancer risk factors.

Intriguingly, alcohol consumption seemed to induce fewer driver mutations than expected from the increase in mutation rates. Analysis of the distribution of SigB mutations revealed that this is due to a low driver-generation potential of SigB/SBS16, as ATA>ACA or ATT>ACT SigB mutations are heavily biased towards intronic sequences (**Fig. 4g,h**).

Finally, this dataset offers an opportunity to start investigating germline influences on somatic mutation rates. First, we leveraged our twin design to test for evidence of heritability. We compared the difference in mutation rates and driver frequencies between identical (monozygotic [MZ]), non-identical (dizygotic [DZ]), and unrelated same-age pairs of donors, while accounting for confounders (**Supplementary Note 8**). This revealed modest but suggestive evidence of heritability for SigA/SBS5 (MZ vs DZ *P*=0.004), *NOTCH1* (*P*=0.023) and *TP53* (*P*=0.018) (**Fig. 4i**). Similar signals were found with more formal ACE and genomic-relatedness tests (**Supplementary Note 8**). Second, although our statistical power is low, we performed genome-wide association studies (GWAS) of mutation rates and driver densities for completeness, finding no genome-wide significant associations (**Supplementary Note 8**). Overall, heritability analyses suggest that germline factors likely influence mutation rates, signatures and driver landscapes, but larger studies are needed to identify these associations. We envision that larger single-molecule sequencing studies of non-invasive samples should enable the systematic identification of germline mutations influencing somatic mutation rates, which might help illuminate the mechanistic bases of SBS5, and perform causal inference of the role of somatic mutations across common diseases ^60^.

## Discussion

Building on duplex sequencing, we have developed a new version of NanoSeq that achieves accurate somatic mutation detection on single DNA molecules (<5 errors/gigabase) while being compatible with whole-genome, whole-exome and deep targeted sequencing. This method greatly simplifies the study of somatic mutation rates, signatures and driver landscapes in any tissue, regardless of clonality.

Applying targeted NanoSeq to oral epithelium, we have unveiled an unprecedentedly rich landscape of selection in a normal solid tissue, with 49 genes under positive selection, over 62,000 driver mutations and several genes under negative selection. These data also exemplify how deep single-molecule sequencing of highly polyclonal tissues can be used to build high-resolution maps of selection across sites in genes under strong selection. This method has the potential to complement in vitro saturation mutagenesis efforts to help variant annotation for genetic diagnosis. Whereas this approach is limited to genes under selection in a tissue, a wider range of disease-relevant genes can be assayed across tissues (Neville *et al*, companion paper).

The ability to measure somatic mutation rates, signatures and driver landscapes in large sample cohorts offers several opportunities to augment traditional cancer epidemiology. First, systematic studies of the mutation landscape across individuals with different risk factors, as well as case-control studies and intervention studies, could help build mechanistic risk models connecting risk factors to mutation and clonal landscapes, and these landscapes to cancer risk (**Fig. 4j**). Such studies could provide insights into the mode of action of poorly understood exposures or risk factors (e.g. obesity), as well as enable risk prediction or stratification. Second, studies of the mutation landscapes of normal tissues in countries or populations with unusually high rates of certain cancers could shed light on unknown exposures, potentially helping develop prevention strategies. Third, mutation and clonal landscapes may be informative as surrogate risk markers in cancer prevention and molecular prevention trials. Although molecular prevention of cancer in the general population is rarely discussed, the discovery of simple markers of cardiovascular disease risk, such as low-density lipoprotein cholesterol and hypertension, enabled the development of statins and antihypertensive medications, which have transformed the management of cardiovascular disease ^61^.

Beyond cancer, somatic mutations have long been speculated to contribute to ageing and other diseases. Suggestive associations have now been found between somatic mutations in certain genes and multiple diseases^62–64^. However, systematic studies in polyclonal conditions have not been possible with available technologies. Accurate whole-exome single-molecule sequencing has the potential to enable sensitive and unbiased discovery of somatic driver mutations in any tissue, in health and disease.

## Data Availability

All data produced in the present study will be available through managed access via the TwinsUK Resource Executive Committee (TREC).

## Acknowledgements

We are grateful to members of the TwinsUK cohort for volunteering samples and metadata for this study. We thank G. Davey Smith and P. M. Visscher for advice on epidemiology and heritability analyses; A. Teschendorff for advice on methylation; A. P. Butler and V. Offord for assistance with establishing the bioinformatic pipeline; Y. Hooks for sample processing; K. Roberts, T. Baxter, K. Smith, N. Yilmaz, V. Uksaite, E. Ferla, H. Savin, L. Allen and C. Latimer for helping with sample shipments; and the CASM support and DNA pipeline teams at the Wellcome Sanger Institute for their essential role in data generation. **Funding:** I.M. is funded by Cancer Research UK (C57387/A21777), the Dr Josef Steiner Cancer Research Foundation and the Wellcome Trust. TwinsUK is funded by the Wellcome Trust, Medical Research Council, Versus Arthritis, European Union Horizon 2020, Chronic Disease Research Foundation (CDRF), Zoe Ltd, the National Institute for Health and Care Research (NIHR) Clinical Research Network (CRN) and Biomedical Research Centre based at Guy’s and St Thomas’ NHS Foundation Trust in partnership with King’s College London.

## Author contributions

A.R.J.L., F.A., P.A.N. and I.M. conceptualised the project with support from M.R.S., P.J.C., R.R., and K.S.S. A.R.J.L., F.A., G.K., A.B.-O., and I.M. led the data analysis with support from P.A.N., A.L.R., M.D.C., M.J.P., N.W., D.A., L.M.R.H., J.R.B. and M.D.Y. A.R.J.L., P.A.N. and S.V.L. led the experimental work with support from N.B., S.W., W.C., M.Mo., L.S., M.Ma. and N.M.-S. F.A., R.E.A., T.C., A.B. and I.M. developed algorithms and software. A.R.J.L., F.A., S.V.L., S.W. and I.M. contributed to method development. A.L.R., J.S.E.-S.M., D.V., B.J., A.N., S.W., K.T.A.M. and K.S.-P. collected samples. L.O. and S.A.-G. helped with sample and project administration. A.L.P. and D.M.R. provided histology support. K.S.-P., H.C.M., M.R.S., P.J.C., R.R., K.S.S., and I.M. provided supervision. A.R.J.L., F.A., P.A.N., A.B.-O., and I.M. wrote the manuscript, and all authors contributed to reviewing and editing it.

## Extended Data Figures

**Extended Data Figure 1.**
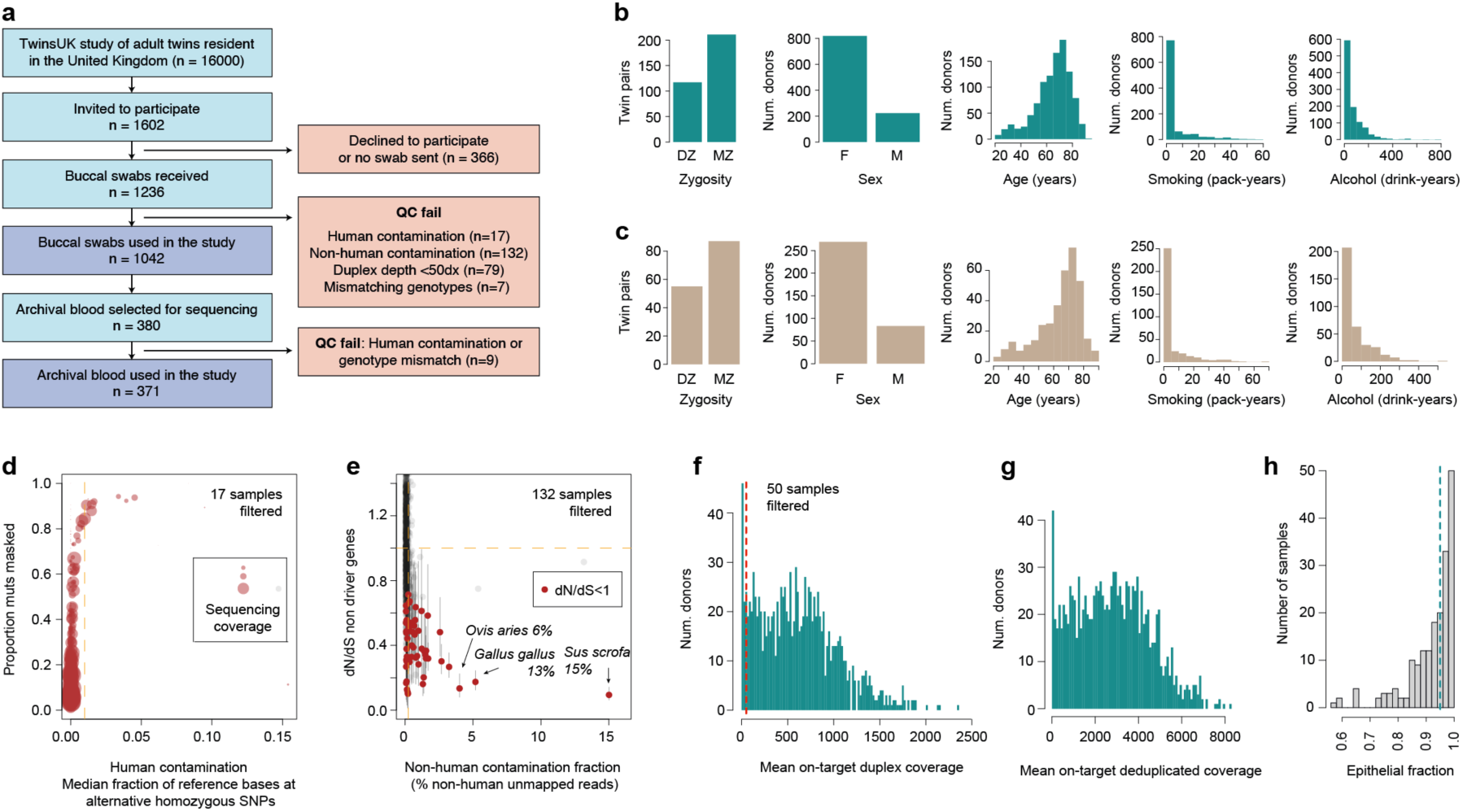
Description of the cohort. **a**, Flow diagram describing the selection of the donor cohort used in this study. **b**, **c**, Distribution of zygosity, sex, age, smoking (pack-years) and drinking (drink-years) values for (**b**) buccal swab and (**c**) blood sample donors. **d**, Identification of samples contaminated with human DNA from another individual, comparing the proportion of mutation calls falling in the ‘SNP+noise’ mask *versus* the median fraction of reference bases at alternative homozygous SNPs; point size is proportional to the duplex coverage. The vertical dashed line indicates our exclusion criterion for human contaminated samples (>0.01). **e**, Identification of samples contaminated with non-human DNA, comparing the dN/dS values in passenger genes *versus* the percentage of unmapped reads mapping to a set of potential contaminant species. Horizontal dashed line indicates neutral dN/dS=1. Red points indicate samples with upper bound 95% CI dN/dS ratio < 1. Vertical dashed line shows our exclusion criterion for non-human contaminated samples (>0.25). **f**, Histogram of duplex coverage (dx) in the buccal swab cohort, at on-target and near-target regions. Vertical dashed line shows our exclusion criterion for low coverage samples (<50dx). **g**, Distribution of the mean deduplicated coverage (×) in the buccal swab cohort, at on-target and near-target regions. Raw sequencing coverage is ∼6.6 times higher due to the average 85% duplicate rate required for duplex consensus calling. **h**, Estimation of epithelial fraction in buccal swab samples by targeted enzymatic methylation sequencing. Vertical dashed line shows the median epithelial fraction of 0.95.

**Extended Data Figure 2.**
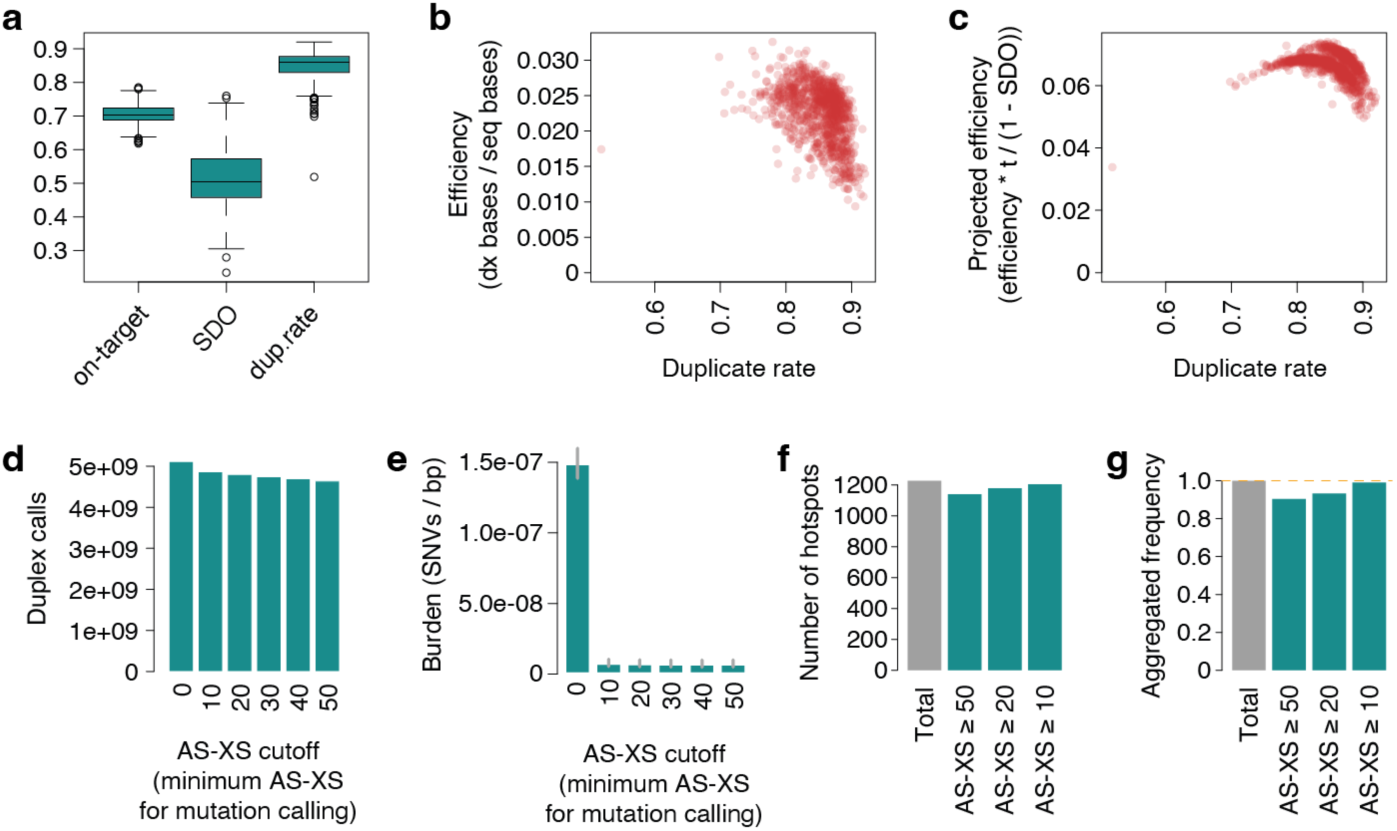
Targeted NanoSeq quality metrics. **a**, Sequencing quality metrics including the on-target capture fractions, estimated excess in strand drop-out (SDO), and the achieved duplicate rates for the buccal swab cohort. By random sampling it is expected to miss one of the original DNA strands in a proportion of cases, following a binomial distribution. We estimated the excess in SDO by calculating and subtracting the observed and expected SDOs. **b**, Relationship between duplicate rates and sequencing efficiency, measured as the number of bases with duplex support divided by the total number of bases sequenced. **c**, Relationship between duplicate rates and sequencing efficiency after factoring in the on-target fraction (t) and the excess in strand drop-out (SDO). **d**, Number of duplex calls as a function of the AS-XS (primary alignment score minus secondary alignment score) threshold **e,** Substitution burdens calculated within each AS-XS threshold corrected for trinucleotide context. Error bars for substitution burdens indicate Poisson 95% CIs. **f**, **g**, Hotspots covered by different AS-XS thresholds, shown as (**f**) total number of hotspots and (**g**) their aggregated frequency in TCGA. Horizontal dashed line indicates the detection of all studied hotspots.

**Extended Data Figure 3.**
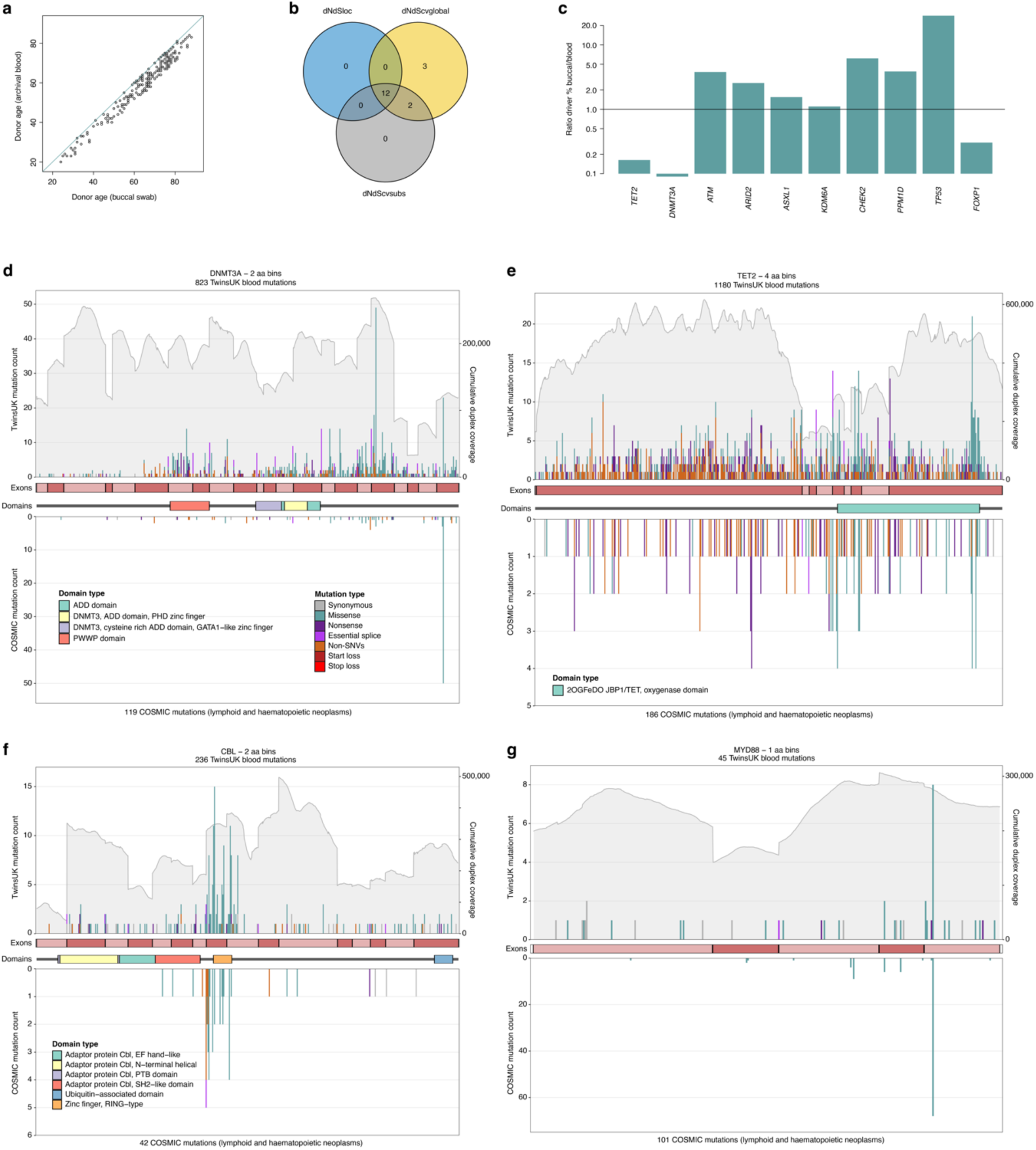
Further description of the blood driver landscape. **a**, Relationship between donor ages (years) for matched buccal swab samples and archival blood samples. The diagonal line represents the identity function, *y*=*x*. **b**, Venn diagram summarising the overlaps between three approaches for identifying genes under significant positive selection by *dNdScv*. **c**, Ratio of estimated driver densities between buccal swab samples and archival blood samples, for 10 genes identified as being under gene-level significant positive selection in both blood and buccal swab samples. **d-g**, Mutation histograms for *DNMT3A*, *TET2*, *CBL* and *MYD88*. The *x*-axis represents coordinates along the coding sequence. Exons and protein domains are indicated along the *x*-axis. The *y*-axis represents number of mutations, either in the 371 TwinsUK blood samples used in this study (top) or in lymphoid and haematopoietic neoplasm samples (whole-exome and whole-genome) from the COSMIC database (bottom). Mutations are coloured according to mutation consequence category. Grey shading indicates cumulative duplex coverage across TwinsUK blood samples.

**Extended Data Figure 4.**
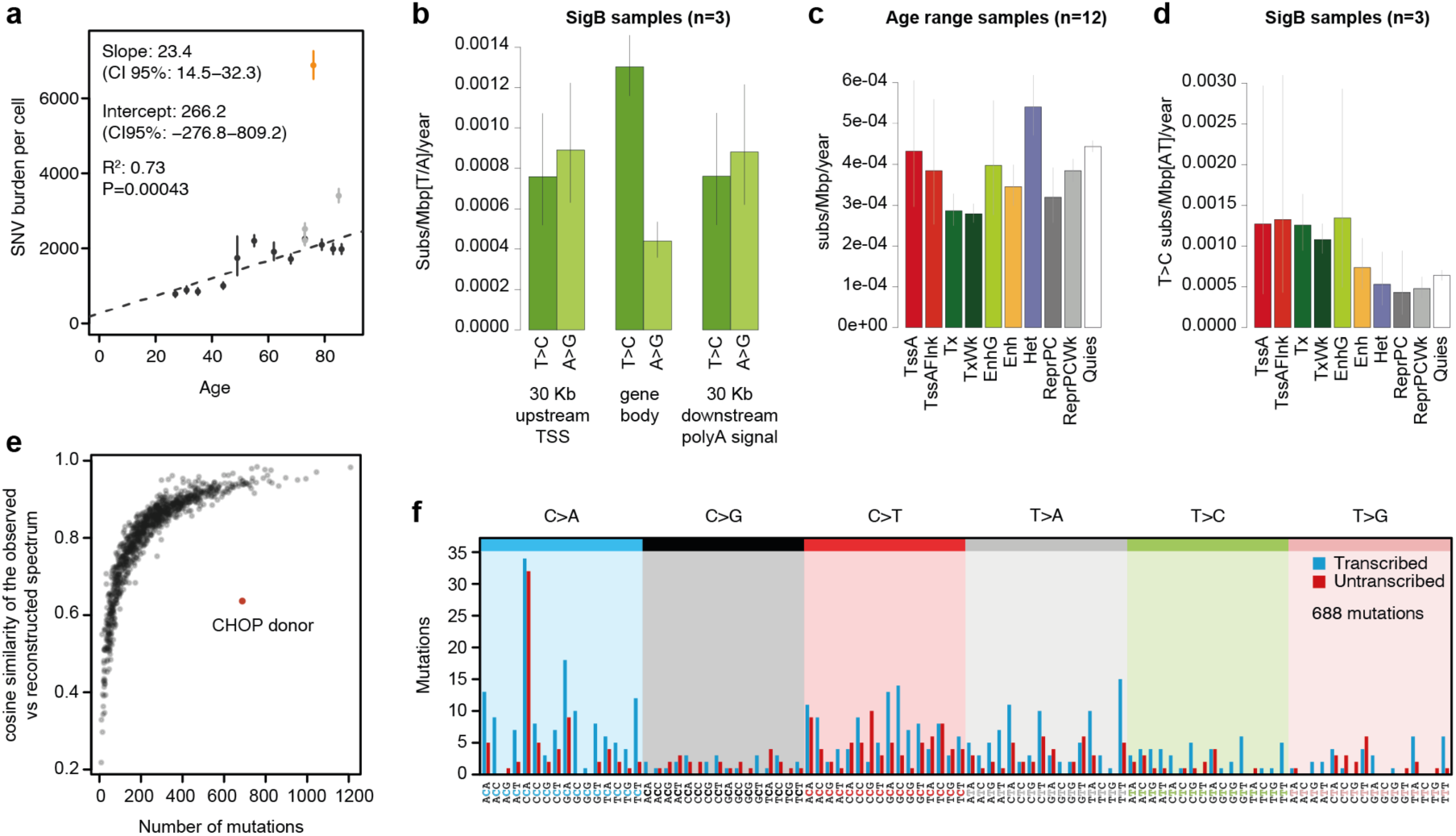
Whole-genome NanoSeq on buccal swabs. **a**, Regression of SNV mutation burden with age for 12 samples selected from across the age range (black dots), three samples with high SigB signature contribution (grey), and one sample with a high mutation burden from a donor with a history of CHOP chemotherapy treatment (orange). The regression results listed within the plot were generated using only the 12 samples randomly selected for their age range. Error bars show Poisson 95% CIs for the estimated burdens. **b**, Transcription-coupled repair and damage in three donors with high contribution of SigB. Estimated substitution burdens plus their associated 95% CIs (error bars) across upstream, transcribed and downstream regions, showing T>C and A>G in the coding strand separately. **c**, Number of substitutions per Mbp per year in 12 age range donors for each of 10 major ENCODE chromatin states. Reference chromatin states were obtained from ENCODE E057 foreskin keratinocytes. Chromatin states BivFlnk, EnhBiv, TssBiv, TxFlnk, and ZNF/Rpts were removed given their smaller footprint and too large confidence intervals. Burdens were normalised to whole genome trinucleotide frequencies. **d**, Number of substitutions per Mbp per year in 3 donors with strong SigB exposure for each of 10 major ENCODE chromatin states. Only T>C rates are shown, calculated as the number of T>C substitutions observed and divided by the number of [TA] bps. **e**, Cosine similarities between the observed and reconstructed substitution profiles as a function of the number of mutations in each sample, highlighting the outlier donor who underwent CHOP chemotherapy (brown). **f**, 192 substitution profile for the outlier CHOP donor.

**Extended Data Figure 5.**
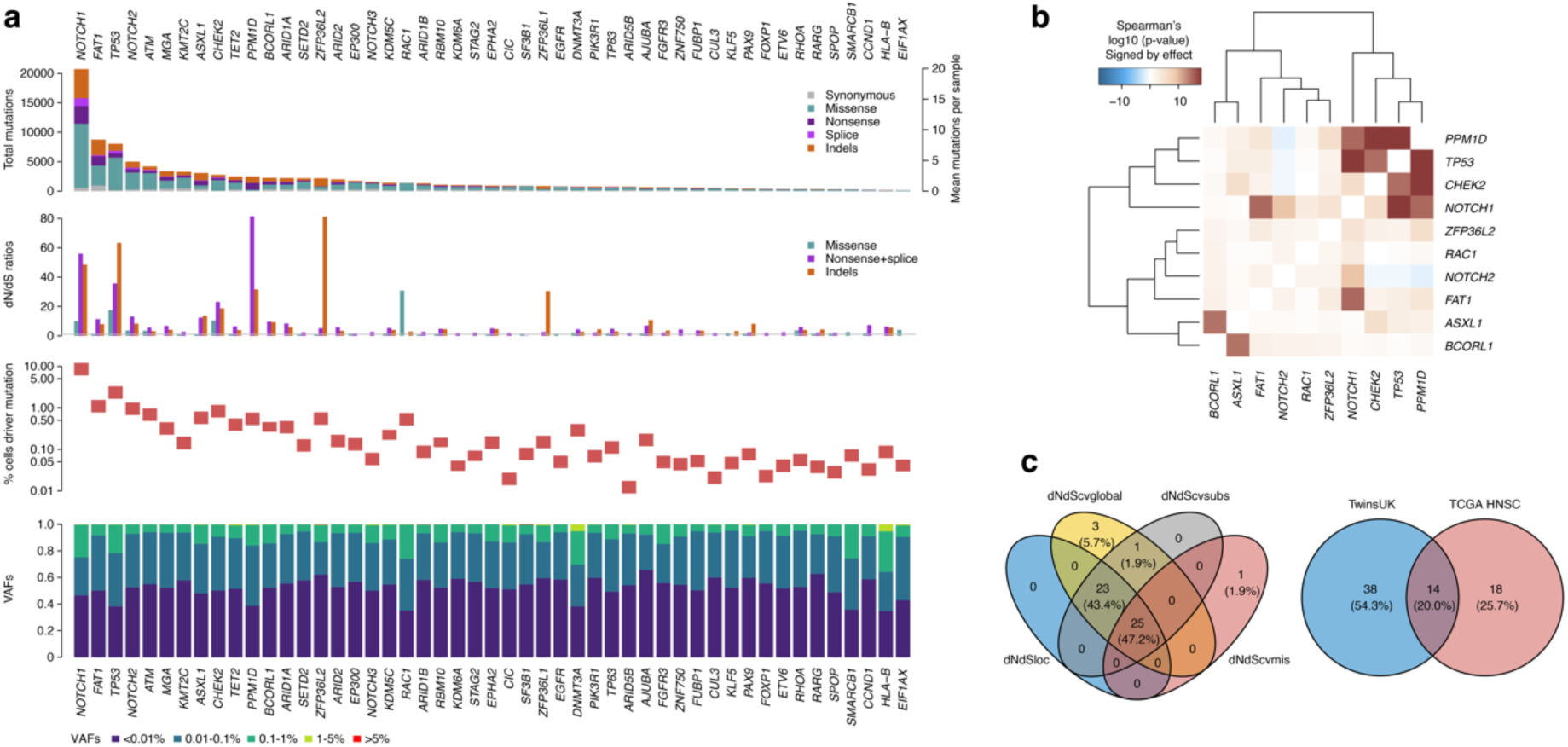
Full buccal driver landscape. **a**, For the 49 significant driver genes in oral epithelium, panels show (top to bottom) mutation counts per mutation consequence category, dN/dS ratios per mutation consequence category (horizontal line indicates neutral dN/dS=1), estimated mutant cell percentages in donors aged 65-85, and the distribution of unbiased VAFs. **b**, Correlation between the generalised linear model residuals for driver burden across top driver genes (defined as genes with >1,000 driver mutations across mutational classes with >80% estimated driver fraction. **c**, Venn diagrams summarising (left) the overlaps between four approaches for identifying genes under significant positive selection genes in oral epithelium by dNdScv, and (right) driver genes in TwinsUK oral epithelium samples and head and neck squamous cell carcinomas (HNSC) in The Cancer Genome Atlas.

**Extended Data Figure 6.**
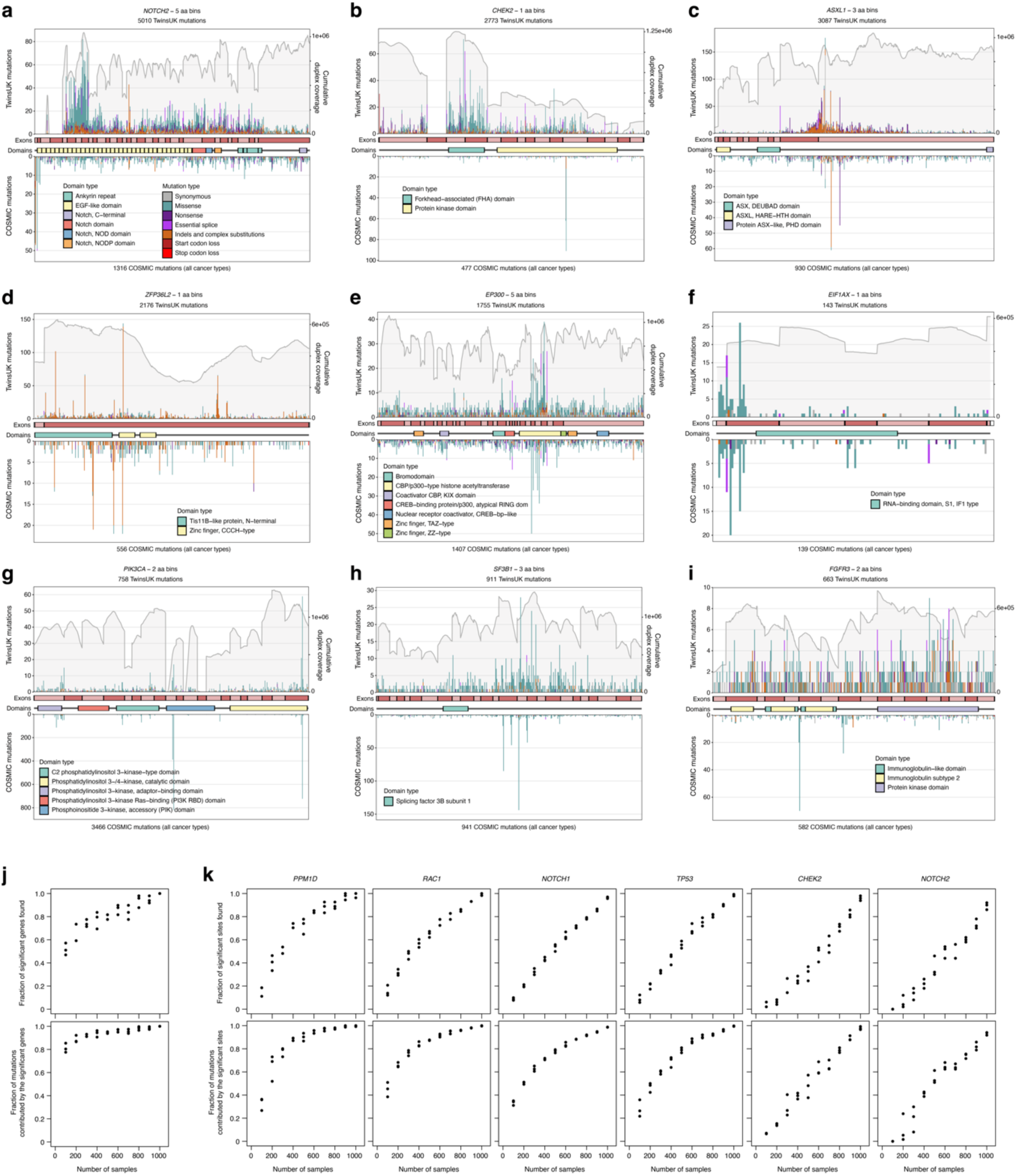
Distribution of mutations within selected buccal driver genes. **a-i**, Mutation histograms for nine selected genes. The *x*-axis represents coordinates along the coding sequence. Exons and protein domains are indicated along the axis. The *y*-axis represents number of mutations, either in the 1,042 TwinsUK oral epithelium samples used in this study (top) or across whole-exome and whole-genome samples of any cancer type in the COSMIC database. Mutations are coloured according to mutation consequence category. Grey shading indicates cumulative duplex coverage across TwinsUK samples. **j**, Fraction of significant genes (top) and fraction of mutations contributed by genes identified as significant (bottom) identified by gene-level dN/dS across subsamples of the buccal swab cohort. **k**, Fraction of significant sites (top) and fraction of mutations contributed by sites identified as significant (bottom) identified by site-level dN/dS for six genes across subsamples of the buccal swab cohort.

**Extended Data Figure 7.**
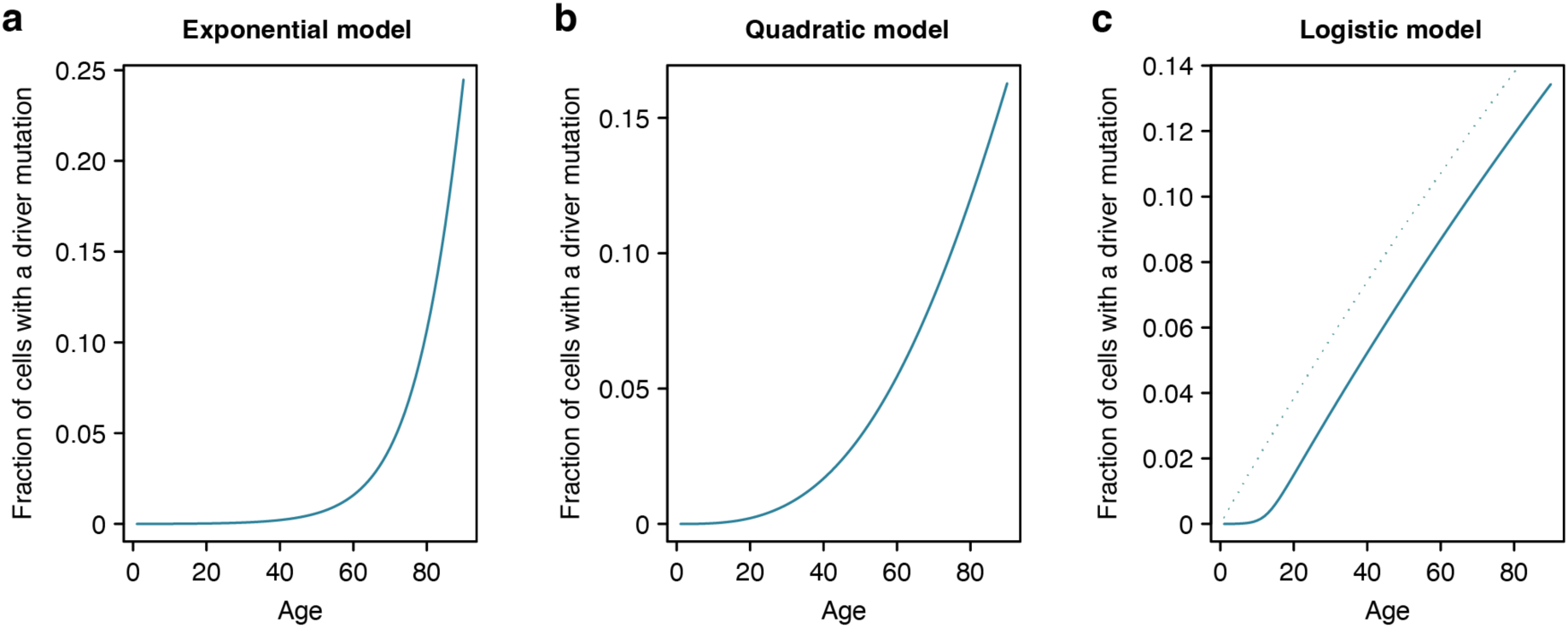
Models of clonal growth. **a-c,** Graphs illustrating the increase in the fraction of cells carrying a driver mutation as a function of age, as predicted by (**a**) exponential, (**b**) quadratic and (**c**) logistic models of clonal growth. The dotted line in (**c**) represents the prediction from a linear growth model. In all three cases, we used a driver mutation rate per cell per year of *μ*=4×10^−6^ (1000 driver sites per genome, 4×10^−9^ mutations per cell per year). Other parameters for the three growth models were chosen to obtain a fraction of cells with a driver mutation around 10-15% by age 80, to facilitate comparison to the buccal swab data in Fig. 4a. The parameters used are the following: (**a**) exponential model using *r*=0.1, (**b**) quadratic model using *r*=0.2, (**c**) logistic model using *L*=500 and *r*=0.5.

**Extended Data Figure 8.**
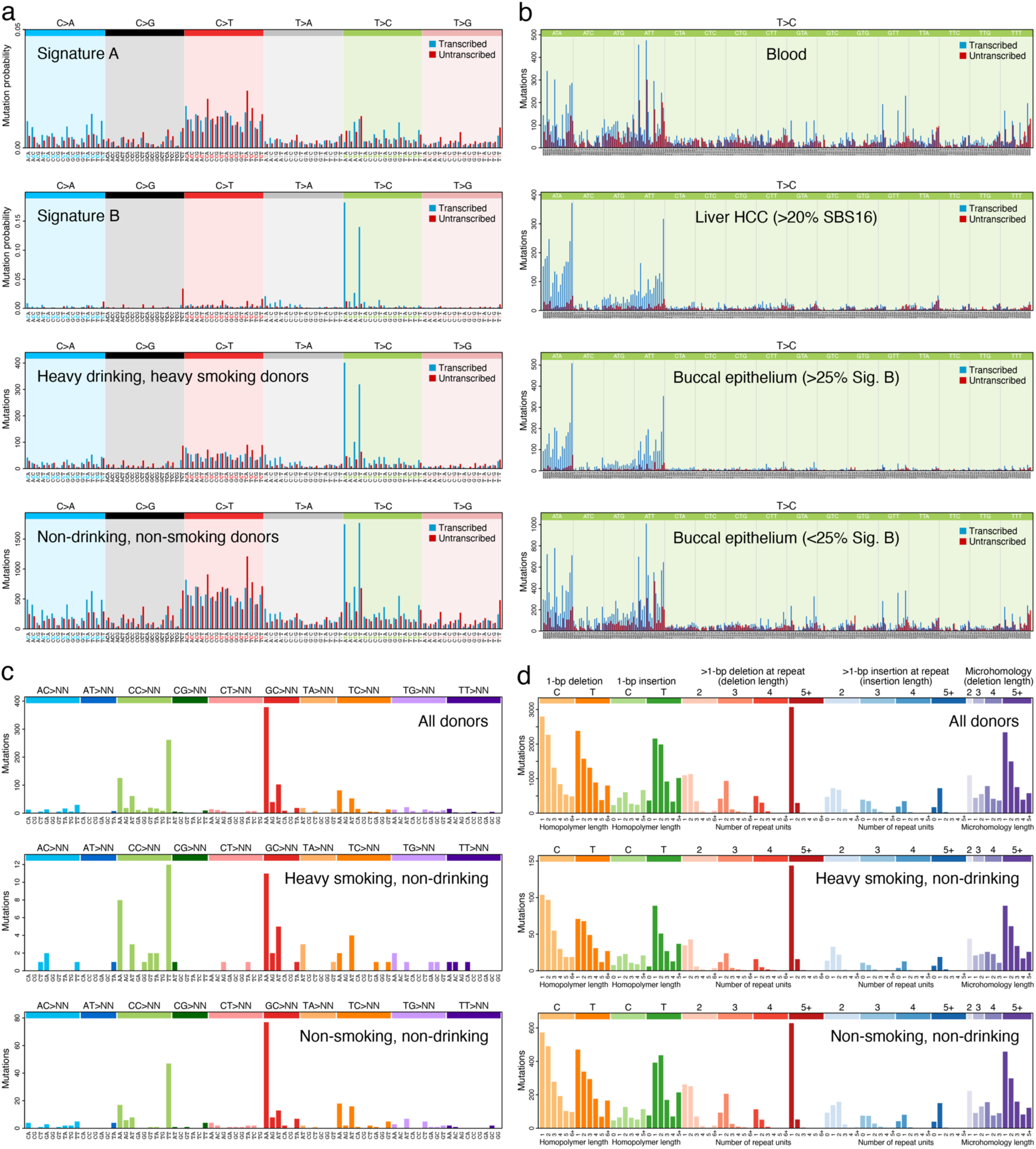
Mutational spectra of somatic single-base substitutions (SBSs), double-base substitutions (DBSs) and indels. **a**, Transcriptional strand-wise versions of the trinucleotide SBS spectra shown in Fig. 4b. **b**, Transcriptional strand-wise pentanucleotide spectra of T>C SBSs in (top to bottom): blood samples (n=371), hepatocellular carcinoma samples (Liver HCC; **Supplementary Note 3**) with >20% SBS16 exposure (n=4), oral epithelium samples with >25% Signature B exposure (n=121), and oral epithelium samples with <25% Signature B exposure (n=921). **c**, DBS spectra in (top to bottom): all oral epithelium samples (n=1042), oral epithelium from heavy smoking, non-drinking donors (n=27), and oral epithelium from non-smoking, non-drinking donors (n=224). **d**, Spectra of indels in the same sample sets shown in **c**.

**Extended Data Figure 9.**
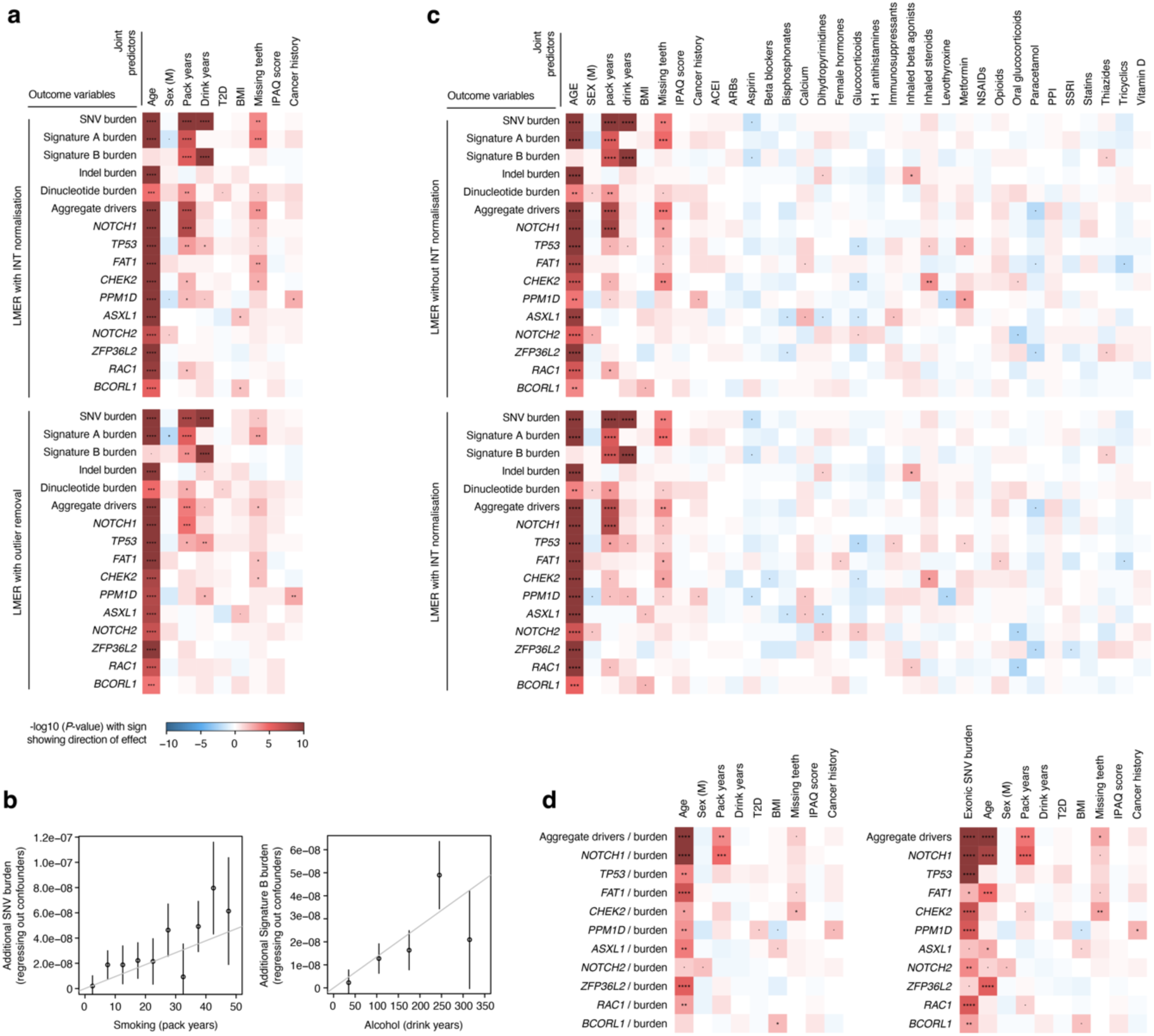
Additional regression models. **a**, Heatmap of associations between different measures of mutation burden, signature burden or driver density (*y*-axis) and relevant donor metadata (*x*-axis), inferred using linear mixed-effects regression (LMER) models with (top) inverse-normal transformation (INT) or (bottom) outlier removal, where outliers are defined as those values larger than 3 × IQR + Q_3_ (where IQR is the interquartile range and Q_3_ is the third quartile for each outcome variable). The *P*-value of each association is indicated by both colour shading (red and blue for positive and negative associations, respectively) and asterisk labels (****: *q* < 10^−4^; ***: *q* < 10^−3^; **: *q* < 0.01; *: *q* < 0.05; •: *P* < 0.05). **b**, Additional SNV burden obtained after regressing out confounders using an LMER model for (left) smoking history across pack-year bins and (right) alcohol consumption history across drink-year bins. **c**, Heatmap (as described in **a**) with medication history included as predictors. **d**, Heatmap (as described in **a**) using a multivariate LMER model with the driver density per gene per donor normalised by the exonic mutation burden in passenger genes per donor. Normalisation was achieved by either (left) using the ratio of driver density and mutation burden as a new outcome variable for each driver gene or (right) including the mutation burden as a covariate.

Supplementary Information is available for this paper.

Correspondence and requests for materials should be addressed to I.M. Reprints and permissions information is available at www.nature.com/reprints.

## Methods

### Cohort selection

The TwinsUK study contains around 16,000 participants. From a preselection of 4,800 donors, we invited 1,796 to participate based on several criteria, receiving buccal swabs from 1,236 (**Extended Data Fig. 2a**). To increase our statistical power to study associations with exposures, risk factors and germline factors, we included all available donors of age 80 or higher (*n*=230), as many complete twin pairs as possible, smokers, individuals with obesity (BMI > 30), and individuals with available genome-wide genotyping information. We also favoured the selection of males and individuals of non-white ethnicity to reduce some of the demographic biases in the TwinsUK registry compared to the general population. To test for associations between the mutational landscape and medications or clinical histories, we favoured the inclusion of individuals with a history of cancer (including all donors with a history of oral cancer, *n*=12) or a self-reported treatment history including tamoxifen, immunosuppressants, metformin, aspirin or ibuprofen. 194 samples were excluded from analysis based on several sequencing quality metrics, leaving a total of 1,042 samples in the study. Exclusion criteria included: removal of contaminated samples with either human (*n*=17) or non-human (*n*=132) DNA, exclusion of samples with mean duplex coverage lower than 50 (*n*=79), and exclusion of swabs with genotyping information not matching the pre-existing genotyping information from TwinsUK (*n*=7) (**Extended Data Fig. 2a**).

From the final 1,042 donors in the study, we also selected 380 individuals with available material the the TwinsUK BioBank for sequencing of archival whole-blood DNA. 371 samples passed quality controls for study inclusion. The selection of blood donors was based on several criteria: 12 donors (and their twins) treated with metformin, 30 donors (and their twins) with the highest mutation burden per year in the buccal swabs, 25 donors (and their twins) with the highest driver fractions, 25 donors (and their twins) with the lowest driver fractions, 25 donors with high driver fractions in the buccals for known clonal haematopoiesis drivers (*TET2*, *SF3B1*, *DNMT3A*), and 5 donors with high driver fractions in the buccal swabs for each of the following drivers: *PPM1D*, *ASXL1* and *NOTCH3*. The remaining twin pairs were sampled randomly.

### Metadata

Metadata was provided by TwinsUK, obtained through periodical questionnaires that were collected longitudinally for most donors. For each participant, TwinsUK provided age, sex, height, weight, BMI, twin zygosity, and ethnicity. A few self-reported zygosities were corrected based on genotyping information. Self-reported medication histories were also obtained from questionnaires, however these are expected to be incomplete. Additional information on a short list of pre-specified treatments was provided by TwinsUK from anonymised medical records: metformin, tamoxifen, rapamycin, aspirin, non-steroidal anti-inflammatories, immunosuppressants, and history of herpes labialis. Cancer history was provided and coded as: 0 (no cancer), 1 (non-melanomatous skin cancer), 2 (other cancer), and 3 (oral cancer).

For major oral cancer risk factors and other relevant variables, we processed available questionnaires further to obtain summary metrics, including: tobacco smoking, alcohol consumption, physical activity, weight, height, BMI, oral hygiene, gastro-oesophageal reflux, diabetes, history of cancer, and medication histories.

#### Smoking and alcohol consumption

Self-reported smoking and alcohol consumption was collated from 14 periodical questionnaires. We focused on the most recent questionnaires due to the relevance of the questions asked in them and the coverage of answers across individuals. For smoking, we kept the maximum value of reported pack-years per donor across questionnaires. As standard, one pack-year was defined as 365 packs of cigarettes (7,300 cigarettes). For alcohol intake, self-reported current weekly consumption was available for most donors, but self-reported information on lifetime consumption was only available for a minority of donors. An estimate of drink-years was calculated by multiplying the average current weekly alcohol consumption, across multiple questionnaires if available, by the duration of adult life (age – 18). We note that this estimate is an extrapolation and should be used with caution, but regression models suggest that this estimate was more explanatory than self-reported lifetime consumption (see **Supplementary Note 7** for analyses on alternative metrics).

#### Oral health

Self-reported information on gingivitis, periodontitis, and gum bleeding was only available for a minority of donors. In contrast, the number of natural teeth remaining was available for most donors, recorded as an ordinal variable. For ease of interpretation in the regression models, we inverted this variable to reflect the number of missing teeth, as follows: 0=20 or more natural teeth, 1=10-19, 2=1-9, and 3=no natural teeth. Where multiple answers were available from questionnaires on different years, the lowest number of natural teeth left was used.

#### BMI, weight, and height

Weight and height were provided by TwinsUK for most donors. Both metrics were averaged across questionnaires for each donor. BMI was calculated using the standard formula: weight/(height^2^).

### Buccal swab processing and sequencing

Puritan buccal swab kits with instructions for self-collection were posted to the homes of voluntary donors by TwinsUK (CamBio, catalogue ID CA-1723-H100). Kits contained a primary and secondary plastic container, an outer rigid container (Alpha Laboratories, catalogue ID RF95-LL1) and a prepaid return envelope. Participants mailed their buccal swabs directly to the Wellcome Sanger Institute. Swabs were refrigerated at 4℃ upon arrival.

To extract DNA, buccal cells were dissociated into 1 mL PBS solution in an Eppendorf tube through manual agitation for one minute. The swab tip was then cut with scissors and left in the tube for 30 minutes, before removing it. The solution was then centrifuged at 1,000 × *g* for one minute. The supernatant was removed leaving a cell pellet with minimal residual PBS (<100 μL). The QIAGEN QIAmp DNA Micro Kit (catalogue ID 56304) was used for cell lysis and DNA extraction. First, 180 μL buffer ATL and 20 μL Proteinase K were added to the resuspended cell pellet, followed by overnight incubation on a thermomixer at 56℃ and 800 rpm. DNA extraction followed the manufacturer’s protocol with several modifications: centrifugation steps were all performed at 20,000 × *g* , the elution buffer and volume was 50 μL buffer EB (10 mM Tris-Cl, pH 8.5, catalogue ID 19086), incubation with the first elution step was for five minutes, and the eluent was passed through the spin column for a repeat elution into an Eppendorf Lo-Bind 1.5 mL tube (catalogue ID 0030108051). The final volume of extracted DNA was stored at -20℃. 40 μL of the thawed volume was diluted to 120 μL with buffer EB (catalogue ID 19086) and submitted for NanoSeq library preparation on an Abgene AB0800G plate (catalogue ID AB0800G, Thermo Fisher Scientific).

A detailed description of the targeted NanoSeq and standard duplex sequencing library preparation protocols is provided in **Supplementary Note 1**.

### Mutation calling

Sequencing data was mapped to the human genome (build GRCh37+decoys) with BWA-mem ^66^ as described before ^11^. Bases were called when there was duplex consensus with at least two reads per original strand, requiring a minimum consensus base quality score of 60, a VAF lower than 0.1 in the matched normal, a minimum AS-XS of 10 (see below), no more than an average of three mismatches per read (or four if a variant is called), a minimum coverage of 25x in the matched normal, and trimming 8 bp from each read end. Instead of sequencing independent matched normals to filter out germline variation, we took advantage of the high coverage and polyclonality of the buccal swab samples to remove germline SNPs by filtering out variants with VAF≥10%. We note that this is adequate as long as the samples are highly polyclonal. Relaxing this cutoff to VAF≥30% did not seem to recover genuine mutations in the buccal swabs but led to an increase in mapping artefacts. Since all blood samples had matching buccal swab data, somatic mutations in blood were called using their buccal swabs as matched normals, excluding as likely germline any variants with VAF≥10% in the buccal swabs.

A significant modification in the targeted NanoSeq calling pipeline compared to our published whole-genome NanoSeq pipeline is the relaxation of the AS-XS threshold from 50 to 10. AS-XS measures the difference in mapping quality between the primary and secondary alignments, excluding regions with ambiguous mapping from analysis. For mutation burden and signature analyses with whole-genome NanoSeq, we previously recommended a strict AS-XS cutoff to minimise the impact of mapping artefacts ^11^. However, for driver discovery it is important to preserve regions with less unique mapping qualities. Using a list of 1,152 oncogenic hotspots from TCGA and MSKCC provided by the dNdScv package ^29^, we noticed that the original AS-XS cutoff would have filtered out a significant number of them. Reducing the AS-XS cutoff from 50 to 10 ensured the retention of duplex coverage on nearly all canonical cancer hotspots while still ensuring accurate mutation rates and signatures in control cord blood samples (**Extended Data Fig. 2**).

Two additional filters are important to avoid recurrent mapping artefacts and to minimise the effect of inter-individual contamination. First, a ‘SNP+noise’ mask containing common germline SNP sites and recurrent mapping artefacts was generated for targeted NanoSeq as described before ^11^. Second, we noticed that mapping errors not captured by this mask can manifest as recurrent artefacts where the mutant base is often seen at specific positions within a read. This can be caused, for example, by mismapping of reads from polymorphic segmental duplications. A Kolmogorov–Smirnov test on the position of the mutant bases within reads was applied to remove recurrent artefacts after mutation calling. Indels were also filtered out if their overlap with the ‘SNP+noise’ mask was ≥50%, if they occurred at sites without a base called, if they had a VAF >0.1, or if they were seen in >50 samples. This only removed a small number of artefactual indel sites, which also had a strong read positional bias.

### Duplex VAFs and unbiased VAFs

The VAF represents the proportion of reads at a specific site carrying a variant, relative to the total reads at that site. When working with standard duplex sequencing or targeted NanoSeq data, only a fraction of read bundles reach the ‘2+2’ requirement for duplex calling (i.e. read families with at least two reads from both strands). We can then calculate three separate VAFs: (1) the “duplex VAF”, defined as the fraction of callable (2+2) read bundles supporting a given mutation, (2) the “BAM VAF”, calculated using the deduplicated BAM file containing one representative read per read bundle (and including calling and non-calling read bundles), (3) the “unbiased BAM VAF”, calculated using the deduplicated BAM file but excluding calling read bundles.

These VAFs can be used for different purposes. (1) Estimation of the fraction of cells in a sample carrying a specific mutation. If a mutation was discovered in a sample using duplex (2+2) reads, duplex VAFs or BAM VAFs tend to overestimate the fraction of cells carrying the mutation in the sample due to the discovery bias resulting from the inclusion of reads used for mutation calling. For this purpose, “unbiased BAM VAFs” provide an unbiased estimate of the VAF of a mutation in the sample as they are calculated from reads not used for duplex calling. (2) Estimation of the fraction of cells carrying somatic mutations in a given gene. The molecules that reach duplex calling (2+2) in a targeted NanoSeq experiment represent a random sample of all copies of a gene in a population of cells. The duplex VAF for a given site represents the fraction of mutant molecules at the site. If we assume that all (or nearly all) cells are diploid and that cells carry at most one driver mutation per gene (heterozygous), then we can estimate the fraction of cells with mutations in a given gene by summing the duplex VAF (*v*_d_) of mutations across all sites in the gene (*F* = 2 *∑ v_d_*). If we assume that cells may carry up to two mutant copies of the gene per cell or if we are looking at a haploid region of the genome (e.g. X chromosome in males), we can estimate the fraction of mutant cells in the sample using the sum of duplex VAFs across all sites in the gene (*F* = *∑ v_d_*). Some genes, such as *NOTCH1* in squamous epithelia can show biallelic loss by one mutation in each allele (SNVs or indels) or by one mutation and a copy number change (either a deletion or a copy-neutral loss of heterozygosity). We have previously shown that for these conditions, as well as for populations with mixtures of heterozygous and homozygous mutant cells, the fraction of mutant cells in the population falls within the range [*∑ v_d_*, 2 *∑ v_d_*] ^3^. Unless described otherwise, other references to the fraction of mutant cells for a given gene assume a maximum of one driver mutation per cell and a largely diploid population.

Since not all non-synonymous mutations in a driver gene are driver mutations ^29^, to estimate the fractions of cells with driver mutations (**Fig. 1g and 2g**), we multiplied the estimated fraction of cells with non-synonymous mutations by the estimated fraction of mutations that are drivers for each class. We estimated the fraction of mutations that are drivers using (ω-1)/ω, for mutation classes with ω≥1 (where ω is the dN/dS ratio per mutation type per gene). To account for potential differences in clone sizes for driver mutations, we used dN/dS ratios calculated without collapsing mutations reported by multiple molecules into single entries to dNdScv (**Supplementary Code**).

### Epithelial purity and targeted methylation

To quantify the epithelial fraction of a representative set of buccal swabs, we used two approaches: (1) targeted enzymatic methylation sequencing on 187 buccal swabs, and (2) comparing the VAFs of clonal haematopoiesis mutations in the buccal swabs of donors with blood and buccal swab data.

From 187 swabs, we generated low-input enzymatic methylation libraries and then undertook targeted capture with a panel of informative CpG sites, using NEB’s EM-seq kit (NEBNext Enzymatic Methyl-seq Kit NEB #E7120L). We used a custom Twist Bioscience hybridisation panel targeting 1162 CpGs selected from the centEpiFibFatIC.m, centDHSbloodDMC.m and centEpiFibIC.m matrices in the EpiDISH R package ^67^, to deconvolute epithelial, fibroblast, fat and blood cell types. We also targeted 353 CpG from the original Horvath clock ^68^ and 50 CpGs in the promoters of 25 driver genes. The design is available in **Extended Data Table 2**.

For each sample, DNA was quantified and normalised to ∼1 ng/μl. Normalised DNA samples were then sheared with NEB’s Ultrashear fragmentation mix (NEBNext UltraShear NEB #M7634L), end-repaired, A-tailed, adapter-ligated with a methylated TruSeq-compatible adapter stub (all using NEB Ultra II reagents) and, after a SPRI clean-up, the resulting libraries were oxidised using TET2 (converting methylcytosines to carboxylcytosines) and deaminated using APOBEC (converting bare cytosines to uracils but retaining the carboxylcytosines, thus preserving the locations of methylation marks). The deaminated libraries were amplified, and sequencing indexes (and the rest of the adapter sequence) were introduced using NEB Q5U and the Sanger Institute’s UDI primers. After a further SPRI clean-up, libraries were requantified and mixed in an equimolar pool with a cumulative DNA mass of 1-4 μg. Twist Bioscience probes targeting the sequences of interest were then added. After evaporating all the liquid, the probes hybridised to the DNA and the targets were pulled down and cleaned up (using Twist fast hybridisation reagents and Thermo DynaBeads MyOne streptavidin-coupled beads). After a final PCR amplification (KAPA HiFi) and SPRI clean-up, a pool of all samples was QCed by Agilent Bioanalyser and sequenced in a single S4 lane of Illumina NovaSeq 6000.

Epithelial, fibroblast and blood cell fractions were estimated using EpiDISH and hEpiDISH ^67^. The latter allows hierarchical deconvolution, first relying on centEpiFibIC to estimate epithelial, fibroblast and blood fractions, and applying centDHSbloodDMC to deconvolute the different types of blood cell types. The median epithelial fraction across all 187 swabs was 95.1% (**Extended Data Fig. 1h**). Most of the non-epithelial cells were neutrophils, likely a result of saliva contamination of the buccal swabs.

As a complementary analysis of blood contamination in the buccal swab samples, we compared the VAF of blood mutations in buccal swabs. To do so, we used 43 pairs of buccal and archival blood samples where the date of collection of the blood sample was within 3 years of the buccal swab, and which contained at least one large clone in blood (VAF ≥1%). The median of the ratio of buccal VAF to blood VAF for 58 blood mutations that met these criteria was 0.076, which provides an alternative estimate of the median blood contamination in these samples around 7-8%.

### Removal of DNA contamination

The ability of NanoSeq to detect somatic mutations in single molecules of DNA makes it particularly sensitive to DNA contamination, either from other humans (calling germline SNPs from the contaminant individual as somatic mutations in the affected sample) or from other species with sufficient conservation to map to the human genome (which is more likely in targeted NanoSeq due to the higher conservation of coding regions).

#### Human DNA contamination

We have previously shown ^11^ that when analysing whole-genome NanoSeq data, the percentage of contaminating DNA can be estimated using verifybamID ^69^. However, we found verifybamID to be unreliable for targeted NanoSeq data. To qualitatively detect human DNA contamination on targeted NanoSeq data, a useful metric is the fraction of all substitutions filtered by the ‘SNP+noise’ mask. Although useful, this metric may not be reliable for samples with low duplex coverage and few mutations. As a complementary approach, we genotyped common SNPs in targeted regions to identify homozygous alternative (non-reference) SNPs. Presence of reference bases at these sites is indicative of contamination. Whereas this is not a direct estimate of the percentage of contamination given the difficulty of determining the genotype of the contaminant at those alternative homozygous SNPs, it can serve as a sensitive indicator of inter-individual DNA contamination.

We called SNPs with bcftools ^70^ using the following commands: bcftools mpileup --max-depth 20000 -Ou -f $genome $bam | bcftools call --ploidy GRCh37 -mv -Ob -o BCFTOOLS/$OUT_PREFIX.calls.bcf; bcftools view -i ‘%QUAL>=100’ BCFTOOLS/$OUT_PREFIX.calls.bcf > BCFTOOLS/$OUT_PREFIX.calls.filtered.vcf

For the assessment of contamination, we restricted the analysis to SNPs overlapping both our SNP mask and our targeted panel. We used bam2R (from the deepSNV R package) ^71^ to obtain the number of reads supporting the alternative and reference alleles, and kept SNPs with a mean coverage across samples >200×. For each SNP in each sample, the genotype was set to ‘NA’ if the coverage was <20×, to alternative homozygous (1/1) if the VAF was >0.8, to heterozygous (0/1) if the VAF was between 0.3 and 0.7, and to reference homozygous (0/0) if the VAF was <0.1. Finally, we only kept SNPs seen in ≥2 samples and in <1,000 samples. For each homozygous SNP, we calculated the reference fraction and we report the median across all homozygous SNPs in the sample at hand. 17 samples with a median reference-base VAF >0.01 at non-reference homozygous SNP sites were considered contaminated and excluded from analysis (**Extended Data Fig. 1d**).

#### Cross-species contamination

Donors were requested to rinse their mouths before buccal swab collection to minimise non-human DNA contamination from food or bacteria, but some samples showed evidence of it. This can lead to mismapping of non-human DNA reads to the human genome, detectable as an excess of clustered synonymous mutations. To systematically identify these samples we used Kraken v2 ^72^, using 1 million unmapped reads per swab and a database of potential sources of contamination able to map to the human genome: *Mus musculus*, *Bos taurus*, *Ovis aries*, *Sus scrofa*, *Equus caballus*, *Oryctolagus cuniculus*, *Meleagris gallopavo*, and *Gallus gallus*. Bacterial contamination should not be a problem given their sequence divergence from the human genome. In addition, for each sample we calculated the global dN/dS ratio across passenger genes, and compared the contamination fractions estimated with Kraken to the observed dN/dS ratios. dN/dS ratios decrease with non-human contamination because of evolutionary conservation of non-synonymous sites. Based on the impact of contamination on dN/dS ratios (**Extended Data Fig. 1e**), we excluded from further analyses 132 samples with >0.25% of non-human unmapped reads.

### HPV detection and characterisation

The genome sequence of 19 HPV types considered high-risk ^73^ were retrieved from GenBank. We built a multiple sequence alignment of these genomes with MAFFT ^74^ using Jalview ^75^. Based on conservation across these highly divergent HPV strains, we retained ∼3000 bp for each of the strains to design HPV-specific probes that we included in our Twist target gene panel. The GenBank accession numbers of the 19 selected HPV types were: KU298887.1, KU298893.1, KU298928.1, KX514417.1, KX514421.1, KX514431.1, KY225967.1, LR862061.1, LR862064.1, LR862079.1, MT218010.1, MT783412.1, MT783416.1, MT783417.1, MZ374448.1, MZ 509108.1, NC_001357.1, NC_001526.4, NC_001583.

Once our targeted sequencing data was mapped to the human genome we retrieved the unmapped reads and remapped them to the genomes of the 19 HPV strains using BWA-mem ^66^. Mapping results were reviewed manually to distinguish between unreliable mappings (very repetitive, low-complexity, soft-clipped reads) and likely true HPV sequences. For ambiguous cases, we searched the mapped read with BLAST against the NCBI’s non-redundant nucleotide database. This allowed us to identify some hits to HPV strains not originally covered in our panel.

We detected HPV in 12 samples, in some cases supported by thousands of reads while in others by as little as one single read. Multiple HPV strains were detected. The following (anonymised) list of donors show the results: X1 donor (HPV 16, 44 reads), X2 (HPV 53, 6 reads), X3 (HPV 33 and HPV 58, 20 and 5 reads), X4 (HPV 33, 115 reads), X5 (HPV 53, 6667 reads), X6 (HPV 59, 12 reads), X6 (HPV 56, 707 reads), X7 (HPV 51, 236 reads), X8 (HPV 56, 1 read), X9 (HPV 21 not in panel, 2 reads), X10 (HPV 24 not in panel, 1 read), X11 (HPV 30, not in panel, 1 read), X12 (HPV 33, 6 reads).

Given that only 12/1,042 samples had detectable HPV presence using the targeted capture and that this is not a validated assay for HPV detection, we were unable to study the impact of HPV on the mutation and selection landscape in the oral epithelium, which remains an important question for future studies. Instead, we excluded these 12 samples from the epidemiological regression analyses to reduce the risk of confounding effects.

### Germline genotyping

#### Genotyping array data

Pre-existing array genotyping data from TwinsUK was used for GWAS and other analyses. The samples had been genotyped with the following arrays: HumanHap300, HumanHap610Q, 1M-Duo and 1.2MDuo 1M. Following genotype calling, some samples were excluded from analyses involving genotyping data based on different criteria: a sample call rate <98%, heterozygosity across all SNPs ≥2 standard deviations from the sample mean, evidence of non-European ancestry as assessed by PCA comparison with HapMap3 populations, observed pairwise Identity By Descent (IBD) probabilities suggestive of sample identity errors. We also used IBD probabilities to correct misclassified zygosity. We then excluded SNPs using the following criteria: Hardy-Weinberg *p*-value <10^−6^, assessed in a set of unrelated samples; minor allele frequency (MAF) <1%, assessed in a set of unrelated samples; SNP call rate <97% (SNPs with MAF ≥5%) or <99% (for 1%≤MAF<5%). Following genotype and sample filtering, the data were imputed using the Haplotype Reference Consortium reference panel and SNPs with an imputation *R2* <0.5 were excluded.

#### Germline genotyping from sequencing data

For analyses relying on common SNPs, we called SNPs using bcftools as described in the DNA contamination section. For analyses relying on both common and rare SNPs we run GATK’s HaplotypeCaller (v4.0.1.2) ^76^, using default options, setting ploidy to 2 except for male chromosome X (haploid), and providing dbSNP v141 (<u>ncbi.nlm.nih.gov/snp</u>) (PMID:10447503) for annotation of the calls. The resulting VCF files were intersected with our panel regions using bedtools ^77^ and missing genotypes were annotated as REF with bcftools +missing2ref ^70^, on the basis of the high coverage available.

### Selection analyses

A detailed description of the methods used to analyse positive and negative selection in this study is provided in **Supplementary Note 2**. This includes a description of the new one-sided tests in dNdScv, the use of duplex coverage correction in dNdScv, estimates of the number of driver mutations in the dataset, and a description of dN/dS analyses at the level of single sites and groups of functionally related sites within genes.

### Mutation burdens and signatures

Mutation burden is defined as the number of mutations per bp in a given region, and it is calculated in NanoSeq data as the number of mutant bases divided by the total number of bases sequenced with duplex information. Estimating mutation rates from targeted data can have several challenges. First, the mutation burden of a given region of the genome will be affected by its sequence composition. We can remove this confounding effect by correcting mutation burdens by the trinucleotide frequencies of a targeted region (relative to the whole genome) and the mutability of each trinucleotide, as described before ^11^. Whereas this corrects for the effect of different sequence composition, it does not correct for a systematic difference in the mutability of different regions, such as genic and intergenic sequences, as explained in the text. Whole-genome NanoSeq can be used for an unbiased genome-wide measurement of mutation burdens (e.g. **Extended Data Fig. 4**, **Supplementary Note 3**). Second, when estimating mutation rates from gene sequences, particularly from panels of positively selected genes, positive selection can lead to an inflation of the apparent mutation rate. To avoid this, the mutation burdens described in this manuscript were estimated only from passenger genes. Synonymous sites can also be used as a proxy for the neutral mutation rate, as described before ^3^. Finally, mutation burdens estimated from targeted regions can be inflated or deflated by the undue influence of one or a few large clones. For example, if a sample is dominated by a large clone, the presence of a passenger mutation in the clone overlapping the target region would lead to an overestimation of the mutation burden, whereas the absence of any mutation in the clone in the target region would lead to a modest underestimate of the burden. This is apparent in the targeted NanoSeq data for blood (**Fig. 1e**), where a few samples show inflated burden estimates due to high VAF passenger mutations. Some duplex sequencing studies avoid this by counting each mutant site only once, but this leads to a systematic underestimation of mutation burdens, leading to lower bound estimates of the mutation burden. Instead, for the targeted NanoSeq blood data, we calculated the confidence intervals for the mutation burden using Poisson bootstrapping of the mutant sites, resulting in wider confidence intervals when one or a few sites had an undue influence in the burden estimate. In general, burden estimates from targeted NanoSeq are expected to be most reliable when working with highly polyclonal samples or when the size of the panel is considerably larger than the inverse of the mutation rate per bp.

We inferred mutational signatures of single-base substitutions (SBS) using the sigfit (v2.1.0) R package ^78^. Genome strand information for each target gene was used to produce transcriptional-strand-wise (TSW) trinucleotide mutation catalogues (192 mutation categories) for mutations within genes, using the *build_catalogues* function in sigfit. Inference was performed for a range of signature numbers (*N*=2,…,5), using the TSW mutation counts from 92 oral epithelium samples having ≥500 mutations each. To account for variation in sequence composition, observed mutation opportunities (trinucleotide frequencies based on the NanoSeq coverage per site for each sample) were supplied to the *extract_signatures* function. Mutation opportunities were assumed to be equal between the transcribed and untranscribed strands. The best-supported number of signatures, on the basis of overall goodness-of-fit and consistency with known COSMIC signatures (v3.0; <u>cancer.sanger.ac.uk/signatures</u>), was found to be *N*=2. Of the two inferred signatures, Signature A corresponded to a combination of COSMIC signatures SBS1 (6%) and SBS5 (94%) (cosine similarity 0.90), while Signature B (SigB) was highly similar to COSMIC SBS16 (cosine similarity 0.97). To estimate the contribution of both signatures to all epithelium samples, these two signatures were fitted to the TSW mutation counts for each sample using the *fit_signatures* function. Signature burdens (mutations per diploid genome attributed to each signature) were calculated by multiplying the signature exposure estimates by the whole-genome passenger mutation burden estimates for each sample. Prior to plotting using the *plot_spectrum* function, signatures were transformed to a genome-relative representation by scaling their probability values according to the corresponding whole-genome human trinucleotide frequencies, using the *convert_signatures* function.

A high rate of T>C mutations at ApT dinucleotides is common to the COSMIC SBS5 and SBS16 signatures. To explore whether these T>C mutations are caused by similar underlying processes, we studied the extended (pentanucleotide) sequence context of T>C mutations in several datasets. To do so, we obtained TSW pentanucleotide counts for T>C substitutions (256 mutation categories) applying a custom R function to mutations in the following sample sets: (i) matched blood samples (*n*=371); (ii) hepatocellular carcinoma (Liver-HCC) samples from the Pan-Cancer Analysis of Whole Genomes study ^79^ (downloaded from dcc.icgc.org/pcawg) for which signature fitting estimated a COSMIC SBS16 exposure >0.2 (*n*=4); (iii) oral epithelium samples with SigB exposure >0.25 (*n*=121); (iv) oral epithelium samples with SigB exposure <0.25 (*n*=921). Prior to plotting using custom R functions, pentanucleotide catalogues were transformed to a genome-relative representation by scaling mutation counts according to the corresponding whole-genome human pentanucleotide frequencies. The results of this analysis are described in **Extended Data Fig. 8** and **Supplementary Note 3**.

Mutation catalogues of double-base substitutions (DBS; 78 mutation categories) and indels (83 mutation categories) were produced for mutations in the following sample sets: (i) all oral epithelium samples (*n*=1,042); (ii) oral epithelium samples from heavy smoking non-drinking donors (*n*=27); (iii) oral epithelium samples from non-smoking non-drinking donors (*n*=224). DBS catalogues were produced using a custom R function, while indel catalogues were produced using the *indel.spectrum* function in the Indelwald tool (24/09/2021 version; github.com/MaximilianStammnitz/Indelwald). Although we attempted both *de novo* extraction and fitting of mutational signatures to the DBS and indel catalogues, mutation numbers were not large enough to allow inference of informative signatures or exposures. Mutation spectra for DBS and indels were plotted using the *plot_spectrum* function in sigfit. The results of this analysis are described in **Extended Data Fig. 8** and **Supplementary Note 3**.

### Regression analyses

To test for associations between epidemiological variables and rates of mutational signatures or driver mutation frequencies, we used mixed-effect regression models (*lmer* function in the lme4 R package; ^80^) as described below.

#### Outcome variables

For the analyses shown in the main text, we ran a separate regression model for each outcome variable: SNV burden, Signature A burden, Signature B burden, indel burden, dinucleotide burden, the sum of all driver frequencies in a sample, and the driver density per sample for eight major driver genes. To avoid excessive loss of statistical power due to multiple testing correction across all outcome variables and predictors, and to focus on the genes with the highest information content, we restricted the regression analyses to eight driver genes with >400 coding mutations in the dataset, and an estimated driver fraction ≥30% based on dN/dS ratios.

#### Predictor variables

For the analyses in the main text, we selected nine variables as predictors in multiple regression models, including major oral cancer risk factors as well as other potentially relevant variables: age, sex (F/M), pack-years, drink-years, type II diabetes (Y/N), body mass index, missing teeth, physical activity score (International Physical Activity Questionnaire [IPAQ] score), and cancer history (Y/N). The twin structure was modelled with a random effect. The R code used for these and supplementary regressions is provided in the **Supplementary Code**, but for illustrative purposes the structure was as follows: *lmer(snvburden ∼ age + sex + packyears + drinkyears + t2d + bmi + missingteeth + ipaq + cancer + (1|familyID), REML=F)*

Only samples with a mean duplex coverage across genes ≥200 dx and available metadata for all the predictor variables and the outcome variable in each multiple regression model were included for analyses. 12 samples with potential evidence of HPV reads (see above) and six samples with a self-reported history of chemotherapy were excluded from the regression. These variables may be expected to have mutagenic and/or selectogenic effects on the oral epithelium, but the number of affected donors was too low for a robust analysis in the current study.

*P*-values were calculated for each covariate in each multivariate regression model using a likelihood ratio test by comparing the likelihood of the full model with a model without each variable, using the *drop1* function in R. Multiple testing adjustment using the Benjamini–Hochberg procedure was then applied to all *p*-values in the main text analyses (126 tests: 14 predictors × 9 outcome variables).

### Additional regression models, GWAS, and heritability analyses

Additional regression analyses, including using extended medication data as predictors, interaction analyses between smoking and alcohol, and measures of selection (corrected for mutation rates) as outcome variables for the detection of selectogenic influences, are described in **Supplementary Note 7**. Methods and supplementary results for GWAS analyses and heritability tests are described in **Supplementary Note 8**.

