## Supplementary Materials for "Somatic mutation and selection at epidemiological scale"

### Supplementary Notes


- Supplementary Note 1. Targeted NanoSeq and duplex sequencing protocols
- Supplementary Note 2. Selection analyses on single-molecule sequencing data
- Supplementary Note 3. Whole-genome data and supplementary mutational signature analyses
- Supplementary Note 4. Further description of the blood and buccal driver landscapes
- Supplementary Note 5. Multistage models linking mutation rates and clonal expansions to cancer risk
- Supplementary Note 6. Simple models of clonal growth in blood and oral epithelium
- Supplementary Note 7. Additional regression models
- Supplementary Note 8. Heritability and GWAS


### Extended Data Figures


- Extended Data Figure 1. Description of the cohort
- Extended Data Figure 2. Targeted NanoSeq quality metrics
- Extended Data Figure 3. Further description of the blood driver landscape
- Extended Data Figure 4. Whole-genome NanoSeq on buccal swabs
- Extended Data Figure 5. Full buccal driver landscape
- Extended Data Figure 6. Distribution of mutations within selected buccal driver genes
- Extended Data Figure 7. Models of clonal growth
- Extended Data Figure 8. Mutational spectra of somatic SBSs, DBSs and indels
- Extended Data Figure 9. Additional regression models


### Extended Data Tables


- Extended Data Table 1. Gene panel
- Extended Data Table 2. Methylation panel
- Extended Data Table 3. dNdScv results
- Extended Data Table 4. Sitednds results
- Extended Data Table 5. Heritability results (GREML and ACE)

**Supplementary Note 1. Targeted NanoSeq and duplex sequencing protocols**

*Development of targeted NanoSeq*

Duplex sequencing (DuplexSeq) is an error correction strategy for next generation sequencing that tags individual molecules of DNA with random barcodes and uses the consensus of reads derived from both strands of the original DNA molecule to remove sequencing and PCR amplification errors. The error rates of duplex sequencing protocols depend on the specific protocol used and the amount of pre-existing DNA damage in the sample. Nanorate sequencing (NanoSeq) is a duplex sequencing protocol with several innovations to further reduce error rates to $<5\times10^{-9}$ errors/bp. As described before [1], key changes with respect to standard duplex sequencing include: (1) genome fragmentation that avoids the standard use of polymerases during end repair, thus reducing the copying of errors between strands, (2) use of ddBTPs to prevent nick extension during A-tailing, (3) a qPCR step to ensure optimal duplicate rates in any library, independent of DNA input, (4) key filters against alignment errors (particularly the AS-XS filter against ambiguous mapping) and DNA contamination. In the original version of NanoSeq [1], blunt-end restriction enzymes were used for genome fragmentation without end repair, which ensured error rates $<5\times10^{-9}$ errors/bp but led to partial (~30%) coverage of the genome. In the current study, we introduce two alternative genome fragmentation methods that provide full genome coverage whilst retaining the original error rates.

The potential to use sonication and exonuclease digestion in NanoSeq was briefly introduced in our original NanoSeq publication [1]. However, the original proof-of-principle example protocol had very low library yields, approximately 2-10% of those obtained with the restriction enzyme method. We have carried out extensive R&D work to increase the yield of this approach. Amongst other changes detailed above, the mung bean nuclease concentration was increased to 5 units/reaction. The phosphorylation and A-tailing reactions were combined into a single reaction and the units of each enzyme were increased. T4 DNA ligase was substituted with NEB Ultra II and the adapter concentration was increased. Background noise in the qPCR reaction, resulting from adapter dimers, was reduced by diluting the ligation reaction prior to stringent SPRI clean-up. Yields (fmol/ng) for the sonication NanoSeq protocol introduced in the current study are now on par and often greater than those of the original restriction enzyme method.

With the aim of further increasing yields, we developed enzymatic targeted NanoSeq. This utilises NEB UltraShear for DNA fragmentation, an enzymatic fragmentation mix that is formulated specifically to avoid error introduction during fragmentation. To ensure that the low error-rates of NanoSeq are achieved, we required further optimisation of the fragmentation conditions in alternative buffers. NEB Buffer r1.1 with NAD+ supplementation was determined to be the most suitable substitute, based on fragment size distribution and enzyme activity. For formalin-fixed samples, fmol/ng yield was found to be greater with the enzymatic approach than with sonication, although considerably lower than unfixed samples. Yield increases were less pronounced (~1.7×) in less damaged samples (e.g. frozen or PAXgene-fixed). However, we note that the current enzymatic protocol has some drawbacks, including a considerable percentage of improperly paired reads, seemingly caused by ligation of DNA fragments in the library leading to chimeric molecules. Development work is ongoing to optimise blunting and increase the percentage of properly paired reads. Choice of method must thus take into consideration the requirement for higher yields as well as the ability to tolerate improperly paired reads. We also note that whereas the avoidance of end repair and the use of ddBTPs ensures a much lower error rate of NanoSeq than standard duplex sequencing, strand dropouts can be higher, particularly in damaged DNA samples.

*Targeted NanoSeq and duplex sequencing library preparation*

A summary of the standard duplex sequencing and targeted NanoSeq protocols used in **Fig. 1** is provided below. For these analyses, 30 ng of cord blood DNA and 50 ng of pancreas DNA were used per library. Bioinformatic analyses were identical for all protocols. All targeted NanoSeq data for buccal swabs and blood presented in the manuscript were generated using the sonication version of NanoSeq.

*Sonication DuplexSeq*

120 µL DNA was sheared using a Covaris ultrasonicator targeting a 450-bp insert size. DNA was purified using 300 µL AmpureXP beads (Beckman Coulter: A63882) and eluted in 51 µL 1× TE buffer. End repair was performed by adding 3 µL NEBNext Ultra II End Prep Enzyme Mix (NEB E7546S) and 7 µL NEBNext Ultra II End Prep Reaction Buffer (NEB E7546S) to 50 µL DNA. The reaction was incubated at 20 °C for 30 minutes, followed by 65 °C for 30 minutes (hold at 4 °C), with the lid temperature set to 75 °C. Ligation mix consisting of 30 µL NEBNext Ultra II Ligation Master Mix (NEB E7595L), 1 µL NEBNext Ligation Enhancer (NEB E7595L), 1.25 µL NFW and 1.25 µL xGen CS Adapter (IDT 1080799) was added to the A-tailing reaction and ligation was performed by incubating at 20 °C for 15 minutes with the thermocycler lid temperature turned off. DNA was purified by adding 60.76 µL AmpureXP beads (Beckman Coulter: A63882) and eluted in 31 µL nuclease-free water (NFW).

*Enzymatic DuplexSeq*

DNA was concentrated by performing a 2.5× AmpureXP (Beckman Coulter: A63882) bead clean-up. DNA was eluted in 26 µL 1× TE buffer. Fragmentation was performed by adding 14 µL NEBNext UltraShear Reaction Buffer (M7634L) and 4 µL NEBNext UltraShear Enzyme (M7634L). The reaction was incubated at 37 °C for 20 minutes, followed by 65 °C for 15 minutes (hold at 4 °C), with the lid temperature set to 75 °C. A-tailing was performed by adding 2 µL 500 mM DTT and 3 µL NEBNext Ultra II End Prep Enzyme Mix (NEB E7546). The reaction was incubated at 20 °C for 30 minutes, followed by 65 °C for 30 minutes (hold at 4 °C), with the lid temperature set to 75 °C. Ligation was performed by adding 30 µL NEBNext Ultra II Ligation Master Mix (NEB E7595L), 1 µL NEBNext Ligation Enhancer (NEB E7595L), 12.25 µL NFW and 1.25 µL xGen CS Adapter (IDT 1080799) to the A-tailing reaction. Ligation was performed by incubating at 20 °C for 15 minutes (hold at 4 °C) with the thermocycler lid temperature turned off. DNA was purified by adding 60.76 µL AmpureXP beads (Beckman Coulter: A63882). DNA was eluted in 31 µL NFW.

*Sonication NanoSeq*

120 µL DNA was sheared using a Covaris ultrasonicator aiming for a 450 bp target insert size. DNA was purified using 96 µL AmpureXP beads (Beckman Coulter: A63882) and eluted in 26 µL NFW. 25 µL DNA was taken into the end repair reaction; consisting of 3 µL 10× Mung Bean Nuclease Buffer (TAKARA 2420A), 1.875 µL NFW and 0.125 µL Mung Bean Nuclease (TAKARA 2420A). End repair was performed by incubating at 37 °C for 10 minutes with the thermocycler lid tracking 5 °C above the reaction temperature. DNA was purified by adding 75 µL AmpureXP beads (Beckman Coulter: A63882) and eluted in 11 µL NFW. 10 µL DNA was taken into the A-tailing reaction, consisting of 1.5 µL T4 DNA Ligase Buffer (NEB B0202S), 1.5 µL 1 mM equimolar dATP/ddBTP (NEB N0440S; MERCK Life Science 3732738001), 1.5 µL Klenow fragment (3′ to 5′ exo-, NEB M0212L) and 0.5 µL T4 Polynucleotide Kinase (NEB, M0201L). A-tailing was performed by incubating at 37 °C for 30 minutes with the thermocycler lid tracking 15 °C above the reaction temperature. Ligation mix consisting of 30 µL NEBNext Ultra II Ligation Master Mix (NEB E7595L), 1 µL NEBNext Ligation Enhancer (NEB E7595L), 12.75 µL NFW and 1.25 µL xGen CS Adapter (IDT 1080799) was added to the A-tailing reaction and ligation was performed by incubating at 20 °C for 20 minutes with the thermocycler lid temperature turned off. DNA was purified by adding 60 µL NFW and 78 µL AmpureXP beads (Beckman Coulter: A63882). DNA was eluted in 31 µL NFW.

*Enzymatic NanoSeq*

DNA was concentrated by performing a 2.5× AmpureXP (Beckman Coulter: A63882) bead clean-up. DNA was eluted in 24.6 µL 1× TE buffer. Fragmentation was performed by adding 14 µL NEBuffer r1.1 (NEB B7030S), 4 µL NEBNext UltraShear Enzyme (M7634L) and 1.4 µL 50 mM NAD+ (NEB B9007S). The reaction was incubated at 46 °C for 40 minutes, followed by 65 °C for 15 minutes (hold at 4 °C), with the lid temperature set to 75 °C. DNA was cleaned up by performing a 2.5× AmpureXP (Beckman Coulter A63882) bead clean-up. Beads were resuspended in 10 µL water. 10 µL resuspended beads were taken into the A-tailing reaction, consisting of 1.5 µL T4 DNA Ligase Buffer (NEB B0202S), 1.5 µL 1 mM equimolar dATP/ddBTP (NEB N0440S; MERCK Life Science 3732738001), 1.5 µL Klenow fragment (3′ to 5′ exo-, NEB M0212L) and 0.5 µL T4 Polynucleotide Kinase (NEB M0201L). A-tailing was performed by incubating at 37 °C for 30 minutes, followed by 65 °C for 30 minutes (hold at 4 °C) with the thermocycler lid tracking 15 °C above the reaction temperature. Ligation mix consisting of 30 µL NEBNext Ultra II Ligation Master Mix (NEB E7595L), 1 µL NEBNext Ligation Enhancer (NEB E7595L), 12.75 µL NFW and 1.25 µL xGen CS Adapter (IDT 1080799) was added to the A-tailing reaction and ligation was performed by incubating at 20°C for 60 minutes with the thermocycler lid temperature tuned off. DNA was purified by adding 60 µL NFW and 78 µL AmpureXP beads (Beckman Coulter A63882). DNA was eluted in 31 µL NFW.

NB. For the data in **Fig. 1**, $NAD^+$ was omitted from the fragmentation reaction and replaced with 1.4 µL 1× TE buffer. We recommend adding $NAD^+$ for optimal performance.

*Sequencing library quantification*

Library size was determined by running each sample on a Tapestation D5000 tape (Agilent 5067-5588) and by doing a region analysis spanning 150-3500 bp. DNA was quantified by qPCR using a KAPA library quantification kit. The supplied primer premix was first added to the supplied KAPA SYBR FAST master mix. In addition, 20 µL of 100 µM NanoqPCR1 primer (HPLC, 5′-ACACTCTTTCCCTACACGAC-3′) and 20 µL of 100 µM NanoqPCR2 primer (HPLC, 5′-GTGACTGGAGTTCAGACGTG-3′) were added to the KAPA SYBR FAST master mix. Samples were diluted 1:500 using NFW and reactions were set up in a 10 µL reaction volume (6 µL master mix, 2 µL sample/standard, 2 µL water) in a 384 well plate. Samples were run on the Roche 480 Lightcycler and analysed using absolute quantification (second derivative maximum method) with the high sensitivity algorithm. The concentration (nM [fmol/µL]) was determined as follows: mean of sample concentration × dilution factor (500) × 452/library size/1,000 (where 452 is the size of the standard in bp), and multiplied by an adjustment factor of 1.5. Samples were diluted to the desired fmol amount in 25 µL using NFW.

NB. For routine use of the NanoSeq sonication protocol, we do not normally evaluate the size of each library. We use 573 bp as the average molecule size.

*Library bottleneck and sequencing efficiency*

To maximise the efficiency and cost effectiveness of targeted NanoSeq and other duplex sequencing methods, it is important to optimise duplicate rates to maximise the number of read bundles (defined as a family of PCR duplicates) with at least two duplicate reads from each original strand. Sequencing a library too deeply (resulting in excessive duplicate rates) leads to unnecessarily deep sequencing of fewer read bundles, whereas sequencing too shallowly (insufficient duplicate rates) leads to many read bundles not reaching the ≥2 reads per strand required for base calling.

As previously described [1], to ensure optimal duplicate rates in all samples, we optimised the amount of sequencing reads that we devote per fmol of library, theoretically and empirically. If we assume negligible PCR biases, the number of reads expected per read family can be modelled with a zero-truncated Poisson distribution. Let $r$ (sequence ratio) be the ratio between the number of sequencing reads and the number of amplifiable molecules in the library. The mean number of reads per read family, $m$, can be estimated as the mean of a zero-truncated Poisson distribution (with $\lambda = r$): $m = \frac{r}{1-e^{-r}}$. We can then estimate the duplicate rate of a library, $d$ (defined as the fraction of reads that are duplicate copies, in practice identified as reads with the same barcode and mapping coordinates), as follows: $d =$

$\frac{m-1}{m} = 1 - \frac{1}{m} = 1 - \frac{1-e^{-r}}{r}$. We can then define the efficiency of a targeted NanoSeq library (or a duplex sequencing library), $E$, as the ratio between the duplex coverage (the number of base pairs with ≥2 reads per strand) and the raw sequencing coverage (the number of base pairs sequenced). This can be modelled as: $E = \frac{P(x \geq 2|r/2)^2}{r}$. Here, the numerator is the probability of a read bundle having two or more reads from both strands (i.e. the probability of being a usable duplex bundle), where $P$ is the Poisson probability, and the denominator is the average sequence investment in each molecule of the original library. Using this equation, we find that the optimal efficiency of targeted NanoSeq (which maximises the duplex coverage for a given amount of raw sequencing) is achieved for $r \approx 6.4$ and $d$ (duplicate rate) ≈ 84%, and that 80% of the maximum efficiency is obtained in the range $r \approx$ 3.9-11, and $d \approx$ 75-91%. Having estimated the theoretical optimal duplicate rates for NanoSeq, we then used a serial dilution experiment to obtain an empirical efficiency curve [1]. This suggested that empirical duplicate rates ~75-85% maximise cost efficiency, which is also supported by the empirical efficiencies and duplicate rates obtained from the buccal swab samples in the study (**Extended Data Fig. 2a-c**).

Knowing the concentration of a NanoSeq library in fmol/μL (measured using a qPCR reaction on a small aliquot of the unamplified library, as described below) and the desired amount of sequencing (number of read pairs), we can use $r_{opt}$ to calculate the volume of library that we need to take forward to amplification and sequencing, as follows: $fmol_{opt} = \frac{N}{f \cdot r_{opt}}$. Here, $f$ refers to the number of amplifiable DNA fragments per fmol in the library. To determine the value of $f$ for our protocol and qPCR machines, we originally carried out a serial dilution experiment, and compared the empirical values of library complexity (estimated using the Picard software; [2]) and the input fmols into sequencing [1], resulting in an estimate of $f = 10^8 - 3\times10^8$ sequenceable fragments per fmol. Using this equation, we can ensure optimal duplicate rates for any library, independently of the input DNA from each sample. For example, for a typical whole-genome NanoSeq experiment we often use 150 million read pairs per sample (equivalent to ~15× raw human genome coverage using 150-bp reads). With $r_{opt} \approx 6.4$ and $f \approx 10^8$ (based on [1], for the restriction enzyme NanoSeq protocol) this translates into $fmol_{opt} \approx 0.23$ (the estimated optimal fmol input per sample for amplification and sequencing).

Conversely, if we want to sequence the entire available library, we can use the same equation to calculate the amount of sequencing required to ensure optimal duplicate rates, which was the approach used for the sequencing of buccal swabs, to maximise the duplex coverage per sample. When performing targeted or whole-exome capture, the amount of sequencing needed for a given sample needs to consider the size of the regions targeted and the capture efficiency (the fraction of reads on-target for a given panel). We can use the equation below for this ($p$ being the panel size, $g$ being the genome size, and $c$ being the on-target capture fraction):

$$N = \frac{fmol \cdot r_{opt} \cdot f \cdot p}{g \cdot c}$$

Using $r_{opt} \approx 6.4$, $f \approx 2.6\times10^8$ (based on a calibration experiment for the sonication protocol), and $c$ = 0.70 (**Extended Data Fig. 2a**), this means that 750 Gb of sequencing, or 2.5 billion 150-bp read pairs (a common output for one S4 lane of Illumina NovaSeq 6000), should provide approximately optimal duplicate rates for ~90 fmol of library captured with a whole-exome panel of 35 Mb, or for ~3500 fmol of library captured with a targeted gene panel of 0.9 Mb. To apply targeted NanoSeq on other panels, we recommend using these equations to estimate the optimal amount of sequencing per fmol, and adjust the fmol/sequencing ratio based on performance.

In practice, in the current study we multiplexed libraries aiming for ~4000 fmol per S4 lane for the targeted gene panel and ~85 fmol per lane for the exome panel, which typically yielded ~12,000 dx and ~250 dx per lane, respectively. These yields can vary modestly as a function of strand dropout (affected by the amount of unamplifiable DNA lesions in the input DNA, as well as PCR biases; **Extended Data Fig. 2b,c**) and the actual output of each sequencing lane.

*Library amplification and sequencing (all methods)*

Having identified the appropriate amount of library to use for sequencing as described above, libraries were PCR-amplified in a 50 μL reaction volume comprising 25 μL sample, 25 μL NEBNext Ultra II Q5 Master Mix (NEB M0544L) and unique dual index (UDI) containing PCR primers (dried). Primer sequence:

i5:AATGATACGGCGACCACCGAGATCTACAC[barcode]ACACTCTTTCCCTACACGACGCTC TTCCGATC*T

i7:CAAGCAGAAGACGGCATACGAGAT[barcode]GTGACTGGAGTTCAGACGTGTGCTCTTC CGATC*T

The reaction was cycled as follows, where $X$ denotes the number of PCR cycles: step 1, 98 °C 30 s; step 2, 98 °C 10 s; step 3, 65 °C 75 s; step 4, return to step 2 $X$ times; step 5, 65 °C for 5 min; step 6, hold at 4 °C. The number of PCR cycles ($X$) is dependent on the fmol input amount: 0.01-0.524 fmol, 16 cycles; 0.525-1.049 fmol, 15 cycles; 1.05-2.099 fmol, 14 cycles; 2.1-4.199 fmol, 13 cycles; 4.2-8.424 fmol, 12 cycles; 8.425-16.874 fmol, 11 cycles; 16.875-33.749 fmol, 10 cycles; 33.75-67.499 fmol, 9 cycles; 67.5-134.999 fmol, 8 cycles; 135-269.999 fmol, 7 cycles; 270-539.999 fmol, 6 cycles; 540-1079.999 fmol, 5 cycles; 1080-2159.999 fmol, 4 cycles.

The PCR product was subsequently cleaned up using two consecutive 0.7× AMPure XP clean-ups (hereafter referred to as SPRI clean-up; Beckman Coulter A63882). Each sample was quantified as described above.

*Hybridization Capture*

We determined a maximum DNA input per hybe capture reaction of 2,500 ng. For small targeted panels e.g. 0.9 Mb in size, we input up to 2,500 fmol per hybe reaction. For larger panels, e.g. whole exome, we input up to 1,500 fmol per hybe reaction.

To multiplex, we determined the *fraction of the hybe pool* that will be dedicated to each sample as follows: fmol of sample taken into PCR/total fmol in pool. We determined the ng of each sample that was taken into hybe capture as follows: *fraction of the hybe pool* × total ng of hybe pool. We determined the amount (μL) of each sample that was taken into hybe capture as follows: ng of sample into hyb/concentration of sample (ng/μL). For hybe capture we used Twist Bioscience Target Enrichment Standard Hybridisation v1 Protocol, with two rounds of capture to increase % on-target metrics. The following deviations were made to the protocol. 25 μL of an equimolar 200 μM pool of custom blocker sequences

CAA GCA GAA GAC GGC ATA CGA GAT (N:25252525)(N)(N) (N)(N)(N) (N)(N)G TGA CTG GAG TTC AGA CGT GTG CTC TTC CGA T*/3ddC/

GAT CGG AAG AGC ACA CGT CTG AAC TCC AGT CAC (N:25252525)(N)(N) (N)(N)(N) (N)(N)A TCT CGT ATG CCG TCT TCT GCT TG*/3ddC/

AAT GAT ACG GCG ACC ACC GAG ATC TAC AC(N:25252525) (N)(N)(N) (N)(N)(N) (N)AC ACT CTT TCC CTA CAC GAC GCT CTT CCG AT*/3ddC/
GAT CGG AAG AGC GTC GTG TAG GGA AAG AGT GT(N:25252525) (N)(N)(N) (N)(N)(N) (N)GT GTA GAT CTC GGT GGT CGC CGT ATC AT*T /3ddC/

were added to the indexed library pool before dry down (as opposed to adding universal blockers in step 2.3 of the Twist protocol; these were substituted with water). In step 3.22 of the Twist protocol, elution was carried out in 27 μL, as opposed to 45 μL. In step 4.4 of the Twist protocol KAPA HiFi was used for PCR amplification as opposed to Equinox. In addition, 2.5 μL of an equimolar pool (100 μM) of P5: 5′-AAT GAT ACG GCG ACC ACC GA-3′ and P7: 5′-CAA GCA GAA GAC GGC ATA CGA -3′ primers were used for PCR amplification, as opposed to the provided amplification primers. In Step 4.16 of the Twist protocol, elution was performed in 50 μL (as opposed to 32 μL) and all 50 μL were used to perform a second round of capture. The second capture was performed as the first, with the only deviation being the use of three PCR cycles.

**Supplementary Note 2. Selection analyses on single-molecule sequencing data**

To quantify the extent of selection and identify positively and negatively selected genes, we used several functions in the dNdScv R package (github.com/im3sanger/dndscv). Full details and reproducible code are provided in the supplementary files.

To identify genes under positive and negative selection we used the dNdScv algorithm as implemented in the *dndscv* function of the package. This algorithm is described in detail in the original publication [3]. Briefly, this is a maximum-likelihood implementation of dN/dS specifically developed for somatic mutation data. Mutations across genes are modelled using a context-dependent substitution model with 192 rate parameters shared across all genes, with each gene having its own dN/dS estimates for missense, nonsense and essential splice site mutations. The rate of somatic mutations has been shown to vary considerably across genes, often associated with expression levels and chromatin states [4]. The dNdScv algorithm models this variation using a negative binomial regression on the observed number of synonymous mutations per gene, using multiple epigenomic covariates, in practice modelling the unexplained variation in mutation rates across genes as being Gamma-distributed. Several likelihood ratio tests (LRTs) are used to detect selection on missense and truncating (nonsense and essential splice site) mutations, separately (*pmis*, *pnon*) and jointly (*pall*), and a separate negative binomial regression model is used to detect selection on indels (*pind*). A global *P*-value combining evidence from point mutations and indels is then calculated per gene using Fisher's combined *p*-value (*pglobal*). *P*-values were adjusted for multiple testing using Benjamini and Hochberg's false discovery rate. Below we describe several improvements introduced to the *dndscv* function in the present study.

*One-sided selection tests*

The original implementation of dNdScv was a two-sided test, which detects deviations from neutrality in any direction ($\omega \neq 1$, where ω is the symbol for dN/dS). One-sided tests for positive or negative selection could be done for independent mutation classes (e.g. missense or nonsense) by conditioning on dN/dS ratios being >1 or <1, but this was not possible for combined LRT *P*-values (*pall*) across mutation classes. As the dataset in the present study is large enough to enable negative selection tests at gene level, we have updated the *dndscv* function in the dNdScv package to include one-sided negative and positive selection tests (new optional argument *onesided* = T). In the two-sided implementation of LRTs the null hypothesis ($H_0$) is $\omega$mis = 1 and $\omega$non = 1, and the alternative hypothesis ($H_1$) is $\omega$mis ≠ 1 and $\omega$non ≠ 1. Instead, in a one-sided positive selection test, the maximum likelihood estimates for dN/dS ratios can take values <1 under the null hypothesis. In other words, the null hypothesis for a one-sided positive selection test is $H_0$: $\omega$mis≤1 and $\omega$non≤1, and for a one-sided negative selection test is $H_0$: $\omega$mis≥1 and $\omega$non≥1 [5].

To annotate essential genes based on CRISPR screens, we used a list of 2,023 essential genes from the DepMap database 24Q2 (CRISPRInferredCommonEssentials.csv file) [6], of which 17 overlap our target gene panel excluding genes under positive selection.

*Duplex coverage correction*

Since somatic mutations in targeted NanoSeq are called with single-molecule resolution, the density of detected somatic mutations per gene depends linearly on the duplex coverage achieved. To account for this additional source of variation in the mutation density per gene, we have modified the *dndscv* function to optionally use a vector of total duplex coverage per gene (summed across samples) (new optional argument *dc*). Internally, the expected mutation rate of each gene (the offset of the negative binomial regression in *dndscv*) is multiplied by the duplex coverage of each gene relative to the mean across genes. Although coverage affects mutation calling sensitivity in standard sequencing studies too, the relationship is non-linear and we only recommend using the optional *dc* argument in *dndscv* when working with targeted NanoSeq or other forms of single-molecule duplex sequencing, where detection sensitivity is directly proportional to duplex coverage.

*Custom reference database*

The 'SNP+noise' mask and the AS-XS filter used in the NanoSeq mutation calling pipeline result in the removal of mutations at these filtered sites. To ensure that the removal of common SNP and noisy sites did not introduce a small bias in dN/dS, these sites were also excluded from analysis in dNdScv by using a custom RefCDS object.

*Other arguments in dNdScv*

Additional non-default arguments used in the current study when running *dndscv* on the targeted data (**Supplementary Code**) include the following: (1) *max_muts_per_gene_per_sample = Inf, max_coding_muts_per_sample = Inf*; this disables the default cutoffs for the maximum number of mutations per gene or per sample that are only relevant for analysis of standard cancer genomic data. (2) *mingenecovs = 0, maxcovs = 10*; this ensures that the negative binomial regression in *dndscv* uses the first 10 epigenomic covariates (principal components), which is appropriate for a targeted sequencing study of this size. (3) *onesided = T*, (4) *use_indel_sites = F*; by default in dNdScv, only unique indel sites are counted to avoid a dominant effect of indel hotspots, such as microsatellites, but our filtering strategy (which excludes hypervariable sites, **Methods**) makes this unnecessary in the current dataset. Genes with significant evidence of selection on substitutions (as defined by *qall_loc*<0.01) were excluded from the fitting of the background indel model to avoid an inflation of the indel passenger rate by indel driver mutations (*kc* argument in *dndscv*).

In total, 49 genes were found to be under positive selection using the gene-level selection test, based on *qsubpos_cv*<0.01 (the combined *q*-value for point mutations, including missense and truncating substitutions). Similar results were obtained by alternative metrics, such as *qglobalpos_cv*<0.01, which includes evidence from indels (**Extended Data Fig. 5**), or *qpos_loc* <0.01, which uses the dNdSloc algorithm. dNdSloc is normally less powerful than dNdScv, as it only uses the number of synonymous mutations observed in a gene to estimate its background (neutral) mutation rate. However, the size of the buccal swab dataset is large enough to make dNdSloc similarly powered as dNdScv. dNdSloc does not rely on a negative binomial regression, epigenomic covariates or duplex coverage correction across genes, providing additional evidence that our selection results are robust to these assumptions.

*Hotspot discovery with sitednds*

Oncogenes are typically mutated at specific hotspot sites. The *dndscv* function does not exploit this information but two other functions in the dNdScv package are designed to test for selection at the level of individual sites (*sitednds*) or codons (*codondnds*). To detect sites with evidence of positive selection in our data, we applied *sitednds* using a lognormal-Poisson background model (LNP option) to the buccal swab and blood datasets on all genes in the panel. Genes with 4 or more amino acid changes under selection are shown in **Fig. 2l**. To increase sensitivity on known cancer hotspots, we then ran *sitednds* under restricted hypothesis testing (RHT) on 1,200 known oncogenic hotspots found to be significant in cancer and provided by the dNdScv package. *PIK3CA*, *ERBB2*, *KRAS* and *HRAS* each contained at least two amino acid changes with RHT *q*-value <0.01. The full lists of significant sites with and without RHT are provided in **Extended Data Table 4**.

*Withingenednds*

The high density of somatic mutations per gene in the current dataset enables the study of selection in specific groups of sites within a gene. To enable such analyses, we have added a new function to the dNdScv package called *withingenednds*. This function uses the output of *dndscv* to create a table of all possible mutations within a gene, annotating for each site its trinucleotide context, duplex coverage, observed number of mutations in the dataset, expected mutation rate based on the substitution, and amino acid change. Additional annotations (0/1 columns) are added for intron and exon flanks, sites in the last exon (where nonsense mutations are potentially not subject to nonsense-mediated decay), core promoter, and for additional user-defined regions (e.g. specific domains, groups of sites, neoantigens, etc). The function then uses a negative binomial regression across sites to obtain a separate ω (dN/dS) estimate for each layer of annotation. The function can fit a new overdispersion parameter for the rate variation across synonymous sites or use the θ parameter estimated across genes by *sitednds*. Wald and LRT (recommended) *p*-values are then calculated for each ω parameter (testing against neutrality, $\omega \neq 1$). In the present study, we ran *withingenednds* for all genes and *q*-values were calculated using the Benjamini–Hochberg procedure across genes for each functional class (shown in **Fig. 3e**).

*Estimation of the number of driver mutations*

To estimate the number of driver point mutations in the dataset, we used two alternative approaches (**Supplementary Code**). First, we used the global dN/dS ratios across all genes in the targeted panel to estimate the global excess of non-synonymous mutations in the targeted dataset. Briefly, the fraction of non-synonymous mutations predicted to be drivers (i.e. under positive selection) in a gene (or group of genes) can be estimated using: $(\omega-1)/\omega$ [3,7]. Using the global dN/dS ratio for all genes in the panel, this approach yielded an estimated number of driver point mutations of around 43,687 (CI95%: 42,458-44,898). As an alternative method, for the 49 driver genes found under positive selection by dNdScv, we summed the differences between the observed and expected mutation counts per gene for missense, nonsense and essential splice site mutations separately, as provided by the *dndscv* function in the *genemuts* output table. This yielded an estimated number of driver point mutations of ~43,314. Applying the latter approach for indels yielded an estimate of ~18,972 indel drivers. Combining point mutations and indels, this leads to an estimated number of driver mutations in the targeted buccal swab data of over 62,000.

*Physical interpretation of dN/dS ratios using single-molecule calling*

Standard deep sequencing of normal tissues only detects mutations that reach a certain VAF. In that context, as we have described before, dN/dS ratios can be interpreted as measuring the relative probability of a cell with a non-synonymous mutation in a gene reaching a detectable clone size (or VAF) in the cell population, compared to a cell carrying a synonymous mutation in the same gene [8].

In contrast, when using single-molecule (or single-cell) sequencing, the detection probability of a mutation is proportional to the frequency of the mutation in the sample or cell population. In that context, dN/dS ratios have a subtly different physical interpretation, measuring the relative increase in the number of cells affected by non-synonymous mutations in a gene compared to synonymous mutations as a result of clonal expansion or preferential survival, normalised by their respective mutation probability. As a result, dN/dS ratios obtained by single-molecule sequencing depend on the type and sizes of clonal expansions and (unlike standard sequencing) they should be unaffected by sequencing coverage. We note that this physical interpretation applies to dN/dS ratios obtained when counting mutations as many times as they are observed in a sample. In the current study, when running dNdScv, we have instead conservatively treated multiple mutant molecules reporting the same somatic mutation in a sample as a single mutational event. Since most mutations are only seen in a single read in the buccal swab data, this only leads to a slight underestimation of dN/dS ratios, but provides more robust *P*-values in dNdScv for the purpose of driver discovery by avoiding an undue influence of one or a few large clones on the substitution model and on dN/dS ratios.

### Supplementary Note 3. Whole-genome data and supplementary mutational signature analyses

*Whole-genome NanoSeq data*

Using the original restriction enzyme protocol [1], we performed whole-genome NanoSeq on 16 samples to better characterise the mutational processes acting on oral epithelium and to estimate genome-wide mutation rates. Of these samples, 12 were chosen to cover a wide age range, three donors were selected because of their strong Signature B (SigB) activity, and one donor was chosen because of their high mutation burden (associated with a history of chemotherapy) (**Extended Data Fig. 4a**).

COSMIC SBS16 has been previously associated to alcohol-induced mutagenesis in normal oesophageal epithelium [9], oesophageal tumours [10], and liver cancer [11]. The link between SBS16 and alcohol metabolism is further confirmed by its strong association with two risk alleles (*ALDH2* rs671 and *ADH1B* rs1229984) particularly common in Asia [9]. Previous studies have also shown that SBS16 has a characteristic pattern of transcription-coupled repair of the transcribed strand in expressed genes, and transcription-coupled damage of their untranscribed strand. This manifests as a lower rate of A>G mutations in the transcribed strand and a higher rate in the untranscribed strand compared to adjacent intergenic regions (or the opposite pattern when referring to these mutations as T>C changes).

To evaluate whether the observed SigB in the buccal swabs shows the same phenomenon of transcription-coupled damage and repair, we estimated the rate of T>C substitutions in the transcribed (template) and untranscribed (coding) strands, comparing it to 30 Kbp upstream and downstream the transcription start site and polyadenylation site, respectively. T>C rates were corrected to account for trinucleotide composition variations by normalising to whole-genome trinucleotide frequencies. Compared to flanking regions, our results show an increase in T>C rates in the transcribed strand and a decrease in T>C rates in the untranscribed strand in transcribed regions (**Extended Data Fig. 4b**). This pattern of transcription-coupled repair of A>G mutations in the transcribed strand and A>G transcription-coupled damage of the untranscribed strand is consistent with previous studies on SBS16 [11], providing further evidence that SigB in the oral epithelium is the same mutagenic process previously reported in oesophagus and liver cancers.

Next, we estimated mutation burdens for genomic regions with different chromatin states, using 15 chromatin states from the original ENCODE manuscript with sufficient number of mutations for analysis [12] (**Extended Data Fig. 4c,d**). Specifically, we used the chromatin states for E057 (foreskin keratinocytes) as reference. All the observed burdens in these chromosomal segments were normalised by the trinucleotide composition of each region and extrapolated to whole-genome trinucleotide frequencies. Using the set of 12 samples selected for their age range (see above), we observed higher burdens in heterochromatic regions (states "Het" and "Quies") and around transcription start sites (states "TssA" and "TssAFlnk"). Transcribed regions ("Tx" and "TxWk") showed lower burdens in this sub-cohort. The latter contrasts with the pattern observed in the 3 SigB-rich samples, in which transcribed regions showed higher burdens, especially for T>C substitutions, consistent with the process of transcription-coupled damage described above.

Mutational signature outliers

With only two mutational signatures extracted from the cohort, we wanted to examine whether any outlier donors had a mutational spectrum poorly explained by these two signatures. To systematically identify potential outliers, we re-fitted signatures SigA and SigB to each sample and calculated the cosine similarity between the reconstructed and observed mutational spectra. Cosine similarities are highly dependent on the number of mutations due to sampling noise (sparser profiles tend to have lower cosine similarities), so we plotted the cosine similarities as a function of the number of mutations in each sample. This analysis revealed a single strong outlier (**Extended Data Fig. 4e,f**), corresponding to a donor with a history of CHOP chemotherapy (cyclophosphamide, doxorubicin hydrochloride, vincristine sulfate, and prednisolone), typically used for the treatment of non-Hodgkin lymphoma.

### Extended sequence context analysis of T>C mutations

A high rate of T>C mutations at ApT dinucleotides is common to the COSMIC SBS5 and SBS16 signatures. To explore whether T>C mutations in SBS5 and SBS16 are caused by similar mutagenic processes, we studied the extended (pentanucleotide) sequence context of T>C mutations in several datasets. Specifically, we obtained transcriptional-strand-wise pentanucleotide mutational spectra for T>C mutations in buccal swab samples, matched blood samples, and hepatocellular carcinoma (HCC) samples from the Pan-Cancer Analysis of Whole Genomes study [13] that presented an SBS16 exposure >0.2 (**Methods**).

These pentanucleotide mutational spectra revealed that T>C mutations, especially those in ATN sequence contexts (i.e. at ApT dinucleotides), are the product of at least two distinct mutational processes with varying contributions across tissues (**Extended Data Fig. 8b**). In particular, T>C mutations in blood samples and buccal swab samples with low SigB exposure (<0.25, i.e. <25% of mutations attributed to SigB) are dominated by a process that largely matches the T>C component of SigA (resembling a combination of COSMIC SBS1 and SBS5). This component is characterised by a preponderance of T>C mutations at NATTG and NATAG contexts (and to a lesser extent NTTTG, NGTTG and NGTAG), with a bias towards mutations on the transcribed strand. On the other hand, T>C mutations in liver HCC samples and buccal epithelium samples with a SigB exposure >0.25 are dominated by the T>C component of SigB (resembling COSMIC SBS16), which is characterised by T>C mutations at NATAN and NATTN contexts (especially AATAN, TATAN and TATTT) and an extreme bias towards transcribed-strand mutations.

Thus, while COSMIC SBS5 and COSMIC SBS16 show similar T>C peaks at ATA and ATT contexts in the trinucleotide spectra (with subtly different contributions of the two trinucleotides), their pentanucleotide context is very distinct (**Extended Data Fig. 8b**). These results indicate that the T>C components of SBS5 and SigB/SBS16 reflect two distinct underlying mutational processes, both of which are associated with T>C mutations at ApT dinucleotides.

### Supplementary Note 4. Further description of the blood and buccal driver landscapes

*Driver landscape in blood*

Aggregating the duplex VAFs of all mutations detected in driver genes, we can estimate the fraction of cells carrying driver mutations in a polyclonal sample, without the limitations of incomplete detection sensitivity of traditional bulk sequencing (**Methods**). This revealed that, on average across samples, ~1-2% of blood cells in donors aged 65-85 carry *DNMT3A* or *TET2* driver mutations, with values ranging 0.1-0.5% for other driver genes. Although we note that this number varies widely across donors due to the presence of large clones in the blood of some individuals (clonal haematopoiesis).

The genes under positive selection in blood are associated with the following biological processes: epigenetic modification and chromatin remodelling (*DNMT3A*, *TET2*, *ARID2*, *ASXL1*, *KMT2E* and *KDM6A*); DNA damage response and cell cycle control (*ATM*, *CHEK2*, *TP53*, *PPM1D* and *CDKN1B*); receptor tyrosine kinase signalling (*NF1* and *CBL*); G-protein coupled receptor signalling (*GNB1*); inflammation mediation (*JAK2*, *STAT3* and *MYD88*); transcription regulation (*FOXP1*); and mRNA splicing (*SRSF2* and *SF3B1*). Specific genes are discussed in more detail below.

*DNMT3A* encodes a DNA methyltransferase that modifies cytosine in CpG contexts to 5-methylcytosine. Unlike DNMT1, which maintains genomic methylation patterns during DNA replication, DNMT3A is responsible for establishing de novo methylation. *DNMT3A* is the most frequently mutated driver of clonal haematopoiesis [14]. As has previously been observed [15], the distribution of mutations in *DNMT3A* differs between cancer and clonal haematopoiesis. In acute myeloid leukaemia (AML), most *DNMT3A* mutations (~55%) occur at arginine 882, which is recurrently mutated to histidine or, less frequently, cysteine [16]. By contrast, R882H/C accounts for only ~10% of *DNMT3A* mutations in clonal haematopoiesis, with other mutations rarely seen in AML occurring instead, including Y735C, V657M and R736H/C missense mutations, essential splice site mutations and indels (**Extended Data Fig. 3d**). The R736 and Y735 mutations occur at the interface between the central and peripheral subunits in DNMT3A homotetrameric and DNMT3A/DNMT3L heterotetrameric complexes [17].

*TET2* is the second most common driver of clonal haematopoiesis. *TET2* encodes an Fe(II) and α-ketoglutarate-dependent dioxygenase that mediates DNA demethylation by converting 5-methylcytosine to 5-hydroxymethylcytosine. Truncating mutations in *TET2* are frequently seen in both clonal haematopoiesis and cancer [15]. In addition to nonsense mutations and indels distributed throughout *TET2*, we observed a cluster of missense mutations affecting β strands 15 to 17 of the dioxygenase domain [18] (**Extended Data Fig. 3e**). Unlike most studies of clonal haematopoiesis conducted using standard sequencing, we observed more non-synonymous mutations in *TET2* ($n$ = 1,104) than *DNMT3A* ($n$ = 800), which also corresponded to a higher estimated number of driver mutations in *TET2* ($n$ = 853 and 743 respectively). However, we obtained substantially higher cumulative duplex coverage across *TET2* (436,473 dx) than *DNMT3A* (177,102 dx). Correcting for this difference, the *DNMT3A*:*TET2* ratio of observed non-synonymous mutations or estimated driver mutations per duplex coverage (1.79 and 2.15 respectively) is substantially closer to the *DNMT3A*:*TET2* ratio of non-synonymous mutations observed in a large clonal haematopoiesis study (2.37) [14]. The remaining discrepancy may reflect differences in the exponential growth of *DNMT3A*- and *TET2*-mutant clones (**Fig. 4a**), for example due to lower fitness coefficients for some *TET2* mutations making them less likely to reach detectable clone sizes with standard bulk sequencing.

*CBL* encodes a protein that acts as both a positive and negative modulator of receptor tyrosine kinases (RTKs). RTK signalling is downregulated by the E3 ubiquitin ligase activity of the RING domain of CBL, which targets the RTK for degradation. Upregulation of RTK signalling is mediated by the adaptor function of CBL, which recruits signalling molecules (such as phosphoinositide 3-kinase) to the RTK [19]. As previously seen in cancers, we observed clustering of missense mutations within the RING domain, which abrogates the ubiquitin ligase activity of CBL while retaining its adaptor function (**Extended Data Fig. 3f**).

*MYD88* encodes an adaptor protein for interleukin-1 receptors and Toll-like receptors. *MYD88* L265P mutations are frequently observed in diffuse large B-cell lymphoma, Waldenström macroglobulinemia and chronic lymphocytic leukaemia, resulting in NF-kB activation [20]. Although *MYD88* was not identified by gene-level selection analyses, this hotspot was found to be significantly recurrent by *sitednds* (**Extended Data Fig. 3g**, **Supplementary Note 2**, **Extended Data Table 4**). Similarly, known activating hotspots were identified in *JAK2* (V617F), *SF3B1* (Y765C and K700E), *SRSF2* (P95H), *GNB1* (K57E) and *STAT3* (S614R, G618R, Y640F and D661Y).

*Driver landscape in oral epithelium*

Of the 49 genes identified as under significant positive selection in oral mucosa (**Extended Data Fig. 5a**), 10 were also identified as driver genes in blood: *TET2*, *DNMT3A*, *ATM*, *ARID2*, *ASXL1*, *KDM6A*, *CHEK2*, *PPM1D*, *TP53* and *FOXP1*. One potential explanation for this overlap could be that clonal haematopoiesis drivers are detectable in buccal swabs due to blood contamination. Using targeted enzymatic methylation sequencing, we estimated the median epithelial purity of the buccal swabs to be 95.1% (**Methods**, **Extended Data Fig. 1h**). This was concordant with an estimate of 7-8% median blood contamination obtained by attempting to detect in the buccal swabs clonal haematopoiesis mutations that reached levels observable by standard sequencing approaches (VAF≥1%) in the matched blood (**Methods**). Despite this relatively high epithelial purity, the observed degree of contamination corresponds to ~34,000 dx of the 693,208 dx cumulative coverage in buccal swab samples being derived from blood. Therefore, we additionally calculated the buccal:blood driver density ratio for each of these genes (**Extended Data Fig. 3c).** For all of the genes apart from *DNMT3A*, *TET2* and *FOXP1*, the estimated driver density was higher in the buccal swab samples than blood. This strongly suggests that the other 7 genes are bona fide drivers in oral mucosa. Additional evidence that the majority of these genes are drivers in squamous epithelium comes from the previous observation that *TP53*, *PPM1D*, *CHEK2* and *ARID2* are under significant positive selection in physiologically normal oesophagus and *KDM6A* is a known driver in oesophageal squamous cell carcinoma [9,21].

Excluding the three genes where the signal of selection could be attributable to blood contamination (*TET2*, *DNMT3A* and *FOXP1*), the remaining 46 genes under positive selection in oral mucosa are associated with the following biological processes: Notch signalling pathway (*NOTCH1*, *NOTCH2* and *NOTCH3*); DNA damage response and cell cycle control (*TP53, ATM*, *CHEK2*, *PPM1D* and *CCND1*); epigenetic modification and chromatin remodelling (*KMT2C*, *KDM5C*, *KDM6A*, *ASXL1*, *ARID1A*, *ARID1B*, *ARID2*, *ARID5B*, *SETD2*, *EP300* and *SMARCB1*); regulation of transcription (*MGA*, *BCORL1*, *CIC*, *TP63*, *ZNF750*, *FUBP1*, *KLF5*, *PAX9*, *ETV6* and *RARG*); RNA processing (*ZFP36L2*, *ZFP36L1*, *RBM10* and *SF3B1*); receptor tyrosine kinase signalling (*EPHA2*, *EGFR*, *FGFR3* and *PIK3R1*); cell adhesion and cytoskeletal organisation (*FAT1*, *RAC1*, *RHOA* and *AJUBA*); protein turnover (*CUL3*, *SPOP* and *EIF1AX*); sister chromatid separation (*STAG2*); and antigen presentation (*HLA-B*). Specific genes and pathways are discussed in more detail below.

*Notch signalling pathway genes*

As has previously been observed for other squamous epithelia [7,9,21], the most frequently mutated driver gene in oral mucosa is *NOTCH1*. NOTCH1 is a cell surface receptor with an extracellular domain (residues 19-1735) and a cytoplasmic domain (residues 1757-2555). The extracellular domain consists of 36 EGF-like repeats (residues 20-1426), three Lin-12-Notch repeats (residues 1442-1571) referred to as the Notch domain, a NOD domain (residues 1566-1622) and a NODP domain (residues 1670-1732). The cytoplasmic domain consists of multiple ankyrin repeats (residues 1871-2126) and a C-terminal domain (residues 2478-2541), which includes a PEST domain. Upon binding one of its ligands (JAG1, JAG2, DLL1 or DLL4) via EGF repeats 8-12 [22], NOTCH1 undergoes a conformational change that exposes a cryptic proteolytic site. Subsequent cleavage of this site by a member of the ADAM protease family followed by cleavage of a second proteolytic site by $\gamma$-secretase releases the intracellular domain of NOTCH1 from the plasma membrane. The liberated intracellular domain is imported into the nucleus and mediates expression of Notch target genes via association with RBPJ and MAML1. Degradation of the intracellular domain is accelerated by the presence of an intact PEST domain.

Consistent with the pattern seen in other squamous epithelia, we observed nonsense mutations, essential splice site mutations, insertions and deletions distributed throughout the gene body of *NOTCH1*, as well as a cluster of missense mutations affecting the ligand binding region (EGF repeats 8-12, residues 295 to 488) (**Fig. 3c**). Consistent with missense mutations in EGF repeats 8-12 altering protein function, this region also had a high proportion of deletions and insertions that are in-frame (66% and 37% respectively), whereas the vast majority of deletions and insertions in the rest of the gene result in a frameshift (92.5% and 98% respectively). These mutations are expected to impair Notch signalling, which mediates a program of cell cycle arrest and terminal differentiation in squamous epithelia [23].

By contrast, Notch signalling promotes self-renewal of haematopoietic stem cells [24] and so activation of this pathway is a common feature of haematological malignancies. Therefore, the pattern of *NOTCH1* mutations in lymphomas and leukaemias is markedly different to squamous cell carcinomas, with recurrent missense mutations and in-frame insertions and deletions affecting the NOD domain and nonsense and frameshift mutations occurring immediately prior to the PEST domain. The latter mutations cause C-terminal truncation of the protein rather than nonsense-mediated decay of the mRNA, resulting in stabilisation of the intracellular domain due to loss of the PEST domain. Given the differential impact of nonsense mutations that occur after the final exon junction complex in a gene, we expected to observe comparatively few truncating mutations in the last exon of *NOTCH1* in oral mucosa. Indeed, the dN/dS ratio for nonsense mutations in the final exon was substantially lower than across the rest of the gene (6.9 and 68.4 respectively). Of the 73 nonsense mutations present in the last exon of *NOTCH1*, 64 of them occurred in the 9 sites closest to the exon junction complex (out of 117 possible nonsense sites). We observed no enrichment of nonsense mutations across the remaining 108 sites (9 observed, 11.3 expected).

Of the 599 amino acid changes in *NOTCH1* found to be under significant positive selection by site-level dN/dS (*sitednds*), 5 codons were affected by recurrent synonymous mutations (V199, G394, G472, T634 and C1528) (**Extended Data Table 4**). The mutations in two of these codons (C1528 and T634) occur extremely close (3 bp and 2 bp upstream respectively) to an exon-intron boundary and are predicted by SpliceAI [25] to disrupt the nearby splice donor site (donor gain $\Delta$ score = 0.9 for 9:139399764 G>A, donor loss $\Delta$ scores = 0.95 and 0.93 for 9:139409936 T>A and T>C respectively). The other three sites (G472, G394 and V199) are located further away from existing exon-intron boundaries but are predicted with high confidence to generate novel splice donor sites by SpliceAI (donor gain $\Delta$ scores = 1, 0.99 and 0.8 for 9:139412229 C>A, 9:139412662 G>A and 9:139417447 G>T respectively). This pattern of synonymous mutations affecting splicing has previously been observed in *TP53* (codons T125, E224 and Q331) [26] and two sites in codon T125 of *TP53* (17:7579312 C>T and C>A) were significantly recurrently mutated in our cohort. However, there were only 19 such recurrent synonymous mutations amongst the 1,220 amino acid changes under significant positive selection in oral mucosa, suggesting that this is a rare class of driver mutation in normal tissue, as well as in cancer [3], and so is less frequent than has previously been suggested [26].

Both *NOTCH1* and *NOTCH2* (as well as *TP53* and *CHEK2*) exhibited significant enrichment of mutations at sites in the 10 bp flanks of introns that are not classified as essential splice site mutations by dNdScv (**Fig. 3e**). For *NOTCH1*, 269 mutations were observed compared to ~86 mutations expected by chance under neutrality. Several of the most recurrent sites in *NOTCH1* (9:139410175 C>T observed 34 times, 9:139400339 C>T observed 14 times, 9:139400340 G>T observed 8 times, 9:139410554 C>T observed 7 times and 9:139396548 G>C observed 5 times) are all predicted to generate novel splice acceptor sites by SpliceAI (all have acceptor gain $\Delta$ scores = 1). The mean and median acceptor gain $\Delta$ scores across all 1,452 intronic flanking sites in *NOTCH1* were 0.049 and 0 respectively. Additionally, we observed high confidence examples of acceptor loss mutations (e.g. 17:7578292 A>C in *TP53* observed 7 times with acceptor loss $\Delta$ score = 0.89) which introduced a guanine at the -3 position of the splice acceptor site, which is the only base not tolerated by typical U1- and U2-type splice sites [27]. These results highlight the potential benefit of extending the definition of splice impacting mutations beyond specific positions in the splice acceptor and donor regions.

It has previously been reported that the proportion of cells in histologically normal oesophagus that contain a *NOTCH1* driver mutation by middle age (30-80%) is much higher than the proportion of oesophageal squamous cell carcinomas that bear *NOTCH1* mutations (~10%) [21]. This has led to the proposal that while *NOTCH1* mutations are capable of driving clonal expansion in normal oesophageal epithelium, *NOTCH1* clones have a lower risk of further evolving into cancers than wild-type cells [21] and may even impair tumour growth by clonal competition [28]. The same protective effect does not necessarily appear to be the case for oral mucosa as the frequency of *NOTCH1* driver mutations in oral cancer (~16%) is comparable to the mutant cell fraction in normal oral mucosa (~10%) (**Fig. 2h**). However, the similar frequency of *NOTCH1* mutations in normal oral epithelium and oral cancers still suggests that *NOTCH1* mutations do not confer a considerably higher tumourigenic risk in oral epithelium.

The distribution of mutations in *NOTCH2* (**Extended Data Fig. 6a)** closely resembles that of *NOTCH1* with loss-of-function mutations distributed throughout the gene body, comparatively few truncating mutations in the final exon (dN/dS ratio of nonsense mutations in last exon and rest of gene are 17.1 and 1.47 respectively) and a cluster of missense mutations in the ligand binding region. We were unable to call many mutations in the first four exons of *NOTCH2* due to the high degree of homology in this region to another gene on chromosome 1q (*NOTCH2NLA*) [29], thus failing our AS-XS cut-off (**Methods, Extended Data Fig. 2**). Of note, several highly recurrent mutations are called in these exons in the COSMIC database (C19W, A3S/V/F, A21T, R5R, N46S and E38K) and so these should be treated with caution as potential alignment artefacts.

*DNA damage response and cell cycle control genes*

*TP53* is the most frequently mutated driver gene across many cancer types [3] and has been extensively studied. The distribution of mutations across the gene body in oral mucosa is remarkably concordant with the pattern previously observed in squamous cell carcinoma (**Fig. 2a)**, with truncating mutations distributed throughout the gene and missense mutations occurring at specific residues in the DNA binding domain (residues 100-288), which are known to have a dominant negative effect by forming tetramers with wild-type p53 that are unable to bind DNA with high affinity [30].

There was a high degree of overlap between the sites identified as significant in oral mucosa without restricted hypothesis testing and previously known hotspots from The Cancer Genome Atlas (218/279; 78%) and a similar proportion of coding substitutions occurring at the most frequently mutated hotspot codons in oral mucosa compared to squamous cell carcinoma (R273: 5.4% vs 3.7%; R175: 3.4% vs 3.7%; R248: 3.2% vs 4.7%; R282: 2.5% vs 3.1%; H179: 2.4% vs 2.7%; H193: 2.1% vs 1.7%). However, one codon accounted for a strikingly different proportion of coding *TP53* substitutions. P177 is the 7$^{th}$ most frequently mutated codon in oral mucosa (2.1%) but is rarely mutated in squamous cell carcinoma (0.08%). This difference was also observed in a meta-analysis of cancer and non-neoplastic tissue sequencing studies [31]. Proline 177 is located in the short H1 $\alpha$ helix that lies within the L2 loop of p53 and is situated between two of the four residues (C176, H179, C238 and C242) that tetrahedrally co-ordinate the zinc ion [32]. In an early site-directed mutagenesis assay of *TP53*, it was found that P177H only partially reduced the transcriptional activation activity of p53 across 8 assayed target promoters, whereas known hotspot mutations (such as R175H) almost completely inactivated p53 [33]. Another difference between the mutation distribution in oral mucosa and squamous cell carcinoma is the proportion of coding substitutions that are nonsense mutations (10.7% vs 19.1% respectively), potentially suggesting that dominant negative missense mutations provide a greater selective advantage in normal tissues than heterozygous truncating mutations.

There was also noticeable enrichment of several mutations outside the coding region of *TP53* (**Fig. 3f**). Non-coding drivers in *TP53* affecting the transcription start site or donor splice site of the first non-coding exon have been previously described by the Pan-Cancer Analysis of Whole Genomes (PCAWG) consortium [34]. We also observed recurrent mutations affecting the canonical GGTAAG sequence [27] at the -1 to +5 positions of the splice donor site of the first non-coding exon (7:7590690-7590695), with 64 substitutions observed compared to 0.8 expected. *TP53* was one of the genes that exhibited significant enrichment of substitutions in its core promoter region (±200 bp of the transcription start site) compared to a background model of synonymous mutations within the gene (**Fig. 3e**). These substitutions were broadly distributed throughout the promoter region with a relatively low degree of recurrence (≤6 mutations) at any given site, apart from a single hotspot of 10 substitutions occurring 2 bp upstream of the transcriptional start site (17:7590858). We note that several genes that were not classed as drivers by any other type of mutational enrichment had a similar diffuse pattern of substitutions in their core promoter (**Fig. 3e**) and across all genes there was an excess of substitutions in the 5' UTR immediately after the transcription start site (**Fig. 3f**). This likely indicates that in many cases the observed excess of substitutions in the core promoter is due to increased mutability of this region rather than selection. However, unlike the other genes, *TP53* had a large number of insertions and deletions ($n$ = 43) within its core promoter, which is consistent with the enrichment previously described in cancer [34,35].

Two other genes with markedly different patterns of mutations in their promoters were *TERT* and *SRSF2*. *TERT* promoter mutations are well-known drivers in cancer that create novel ETS transcription factor binding sites [36,37]. In addition to the two canonical *TERT* promoter mutations (5:1295250 G>A observed 38 times; 5:1295228 G>A observed 23 times), we also observed several other highly recurrent sites (5:1295158 G>T observed 21 times; 5:1295161 T>G observed 16 times; 5:1295149 C>A observed 10 times), which have previously been observed infrequently in cancer-derived specimens [38]. *SRSF2* had a cluster of mutations within its core promoter (63 mutations observed vs 1.4 expected in 17:74733377-1774733390, of which 37 occur at 17:74733380), the consequence of which is unclear.

An additional non-coding region of *TP53* that exhibited a striking enrichment of mutations was the polyadenylation signal (31 mutations observed vs 0.7 expected in 17:7571751-7571756, of which 25 occur at 17:7571755). Germline variants that disrupt the polyadenylation signal (rs78378222; 17:7571752 T>G) have previously been reported to confer increased susceptibility to cancer [39]. Of note, we did not call somatic mutations at 17:7571752, as the population frequency of rs78378222 is sufficiently high (1.2% [40]) that this site is excluded by our SNP mask (**Methods**).

*Saturation analysis*

As described in the main text, ultra-deep single-molecule sequencing of polyclonal samples has the potential to provide a form of in vivo saturation mutagenesis. In this section, we describe some supplementary analyses on the extent of saturation achieved in the current study.

First, a valuable metric can be the density of mutations per site at neutral sites. This value depends on the mutability of each site, which is particularly affected by the trinucleotide sequence context of each base. The mean neutral mutation rate per site for each trinucleotide substitution is calculated in the substitution model of dNdScv (*dndsout$mle_submodel*). In the buccal swab dataset, the highest average neutral mutation rate per site was ~0.43-0.60 mutations/site for C>T changes in all four possible CpG contexts, and the lowest rates per site were ~0.007-0.008 mutations/site for A>C mutations at certain contexts (**Supplementary Code**). The mean rate across all 192 possible trinucleotide changes was ~0.056 mutations/site. These rates refer to the neutral mutation rate for each possible trinucleotide change. When considering SNVs, each base can change to three other bases (e.g. A can change to C, G or T), and each codon can change to nine other codons, and so the average neutral mutation density per base pair or per codon will increase accordingly. This analysis reveals that ~2 or ~6 times higher aggregate depth than currently achieved will be required to obtain an average of one mutation per neutral codon or base pair, respectively.

The description above refers to the neutral mutation density per base change, per base pair, or per codon (or amino acid). However, we note that lower aggregate depths will be needed to find the most important sites under strong positive selection (e.g. with site-dN/dS > 100), while much higher depths will be required to find individual sites under negative selection. To explore the extent to which the landscape of driver mutations is approaching saturation in our dataset, we carried out a downsampling exercise, studying the number of genes and sites under significant positive selection for progressively larger random subsets of our dataset (**Extended Data Fig. 6j,k**). At gene level, this analysis suggests that a dataset half the size of the current dataset is sufficient to find ~80% of the 49 driver genes reported here, and that these genes account for ~90% of all non-synonymous mutations in driver genes. At single-site level (*sitednds*), the pattern of saturation varies considerably across genes. For example, the discovery of sites under positive selection in *RAC1* and *PPM1D* shows clear signs of saturation, where a dataset half the size of the current dataset is enough to find the hotspots responsible for >80% of the mutations at significant hotspots in the full dataset. A trend towards saturation is apparent but slower for *NOTCH1* and *TP53*, suggesting that larger datasets will identify additional relevant sites under positive selection. In contrast, other genes under weaker selection and where selection is spread across many sites, such as *CHEK2* and *NOTCH2*, show no clear trend of saturation in subsamples of the current dataset.

Overall, the current dataset represents an in depth description of the landscape of driver genes and driver sites in the oral epithelium, but larger studies are expected to continue to yield new sites under positive selection, particularly in genes under weaker positive selection. We also note that much larger datasets will be needed to comprehensively study the pattern of negative selection at the level of single genes, and particularly at the level of single sites.

**Supplementary Note 5. Multistage models linking mutation rates and clonal expansions to risk**

Systematic somatic mutation studies in large cohorts of individuals will help build mechanistic models of cancer risk. By performing bulk sequencing studies with single-molecule sensitivity it is now possible to measure mutation rates, mutational signatures and clonal frequencies across individuals with different risk factors (e.g. smoking or alcohol consumption), as well as in cases and controls (e.g. sampling normal tissue from individuals recently diagnosed with cancer). Regression models could then be used to establish quantitative relationships between risk factors (genetics, exposures, lifestyle…) and changes in mutation rates and clonal landscapes, and between these and cancer risk (**Fig. 4j**).

In this context, a brief revision of classical multistage models of carcinogenesis can help understand how changes in mutation rates or clone sizes are expected to alter cancer risk under different assumptions. To this end, we summarise some classical multistage models of carcinogenesis with and without clonal expansions, and discuss them in light of recent discoveries from normal tissues, including those from the present study. Although these classical models are overly simplistic, they provide a framework to understand important features in carcinogenesis and can offer a starting point to build more realistic and empirical mechanistic risk models.

Multistage models without clonal expansions

The incidence rate of most cancers increases rapidly with age. For several major cancer types, the increase in incidence during the ages of 20-75 is approximately proportional to age to the power of 5 or 6. In the early 1950s, Nordling (1953) [41] and Armitage & Doll (1954) [42] showed that this behaviour can be recapitulated by a simple model where stochastic somatic changes (mutations or other somatically-heritable changes) occur in cells with a constant rate throughout life[1] and cancer results when 6 or 7 key changes[2] have accumulated in a single cell. If the probability of a driver mutation ($p$) is low enough, the incidence rate of cancer at age $t$ follows **equation (1)**:

$$I(t) = k\, p_1\, p_2 \ldots p_n\, t^{(n-1)}$$

$n$ being the number of changes required to transform a normal cell into a cancer, and $k$ being a constant term. Under this model, cancer incidence is proportional to $t^{(n-1)}$, that is: $I(t) \sim t^{(n-1)}$. Using a log transformation, we obtain: $\log(I(t)) \sim (n-1)*\log(t)$, which allows estimation of $n$ using the slope of a $\log(I)$-$\log(age)$ plot. This led Armitage & Doll (1954) to predict that many cancers may be the result of 6 to 7 somatic changes in a cell[3].

This early multistage model of carcinogenesis has been highly influential. Despite being too simplistic, this model explains two key aspects of carcinogenesis that still cause confusion among cancer researchers today. First, it provides a simple explanation for how cancer incidence is expected to increase rapidly with age, without needing to invoke tissue ageing or mutation rate acceleration with age. In fact, it may provide a simple conceptual model for other ageing processes, by showing how a linear accumulation of somatic changes in our tissues can lead to rapid (geometric) increases in morbidity if multiple changes in a cell or in separate cells are needed for disease. Second, the recent

[1] The assumption that somatic mutation rates are constant throughout life was unsupported at the time, and in fact some authors expected mutation rates to increase exponentially with age as a result of age-related loss of repair and control mechanisms ("error accumulation" models). However, recent sequencing studies across a range of somatic tissues have shown that somatic point mutations and indels accumulate linearly with age.

[2] Given the importance of somatic mutations in cancer development, the *somatic changes* in multistage models are often assumed to refer to somatic driver mutations, but can refer to epigenetic changes and other changes that are somatically heritable. Under some models, the equations hold if some of the changes occur in other cooperating cells too [43].

[3] The reason why the exponent is $n$-1 rather than $n$ is because the incidence rate of a tumour in one particular day depends on the cumulative number of cells with $n$-1 events in the tissue ($\sim t^{n-1}$), multiplied by the probability of the $n^{th}$ event per cell ($p_n$).

discovery that human ageing tissues contain large numbers of cells with one or two cancer-driver mutations has led some to question whether these mutations truly contribute to carcinogenesis. Although the 1954 Armitage & Doll model did not incorporate clonal expansions, their equation predicts that for a tumour to appear with 6-7 changes, tissues may be expected to carry very large numbers of cells with 1 or 2 genuine cancer-driver mutations.

The prediction that 6 or 7 somatic changes in a single cell might be required to explain cancer was controversial at the time, as it seemed an excessively complex model of carcinogenesis in the absence of any detailed mechanistic understanding. This led Armitage & Doll, and other contemporary authors, to propose alternative multistage models incorporating clonal expansions, which could explain human cancer incidence statistics with as few as 2 or 3 somatic changes (see the following section). In the past 15 years, however, cancer genomic studies across cancer types have confirmed that most human tumours carry multiple driver mutations as well as extensive aneuploidy, suggesting that a multistage model of carcinogenesis is, at least, a reasonable conceptual framework [3,13]. This is also supported by genomic studies of premalignant lesions in some cancer types with histologically distinct precursor lesions [e.g. [44–46]].

Multistage models with clonal expansions

In the last decade, advances in DNA sequencing have led to the discovery that clonal expansions are widespread in proliferating somatic tissues. By middle age, ~25% of all cells in normal skin [7], around 40% of cells in normal oesophagus [21], >50% of all cells in endometrial glands [47], ~10-20% of cells in normal bladder urothelium [8,48] and ~10-20% of cells in oral epithelium (present study) carry a driver mutation due to positive clonal selection. In light of these discoveries, it is useful to revisit some classical multistage models that incorporated clonal expansions. These models change the predicted impact of mutagenic carcinogens on cancer risk, and they offer a starting framework for modelling the role of non-mutagenic carcinogens (promoters or selectogens).

In the classical Armitage and Doll model (1954) [42] driver mutations accumulate in single cells without conferring a clonal advantage until the final ($n^{th}$) change. Around the same time, in 1955, Platt suggested that carcinogens may induce changes to cells that make them proliferate faster [49] Motivated by this idea, in 1957 Armitage and Doll proposed a model where driver mutations lead to exponential clonal expansions, which increase the number of cells at risk of subsequent hits. With this model, they showed that the rapid increase of cancer incidence with age observed in multiple cancer types could be explained by a model with as few as two driver mutations, if mutations induce exponential clonal expansions [50]. This led other authors to propose alternative models with different modes of clonal growth, showing that the epidemiological data could be fitted by quite disparate models.

The models introduced by JC Fisher in 1958 [51] are of particular relevance for modelling cancer risk as a result of mutation and clonal expansion in flat epithelia. Fisher argued that in a flat epithelium where clonal competition occurs at the edges of a clone, clones do not grow exponentially but quadratically. If the growth of a clone is quadratic, the number of cells in the clone after time $t$ since the occurrence of the driver mutation is given by **equation (2)** (where $C$ is a constant):

$$\text{clone size} = C\, t^2$$

Introducing only one quadratic clonal expansion into the original Armitage & Doll 1954 model consequently predicts that the incidence of cancer should still increase geometrically with age, as follows (**equation 3**):

$$I(t) = k\, p_1\, p_2 \ldots p_n\, t^2\, t^{(n\text{-}1)}$$

$$I(t) \sim t^{(n+1)}$$

If every sequential driver mutation induces a quadratic clonal expansion, we obtain **equation (4)**:

$$I(t) \sim t^{3(n\text{-}1)}$$

This model still follows the simple power-law increase of cancer incidence with age ($I \sim t^k$) reported by Armitage and Doll, but it requires considerably fewer driver mutations (or changes) for cancer development. For example, a cancer type that increases in incidence as a function of age to the power of 6 can be explained by 7 independent driver mutations without clonal expansions (1954 Armitage and Doll's model), or by just 3 driver mutations where the first two events led to subsequent quadratic clonal expansions (Fisher 1958).

This framework can be used to understand the impact of other types of clonal growth (see **Supplementary Note 6**). For example, let us imagine a tissue where clones expand to a typical maximum size (*L*) and do not grow beyond it. This could be due to spatial constraints (such as colonic crypts), cell-intrinsic mechanisms (e.g. telomere shortening or oncogene-induced senescence) or cell-extrinsic mechanisms (e.g. immune surveillance). Under such a model, rapid clonal expansions increase the probability of subsequent hits by a factor *L*, but do not alter the exponent of the original Armitage and Doll model (**equation 5**):

$$I(t) = k\, p_1\, p_2 \ldots p_n\, L^{(n\text{-}1)}\, t^{(n\text{-}1)}$$

$$I(t) \sim t^{(n\text{-}1)}$$

As discussed in **Supplementary Note 6**, the approximately linear increase in driver frequency with age that we observe in our buccal swab data suggests that clonal expansions are constrained in oral epithelium (**Fig. 4a**). Given that new mutations occur at an approximately constant rate throughout life (**Fig. 4c**), neither exponential nor quadratic clonal expansions seem consistent with the observed approximately linear increase in driver density with age, which instead appears more consistent with equation (5). In fact, the apparently sublinear increase in frequency of *TP53*-mutant clones with age, despite the continuous occurrence of new mutations in *TP53*, suggests a considerable decline in the relative fitness of these clones with age, a feature not captured by the models above.

*Hypermutation versus clonal expansion debate*

The discovery of the widespread existence of clones carrying cancer-driver mutations in normal tissues, and the multistage models above, are also relevant in clarifying a decades-long debate on the role of hypermutation versus clonal expansion in carcinogenesis [e.g. [52–56]]. Estimates of the somatic mutation rate available in the 1980s-2000s, although inaccurate, suggested that normal somatic mutation rates were too low to explain the emergence of cancer under the classical Armitage and Doll 1954 model[4]. This led to two alternative hypotheses: (1) carcinogenesis may require the emergence of hypermutator cells, or (2) cancer may not require the evolution of hypermutation, but require intermediate clonal expansions. The role of both processes in increasing the probability of a cell acquiring the full complement of driver mutations required for transformation can be understood in the models above. Data from cancer genomics and from somatic mutation studies of normal tissues have now largely resolved this debate. They suggest that both hypermutation and intermediate clonal expansions are important factors in carcinogenesis. Clonal expansions have been observed in most mitotic tissues studied to date, and comparison of mutation rates and mutational signatures in cancer genomes and in

---

[4] Somatic mutation rates across most cell types are now known to be on the order of $2\times10^{-7}$ to $4\times10^{-7}$ mutations/bp/cell by middle age [1,57]. Most cell types have a modest number of genes that can act as drivers, typically tens of genes [3]. If we assume that there are on the order of ~10,000 possible driver sites in a genome, and that 6-7 changes are needed to transform a cell, the fraction of cells in a tissue expected to carry one driver mutation (in the absence of selection and clonal expansion) could be as high as ~0.2-0.4%, whereas the probability of one cell acquiring 6 driver mutations by chance would be $<1\text{x}10^{-14}$. This probability is orders of magnitude too low to explain the observed incidence of cancer given the estimated numbers of stem cells per tissue, highlighting the importance of hypermutation and/or intermediate clonal expansions to explain cancer incidence.

normal tissues shows that increased mutation rates and new mutational processes, including chromosomal instability generating extensive aneuploidy, are common features of many tumours and some premalignant lesions.

Carcinogenesis as somatic evolution

Cancer development is best understood not as a result of somatic *mutation* but of somatic *evolution*; that is, the result of both somatic mutation and clonal selection. As is the case for natural selection in species evolution, clonal selection can change over time (it is context-dependent) and encompasses cell-intrinsic changes (e.g. altering the rate of cell division, apoptosis, differentiation, etc) as well as ecological interactions with the cellular microenvironment (e.g. clonal competition, spatial constraints, immune surveillance, etc).

The incorporation of clonal expansions in multistage models is important for several reasons. First, they offer a more complete model of carcinogenesis, where carcinogens can increase cancer risk/incidence by either inducing mutations (mutagens) or altering selection and clonal expansion (selectogens/promoters). Second, these multistage models show that clonal expansions can have a large impact on cancer incidence. For example, doubling clone sizes is expected to have a similar impact as doubling mutation rates in increasing cancer risk (i.e. in increasing the probability of a cell emerging with the full complement of somatic changes needed to form a tumour). In fact, under an exponential clonal expansion model, a small increase in the growth rate per year can cause a much larger increase in risk than a doubling of somatic mutation rates. Yet, whereas much effort has been devoted to understanding the many mechanisms behind hypermutation in cancer, much less is known about the forces that govern and constrain clonal growth in normal tissues. Finally, despite their simplicity, the models above illustrate how the mode of clonal growth has a large influence on the interpretation of epidemiological data, and how changes in both mutation and selection can alter cancer risk.

The models described above can help understand how cancer risk may be expected to vary with changes in mutation rates, in clone sizes, or in the type of clonal growth. However, they lack several important features that will be needed to build more accurate quantitative models of carcinogenesis as we improve our understanding of the mutation rates and clonal dynamics of normal tissues and premalignant lesions. Some of these are listed below:

1. Clonal competition. As clones grow and come to occupy significant fractions of the available space in the tissue, they can enter into competition. Clonal competition has important implications for cancer risk. First, if the fraction of cells carrying driver mutations in a tissue is high enough, clonal competition may lead to a slowdown of clonal expansions with age. This might contribute to the common but poorly understood slowdown of cancer incidence with age in old age, compared to the rapid increase predicted by multistage models without clonal competition [58]. Second, there is mounting evidence from studies of normal tissues that not all mutations driving clonal expansions are equally carcinogenic, which can interfere with the emergence of cancer. For example, *NOTCH1* mutations in squamous epithelia of the skin, oesophagus and mouth are a potent driver of clonal expansions in the normal epithelium but these clones are believed to be largely benign or even cancer-protective, by out-competing more carcinogenic *TP53*-mutant clones [59].
2. Hypermutation. A common step in precancer evolution is the acquisition of hypermutation, for example in the form of DNA repair defects, APOBEC mutagenesis, high rates of chromosome missegregation, etc. This increase in mutation rates is not considered by the simple models above, but could be incorporated into more complex multistage models.
3. Changes in selection: Selection is context dependent and affected by the microenvironment surrounding a clone. Changes in the tissue microenvironment due to environmental exposures [60], injury [61], chronic inflammation, or tissue ageing [62] can alter the selective pressure on clones, accelerating or suppressing clonal expansions.
4. Epistasis. Specific combinations of driver mutations will be more strongly selected than others, leading to different growth models for different combinations of driver mutations. Similarly, there is evidence in pre-leukaemic evolution that acquisition of second or third hits can lead to

much larger fitness gains [63]. And genetically advanced premalignant or non-invasive lesions carrying several driver mutations, such as Barrett's oesophagus or carcinomas in situ of the bladder can grow to centimetres, compared to the typically microscopic clonal expansions observed in histologically normal oesophagus or bladder epithelium [8,21,45].

*Mutagens and selectogens*

Understanding cancer development as a process of somatic evolution can help unify different models of carcinogenesis, including the *somatic mutation theory*, the *initiation-promotion theory* and the *tissue organisation field theory* (see [64]). These models are often presented as different or even opposing models, but they can be understood as emphasising a different aspect of the somatic evolutionary process, namely somatic mutations, non-mutagenic changes to clonal selection, and the role of tissue architecture and the microenvironment in shaping selection, respectively.

The somatic mutation theory has been the dominant paradigm of the last few decades, particularly since the discovery of oncogenes and tumour suppressor genes in the 1970s through to the current era of cancer genome sequencing. Although the theory acknowledges the role of clonal expansions and selection in the development of tumours (see for example Burnet, Cairns or Nowell, [65–68]), it has likely underplayed the role of non-mutagenic influences on clonal selection during carcinogenesis[5].

In the last few years, several discoveries have led to a renewed interest in the initiation-promotion model of carcinogenesis [60,69]. This model originated in the 1940s with animal studies demonstrating that tumours could be induced in animals by the successive application of an initiator (a mutagen) and a promoter (e.g. a non-mutagenic irritant that favours the growth of mutant cells). The renewed interest in this model stems from the need to recognise the importance of non-mutagenic carcinogens in carcinogenesis. However, we argue that the current understanding of carcinogenesis as a somatic evolution process offers a natural and mechanistic way of incorporating the role of promoters in a multi-stage model of carcinogenesis, unifying the somatic mutation theory and the initiation-promotion theory.

Under the paradigm of somatic evolution, most non-mutagenic carcinogens or promoters are expected to act by favouring the expansion of mutant clones, that is, by altering clonal selection [64]. This can happen through a wide variety of mechanisms, such as increasing proliferation, inducing apoptosis of wild-type cells, causing injury and regeneration, altering interactions with the microenvironment, and enabling immune escape. If these processes lead to an increase in the number of cells with cancer-driver mutations, they are expected to increase the risk of cancer, as shown in the multistage models above. In that context, we argue that many (perhaps most) promoters can be referred to as “selectogens”, which we think is a more precise and mechanistic term than “promoter”, just as we currently use “mutagen” instead of “initiator”. We note that the term “selectogen” has already been coined for this purpose [64].

---

[5] Macfarlane Burnet summarised the somatic mutation theory of carcinogenesis in 1959 as follows (Burnet, 1959): "*...cancer represents the development by a clone of cells (or more than one) of the capacity to multiply freely without regard to the normal controls which maintain cell relationships in the body. This state is reached by a series of mutational events, each of which either results in a selective survival advantage or brings the cell to such a state that a further mutation will endow it with an advantage. Some immediate implications of such a view are: (i) that the common forms of cancer will be in cell lineages which are subject to rapid turnover and in which there is scope for the exercise of selective survival advantage; (ii) that anything which abnormally accelerates turnover, such as chronic trauma or inflammation, will increase the likelihood of cancer...*". However, he arguably underplayed the potential role of non-mutagenic carcinogens: "*If the somatic mutation theory of cancer is correct, the words mutagen and carcinogen should be synonymous. Experimentally this is not quite the case, but there is sufficient number of agents with both types of action to allow us to retain the general hypothesis, with the reasonable qualification (...) that the manifestation of malignancy may require conditions beyond simple mutation*".

We think that the term "selectogen" can help reconcile the somatic mutation and the initiation-promotion models of carcinogenesis under the umbrella of somatic evolution. This borrows the important notion of non-mutagenic influences on clonal growth from the initiation-promotion model, while avoiding its classical association with a two-stage model, incompatible with the genomics of most cancers and premalignant lesions. Instead, cancer development can be understood as a multi-stage process of somatic evolution where both increases in mutation rates and changes in selection can increase cancer incidence by increasing the number of cells at risk of transformation.

The concept of mutagenesis and selectogenesis can also be formalised with the help of the multistage models with clonal expansions described above. Mutagens are expected to increase cancer incidence ($I(t)$) by increasing the number of mutations per cell. Selectogens will increase cancer incidence by increasing the number of cells with driver mutations through clonal expansion. Both mutagens and selectogens will therefore increase the likelihood of a single cell acquiring the full complement of driver mutations needed for transformation. These equations also suggest that an exposure that increased the rate of cell division, cell death or regeneration equally on wild-type and driver-mutant cells without increasing mutation rates (mutagenesis) or the fraction of cells with driver mutations (selection), would not be expected to increase cancer incidence. The equations also suggest that other ways of increasing cancer incidence could be an absolute increase in the number of cells (contained within the $k$ parameter) or, more importantly, a reduction in the number of rate limiting steps needed for transformation ($n$). Examples of the latter may be some germline predisposition alleles (e.g. *RB1* mutations in familial retinoblastoma), and potentially certain exposures (e.g. drug-induced immunosuppression could eliminate one barrier to transformation that may have otherwise required an immune-escape driver mutation).

Finally, whereas carcinogens are often classed as either mutagens or promoters, this classification is simplistic as some carcinogens act simultaneously as both. For example, ultraviolet light acts as both a mutagen and a promoter/selectogen on the epidermis, by causing mutations and by favouring the expansion of *TP53*-mutated cells which are more resistant than wild-type cells to UV-associated apoptosis or differentiation [70]. Similarly, several chemotherapies as well as ionising radiation are directly mutagenic while also favouring the expansion of certain mutant clones particularly resistant to cell death [15,71,72]. The ability to separately quantify mutation rates and selection through genomic studies of normal or precancerous tissues exemplified in the current manuscript (see **Fig. 4** and **Supplementary Note 7**) provides a framework to quantify the mutagenic and selectogenic effects of different carcinogens, in humans and in experimental models (in vitro or in vivo).

**Supplementary Note 6. Simple models of clonal growth in blood and oral epithelium**

Several studies have reported that driver-mutant clones in normal blood grow approximately exponentially with age [63,73,74]. However, clonal growth in solid tissues is likely to be spatially constrained and several alternative growth models have been proposed. For example, colonic epithelium is organised into separate crypts, each maintained by a few stem cells. Clonal expansions are often constrained to single crypts, although certain drivers can lead to larger expansions through crypt fission [75]. In flat epithelia, some studies have assumed exponential growth, whereas others have argued that clonal growth may largely happen at the edges of a clone leading to quadratic growth (e.g. [51,76]).

Detailed modelling of clonal dynamics is beyond the scope of this study. However, a brief discussion here is useful for two reasons. First, as described in **Supplementary Note 5**, the mode of clonal growth is important to understand how changes in mutation rates or clonal expansions are expected to impact cancer risk. Second, the simplified models below highlight that the nature of the increase in aggregate driver density with age (i.e. the fraction of cells carrying a driver mutation, summed across clones) when using single-molecule sequencing can provide information about the underlying clonal dynamics, even in the absence of longitudinal samples or accurate clone size distributions. The models below are simplistic but provide a starting point to discuss these concepts.

If clones grow exponentially, the size of a clone that occurred $t$ years ago is given by **equation (6)** (where $r$ is the exponential growth constant):

$$f(t) = e^{rt}$$

If new driver mutations occur constantly throughout life, with a rate $\mu$ per cell per year, the increase with age in the number of cells with driver mutations, aggregated across all clones, can be approximated by **equation (7)** (where $N_0$ represents the starting number of cells in the tissue).

$$N(t) = N_0\, \mu \int_0^{age} f(t)\, dt$$

Since the population size in an adult normal tissue is approximately constant, we can add a correction factor by dividing $N(t)$ (the number of cells with a driver mutation) by the sum of mutant and wild-type cells. The number of wild-type cells can be approximated as: $N_0\, e^{-\mu t} \approx N_0(1 - \mu t) \approx N_0$ (for low mutation rates, as $\mu t$<<1). **Equation (8)** then provides an estimated density (or fraction) of cells with driver mutations in the tissue: $d(t)$. This correction provides a way of modelling clonal competition without modelling local interactions explicitly.

$$d(t) = \frac{N(t)}{N(t) + N_0}$$

For clones growing exponentially, solving the integral in equation (7) yields **equation (9):**

$$N(t) = \frac{N_0 \mu}{r}(e^{rt} - 1)$$

Equations (8) and (9) imply that the sum of many exponentially growing clones occurring with a uniform rate per year throughout life leads to an exponential increase in the aggregated driver density with age in the absence of clonal competition. In the presence of clonal competition ($d(t)$ instead of $N(t)$), a slower than exponential increase in aggregate driver density is expected, but the effect of clonal competition is limited when the frequency of cells carrying driver mutations in the tissue is modest (e.g. <20%, as is the case in the oral epithelium) (see **Extended Data Fig. 7** for the predicted behaviour of $d(t)$ under different growth models). Of note, if the rate of driver mutations per year is low enough and clones grow exponentially, the total fraction of cells with a driver mutation is expected to be dominated by one major clone (typically an old clone with a high growth rate), as it is commonly observed in clonal haematopoiesis (see **Fig. 4a** for *DNMT3A* and *TET2*).

For clones growing quadratically, solving the integral in equation (7) yields **equation (10)**:

$$N(t) = \frac{N_0 \mu r}{3} t^3$$

This shows that even under a quadratic growth model, which accounts for some spatial constraints leading to clonal growth only at the edges of a clone [76], the aggregated driver fraction increases supralinearly with age (approximately as a function of age to the power of 3).

In epithelia lining ducts or tubules that are narrow and non-branching (e.g. testicular tubules), growth at the edge of the clone occurs largely in one dimension (along the axis), leading to an expected linear growth of individual driver clones with age. Integrating this linear function would lead to a quadratic increase in the aggregated driver density with age ($N(t) \sim t^2$). Analogously, clonal expansions in branching systems (e.g. ductal, vascular, or bronchial trees), under a model where positively-selected clones expand only at their edges, might be expected to follow a function of the type: $f(t) \sim t^k$, with $k$ taking an intermediate value between 1 and 3 depending on the fractal dimension of the tree.

Finally, we can consider the case of highly constrained clonal growth, where clones grow to a maximum clone size and do not grow beyond it. This model could apply to clonal expansions constrained to single crypts or glands, or to clones growing under stringent cell-intrinsic or cell-extrinsic constraints. If we model individual clonal expansions as following a logistic growth function, the initial growth of a clone is exponential but slows down to zero as it approaches the clone's maximum size ($L$). This model yields **equations 11 and 12**:

$$f(t) = \frac{L\, e^{rt}}{L + e^{rt} - 1}$$

$$N(t) = \frac{N_0 \mu L}{r} ln(L - 1 + e^{rt}) - \frac{N_0 \mu L}{r} ln(L)$$

Under this model, if clones grow relatively quickly to their maximum clone size, the increase in the aggregated driver density with age is approximately linear. This is easy to understand: as new driver mutations occur linearly with age and clones grow to their maximum sizes, the increase in driver density with age approaches the product of the driver mutation rate per cell and the (average) maximum clone size (**equation 13**) (see **Extended Data Fig. 7** for a comparison of the predicted behaviour under equations 12 and 13):

$$N(t) \approx N_0\, \mu\, L\, t$$

Note that the normalising factor in $d(t)$ ensures that the total population of cells in the tissue remains constant, and so the approximately exponential or cubic increase in aggregate driver density with age under the exponential and quadratic clonal growth models, respectively, are expected to slow down with age in tissues with a high driver density due to clonal competition. This is likely to be a dominant factor in the increase in driver density with age in tissues where the driver density is high, such as oesophagus, but not in tissues where only a minority of cells carry a driver mutation, such as oral epithelium (**Extended Data Fig. 7**).

The models above are highly simplistic but can help interpret the increase in driver density with age observed in different tissues. In oral epithelium, where the estimated fraction of cells with a driver mutation is seemingly <25%, the approximately linear increase in the aggregated driver frequency with age (despite a continuous occurrence of new mutations with age) suggests that clonal expansions in normal oral epithelium are highly constrained. This is further supported by the slow (sublinear) increase with age of the largest clone (or maximum VAF) in a sample (**Fig. 4a**, **Extended Data Fig. 7**).

### Supplementary Note 7. Additional regression models

As described in the **Methods**, to study the effect of different risk factors and other epidemiological variables on mutation rates, signatures, and drivers, we used multivariate linear mixed-effects regression (LMER) models. In this section, we describe several analyses that complement the results shown in the main text.

*Impact of outliers and normalisation*

Some variables in our dataset show a highly skewed distribution, such as the burden of Signature B or the fraction of cells with driver mutations in some genes. To ensure that the associations reported in the main text (**Fig. 4e**) are not driven by outliers, we used two alternative approaches. First, we compared the results in the main text, which used unnormalised values for all predictor and outcome variables, to the regression models using an inverse-normal transformation (INT) of all variables (**Supplementary Code**). Alternatively, we repeated the regression analyses in the main text excluding outliers from each outcome variable. We defined outliers as those values larger than $3 \times IQR + Q_3$ (where IQR is the interquartile range and $Q_3$ is the third quartile for each outcome variable). Both analyses revealed very similar regression results to those reported in the main text (**Extended Data Fig. 9a**). Despite some quantitative differences, most associations remained significant ($q$-value<0.05) with INT normalisation or outlier removal, despite the potential loss of information caused by the non-linear change in scale with INT and by the exclusion of genuinely informative data points.

*Dose-effects for smoking, alcohol and age*

To estimate the dose-effect relationship of different exposures on the acquisition of somatic mutations, we can look at the coefficients of the LMER models. Using the LMER model described in the main text (**Fig. 4e**) and SNV burden as the outcome variable, the estimated coefficients were: ~15.2 SNVs (per cell or diploid genome) per year of life, ~5.38 per pack-year, and ~0.798 per drink-year. At face value, this suggests that one extra year of life causes as many SNVs as ~2.8 pack-years or ~19.1 drink-years.

This result highlights the considerable mutagenic effect of ageing, which seems qualitatively consistent with age being the largest risk factor for oral cancer. However, caution is needed when interpreting these results, for several reasons. First, it is likely that the slopes reported above for pack-years and drink-years are underestimated due to inaccurate self-reporting. Both systematic underreporting and imprecise reporting would tend to underestimate the slopes. Second, given that SBS5 appears responsible for most of the age-related mutations in the buccal swabs, and that SBS5 is often considered to be caused by endogenous mutational processes [1,77], it could be tempting to conclude that endogenous (and potentially unpreventable) sources of mutation dominate the accumulation of mutations in the oral epithelium and possibly cancer risk. While this might be the case, it is also possible that continuous or frequent exposure to common mutagens in the environment contributes to the age-related accumulation of SBS5 mutations and that a larger than expected fraction of cancer risk is due to preventable factors yet to be discovered. Indeed, differences in cancer incidence across countries or regions has been used to conclude that a large percentage of cancers may be preventable [78], and systematic somatic mutation studies could shed additional light on this question and on the underlying mechanisms. Third, the slopes reported here refer to the mutagenic effect of smoking and alcohol on the oral epithelium in the TwinsUK cohort, which is not necessarily representative of other cell types or populations. For example, much larger mutagenic dose-effects are expected for smoking on bronchial epithelium [79] and bladder urothelium [8], or for alcohol on oral epithelium in individuals with certain risk alleles [9].

Finally, we note that the slopes inferred above assume a linear dose-effect for smoking and alcohol. A linear dose-response relationship may be a reasonable approximation and is often assumed in mutagenesis studies, although non-linear effects are known for some mutagens [80]. We also note that effects of mutation rates on cancer risk can be non-linear [81] (**Supplementary Note 5**), that the effects of pack-years and drink-years on cancer risk can vary by duration and intensity of exposure [82], and that the period since exposure cessation may also be relevant [79]; none of these factors are modelled here.

However, to explore the possibility of non-linear dose-effects for smoking and alcohol, we repeated the LMER model shown in the main text binning pack-years and drink-years into intervals (**Extended Data Fig. 9b**) [83]. This analysis suggests that the linearity assumption is not an unreasonable first approximation, but confidence intervals are too large for a more detailed analysis. Larger studies with more detailed exposure history could be conducted with the methods presented in the current study to address these questions.

A discussion on the potential interaction effect of smoking and alcohol in causing Signature B is included below.

*Interaction effects between smoking and alcohol*

Multiple epidemiological studies have found significant synergistic interactions between smoking and alcohol consumption on the risk of multiple cancer types, including oral cancer, oesophageal cancer and head and neck cancers [84,85]. In the absence of interaction terms, the regression model used in the main text found a significant association of the burden of Signature B (SBS16) with both smoking and alcohol consumption independently, with alcohol showing a stronger association. Somatic mutation studies of oesophageal squamous carcinomas [10] and normal oesophagus [9], have reported an association of SBS16 with germline polymorphisms in the *ALDH2* gene (aldehyde dehydrogenase) supporting a mechanistic link between alcohol (and its metabolite acetaldehyde) and SBS16. However, the association between smoking and Signature B in our buccal swab data does not have a known mechanistic basis. This association could be explained by at least three complementary explanations:

- A direct effect of smoking on Signature B, perhaps through acetaldehyde or other mutagens present in tobacco smoke or its metabolites [86].
- An interaction effect between smoking and alcohol, where smoking increases the mutagenic effect of alcohol, consistent with epidemiological studies of cancer risk.
- A possible confounder effect of inaccuracies in our estimates of alcohol intake, which are estimated by extrapolation of recent consumption. Since smoking and alcohol consumption habits are known to be strongly correlated (which is evident in our data: Spearman's rho = 0.20, *P*-value = 0.0038), misreporting of alcohol consumption could lead to a residual correlation of Signature B with smoking.

To explore these possibilities, we ran several analyses. First, we used a LMER model with an interaction term between pack-years and drink-years. To do so, we applied Z-score normalisation of the variables in the model (dependent and independent). This analysis revealed strong independent associations of Signature B with smoking (*P*-value=$1.5\times10^{-5}$) and alcohol consumption (*P*-value=$6\times10^{-14}$) with Signature B, with an almost non-significant effect for the interaction term (*P*-value=0.049, *q*-value=0.136) (**Supplementary Code**). However, we note that this could be affected by the skewness of both variables, leading to a highly skewed interaction term, as well as by possible non-linear interaction effects.

As an alternative analysis, less affected by skewness, extreme outliers and linearity assumptions, we binned smokers into three groups (never smokers: pack-years = 0, light smokers: (0-20], and moderate/heavy smokers: >20) and we tested the interaction of alcohol and this three-level variable on Signature B. This analysis found evidence of a significant interaction effect, where the increase in Signature B per drink-year is higher with an increased amount of smoking. This interaction is stronger when not including pack-years as an additional independent term, but is significant in both cases (*P*-value=$4.8\times10^{-6}$ and *P*-value=0.006, respectively, Likelihood Ratio Tests, **Supplementary Code**).

*Model H0: sig_denovo_sigB ~ AGE + SEX + T2DM + BMI + missingteeth + ipaq_score + cancer + drink_years + (1 | twin)*
*Model H1: sig_denovo_sigB ~ AGE + SEX + T2DM + BMI + missingteeth + ipaq_score + cancer + drink_years:smoking_group + (1 | twin)*

Finally, a limitation of our estimated “drink-years” is that they are extrapolated from self-reported recent consumption (**Methods**). Since most somatic mutations accumulate neutrally in somatic tissues [3], they provide a record of lifelong exposures to mutagens. We thus expect the amount of Signature B to be more closely related to the total lifetime exposure to alcohol than to recent intake, although we note that the burden of smoking-associated mutations in the bronchial epithelium has been shown to go down after smoking cessation [79]. Individuals with a history of previous heavy drinking but low or zero recent alcohol intake could potentially have high rates of Signature B despite low extrapolated “drink-years” based on recent consumption. Given the correlation between alcohol consumption and smoking habits, this could lead to an indirect association of Signature B with smoking. To evaluate this possibility, we took advantage of the availability of self-reported “lifetime consumption” metadata for a minority of donors (only 302 had lifetime consumption information, compared to 1,034 donors with recent consumption data, which was used to calculate “drink-years” as described in **Methods**).

We first repeated the LMER model used in the main text on donors with lifetime consumption information, including both drink-years (extrapolated from recent consumption data) and self-reported lifetime-consumption to explain Signature B. This revealed that “drink-years” estimated from recent consumption is a better predictor of Signature B than the available self-reported lifetime consumption (*P*-values = $7.5\times10^{-10}$ and 0.42 in the joint model, respectively; **Supplementary Code**). This may reflect that self-reported lifetime consumption is less accurate than our extrapolated estimate based on recent consumption, but it is also possible that current consumption is a better predictor of Signature B. To reduce the risk of an apparent association between Signature B and smoking due to individuals with moderate or high lifetime alcohol consumption but zero recent consumption (estimated drink-years = 0), we repeated the binned interaction analysis above restricting it to individuals with both recent and lifetime consumption data, and excluding those with a history of moderate/heavy drinking and zero recent consumption. This analysis is much less powerful as it is restricted to 202 donors with sufficient metadata, but still supports a significant interaction between smoking and alcohol in causing Signature B (likelihood-ratio test *P*-value = 0.014).

Overall, these analyses provide some evidence of an interaction between smoking and alcohol in their association with Signature B, consistent with epidemiological evidence on their synergistic effect on cancer risk. These analyses offer some support to the hypothesis that smoking may increase Signature B by exacerbating the effects of alcohol. However, we cannot rule out the possibility that at least part of the association of smoking history with Signature B is indirect and due to inaccuracy in self-reported alcohol consumption.

*Regression models to detect putative selectogenic associations*

In the absence of changes on selection pressure, the frequency of driver mutations in the cell population is expected to increase linearly with an increase in mutation rates under a broad set of conditions. This is the case if driver mutations do not cause clonal expansions until the full complement of driver mutations is acquired [42], but also if driver mutations lead to exponential, quadratic, or logistic clonal expansions (**Supplementary Note 6**). However, this linearity assumption may be invalid under certain conditions, such as clonal competition or age-specific exposures. For example, in tissues where most cells already have a driver mutation, an increase in mutation rates can lead to a sublinear increase in driver density, as the relative fitness advantage of driver mutations decreases due to clonal competition. Also, a mutagenic exposure early in life may be expected to have a larger impact on the driver density than the same exposure late in life, as the former will have more time for clonal expansion. This is expected to be more relevant for continuous models of clonal growth (e.g. exponential or quadratic) than for constrained models (e.g. logistic) (**Supplementary Note 6**).

While the limitations above are important to consider, they do not appear too relevant in the oral epithelium (as shown in **Extended Data Fig. 7** and **Supplementary Note 6**). Given that the increase in driver density with age in the oral epithelium is approximately linear, with a small intercept, it is reasonable to accept the simplifying assumption that mutagenic exposures should increase the driver density proportionally to the increase in mutation rates. Purely mutagenic carcinogens (mutagens) may thus be expected to increase driver frequencies proportionally to the increase in mutation rates at driver sites, whereas pure selectogenic carcinogens (selectogens or promoters) may be expected to increase driver frequencies without altering mutation rates. Often, however, some carcinogens may be expected to act as both mutagens and selectogens, causing an increase in mutation rates, and an even larger increase in driver frequency. Below, we discuss two alternative ways to separately quantify mutagenic and selectogenic effects on our data.

To disentangle selectogenic and mutagenic effects, we first repeated the multivariate LMER model described in the main text (**Fig. 4e**) normalising the driver density per gene per donor by the mutation burden in the donor. Given that Signature B (SBS16) is biased towards intronic sequences (**Fig. 4g,h**), for this analysis we used the estimated passenger mutation burden on exons. We then incorporated this as a correction in the LMER model in two alternative ways: (1) using the ratio of driver density and mutation burden as a new outcome variable for each driver gene, or (2) including the mutation burden as a covariate in the model described in the main text. The advantage of the latter is that it does not assume a zero intercept for the effect of mutation rates on the driver density. Both analyses yielded similar results (**Extended Data Fig. 9d**). They suggest that most of the increases in driver density with smoking, alcohol and oral health seem to be explained by (i.e. are approximately proportional to) the increase in mutation rates. However, a higher increase in driver frequency than expected from the increase in exonic mutation rates is observed for *NOTCH1* with pack-years, which suggests that smoking may have both mutagenic and selectogenic effects in the oral epithelium. Other weak positive associations, some significant after multiple testing ($q$-value<0.05) and others nominally significant (uncorrected $P$-value<0.05), are observed between smoking or missing teeth and the driver density of several genes, corrected for exonic mutation burden, potentially suggesting modest selectogenic effects of these exposures in addition to their mutagenic effects. However, given the assumptions underlying these models and the weakness of these associations, larger or more focused studies (e.g. deeper sequencing of fewer genes) would be needed to confirm these associations and shed light on their underlying mechanisms. We also note that, in larger or deeper datasets, dN/dS ratios calculated per gene for each donor could also be used as outcome variables to test for selectogenic associations under the assumptions described above (see **Supplementary Code** for example code using dNdScv). However, the number of synonymous mutations per donor in the current study was too low to provide precise enough dN/dS estimates per gene.

As a complementary analysis, dN/dS ratios per gene can be calculated for different groups of donors. As an example, and to validate the putative selectogenic associations found by the burden-corrected regressions above, we calculated dN/dS ratios for *NOTCH1* and *TP53* for never smokers (pack-years = 0, age ≥50), light and moderate smokers (pack-years: 1-40, age ≥50), and heavy smokers (pack-years >40, age ≥50). The results are shown in the main text (**Fig. 4f**), confirming an increase in dN/dS ratios for *NOTCH1* with increased smoking, and a weaker trend for *TP53*, consistent with the results in the burden-corrected LMER models above.

Finally, we note that while the methods described here could help quantify the mutagenic and selectogenic effects of some carcinogens, the effects of some selectogens or promoters may not be easily detectable by studying the clonal landscape of normal tissues, particularly if they specifically act in later stages of carcinogenesis. We also note that some mutagens act by causing DNA breaks and other structural variants, which are not currently detectable by targeted NanoSeq.

*Regression models on extended medication metadata*

In the LMER analyses described in the main text and in the sections above, we included only some of the most important epidemiological variables in the regressions, including age, sex, BMI and the main risk factors of oral cancer for which we had sufficient information. Self-reported information on medication history was also available for a majority of the donors (n = 738), but it is expected to be incomplete and inaccurate as it relies on questionnaires. Nevertheless, for completion, here we describe an extended LMER model with an additional 25 covariates on self-reported medication history. Only medications reportedly taken by 25 or more donors were included in this analysis. Dosage or duration of treatment was not available and so medication history was recorded as binary variables (yes/no), further limiting the utility of this analysis.

Results are shown in **Extended Data Fig. 9c**, with and without INT normalisation. This analysis revealed only a few weak potential correlations between self-reported medication history and mutation rates or driver frequencies. Importantly, these associations need to be interpreted with caution given the limitations of the available medication metadata. These associations are also likely not to be causal, as medication histories are confounded by the diseases treated by these medications, by other comorbidities, and by associated lifestyle factors and exposures. Nevertheless, we include this supplementary analysis to rule out the possibility that medications may have major confounding effects on the results shown in the main text.

Whereas large cross-sectional studies on the impact of medications on the mutational and clonal landscape of normal tissues should be possible, they will be affected by interindividual heterogeneity and by confounding factors such as disease history and lifestyle. We note that more sensitive and better controlled studies on the impact of different medications on the mutation landscape may be possible using longitudinal sampling of non-invasive biopsies before and after an exposure, followed by whole-genome or targeted NanoSeq.

### Supplementary Note 8. Heritability analyses and GWAS

A recent study reanalysing standard bulk sequencing data from blood from 200,453 individuals in UK Biobank detected clonal haematopoiesis mutations in ~5% of them, identifying 11,697 putative driver mutations in 10,924 individuals. This study was able to perform a genome-wide association study, identifying seven genome-wide significant loci associated with clonal haematopoiesis [14]. A subsequent study on 628,388 individuals identified 24 loci conferring predisposition to clonal haematopoiesis [87]. Using these loci, both studies then used Mendelian randomisation analyses to make causal inferences between clonal haematopoiesis and a range of diseases. These and related studies exemplify how large cohort studies can start to be used to identify germline influences on somatic clonal selection.

Although our dataset is much smaller, it has three features that motivated us to attempt to study heritability and germline influences on somatic mutation phenotypes. First, previous studies could not provide information on mutation rates, being only able to detect the presence or absence of an expanded clone. Our study is the first to be able to measure somatic mutation rates in a normal tissue in over 1000 individuals, providing an initial opportunity to start investigating germline influences on somatic mutagenesis. Second, our sequencing strategy has yielded ~1000-times more driver mutations per individual than the studies above (on average ~60 driver mutations per donor compared to ~0.06) providing more information per donor to increase the power of epidemiological and genetic associations. Third, our twin design, including identical (monozygotic, MZ) and non-identical (dizygotic, DZ) twins, provides additional information to study germline influences and quantify heritability of somatic mutation rates and selection.

Given the twin structure of our dataset and the fact that this is the largest dataset of its kind to date, we carried out exploratory heritability analyses and GWAS. We note, however, that these analyses have limited power and are only exploratory. Future studies with much larger sample sizes will be required to better address these questions and identify specific risk alleles associated with different somatic phenotypes. This may open the door to Mendelian randomisation studies to perform causal inference on the role of somatic mutations on a range of human diseases.

For all analyses below, unless described otherwise, we excluded individuals with mean duplex coverage <200dx, individuals with evidence of HPV (**Methods**), and individuals with a self-reported history of chemotherapy. For analyses using genotyping information (GREML and GWAS), we used pre-existing genotyping array data from TwinsUK. We filtered for AF<1%, HWE $p<1\times10^{-10}$, and missingness>5%, and we inferred cohort-wide relatedness with KING [88]. We used the relatedness information to generate a subset of 539 unrelated individuals to perform PCA with GCTA [89]. After projecting the remaining individuals onto the PC1 vs PC2 space using the SNP loadings, 32 individuals were identified as outliers and were removed from other analyses. Overall, applying all the filters above resulted in a set of 590 samples with genotyping information that were used for GREML and GWAS.

#### *Heritability analyses using the twin design*

We applied three complementary approaches to study heritability using twin information: (1) a comparison of the similarity in somatic variables between MZ and DZ twins using a residual analysis (**Fig. 4i**), (2) ACE models, and (3) genomic-relatedness restricted maximum likelihood (GREML) tests. To avoid testing too many somatic variables with limited statistical power, we restricted these analyses to four particularly informative variables in the buccal swab dataset: SigA mutation burden, SigB mutation burden, *NOTCH1* driver density, and *TP53* driver density.

##### *Paired analysis of residuals*

As an initial attempt to evaluate germline influences on these variables, we first quantified the similarity of somatic mutation rates and somatic driver densities in MZ twins, DZ twins and unrelated same-age pairs of individuals (randomly chosen among individuals of the same age in the cohort). We restricted this analysis to same-sex pairs to avoid a confounding effect for different sex in DZ and unrelated pairs.

We also restricted the analysis to samples with mean duplex depth ≥50dx and those without evidence of HPV.

Twins tend to have more similar exposures and lifestyles than unrelated individuals. To account for the effect of major known confounders, we regressed out the effects of age, sex, pack-years and drink-years using a multivariate linear regression model (see **Supplementary Code**). We then calculated the absolute difference in the residuals for each pair of individuals and used a Wilcoxon test to compare the medians of the resulting distributions for MZ (n=208 pairs used in this analysis), DZ (n=104 pairs) and unrelated individuals (**Fig. 4i**). Applying this approach to donor height as a positive control variable with known strong heritability revealed that the median difference in height was ~1.2cm between two MZ twins, ~4.2cm for DZ pairs (Wilcoxon *P*-value=$4\times10^{-17}$ for MZ vs DZ), and ~5.7cm for unrelated same-age pairs (Wilcoxon *P*-value=0.014 for DZ vs unrelated same-age pairs). Applied to the somatic mutation variables of interest, this analysis showed that MZ pairs are more similar to one another than DZ pairs in the burden of SigA (*P*-value=0.004), and in the density of *NOTCH1* (*P*-value=0.02) and *TP53* (*P*-value=0.016) drivers. No significant differences between MZ and DZ or DZ and unrelated pairs were found for SigB (whose variation in our dataset seems to be dominated by alcohol and smoking).

*GREML*

To formalise the analysis of germline influences on somatic mutation variables, we next estimated SNP heritability ($h^2_g$) based on the subset of 590 individuals who had available genotyping data and passed additional quality controls. We used the variance components model y=g+c+e, where the phenotype (y) is assumed to be a sum of genetic (g), common-environment (c), and residual (e) effects, as suggested by previous studies [90,91]. We used GCTA and 7.66 million SNPs (array and imputed) to build a genetic relatedness matrix (GRM) (for the *g* effect), and a custom binary matrix with 1s indicating twins and 0s otherwise (for the *c* effect), which we gave as input to GCTA-REML. We estimated SNP-h2 for SigA somatic mutation burden, SigB somatic mutation burden, *NOTCH1* density, and *TP53* density, as well as weight and height as positive controls. We worked with both raw values and rank-inverse normal transformed (RINT) outcome values (to increase power and avoid false positives due to outliers). We also incorporated relevant covariates in the model, specifically age, sex, ethnicity, pack-years, drink-years, and the top-10 eigenvectors from PCA. Predictors were standardised (mean = 0, and standard deviation = 1). The estimates are summarised in **Extended Data Table 5**. As a negative control, we repeated this analysis after permuting the phenotypes for all individuals and observed no significant estimates (*P*-value>0.30 for any permutation).

We note that the number of samples used for this analysis (n=590) is smaller than the cohort used for the residuals analyses and the ACE models due to the availability of genotyping information for a subset of donors, and so lack of significant results need to be interpreted with caution. Despite this limitation, this analysis identified evidence of significant heritability for *NOTCH1* (h2g=0.55; se=0.23; *P*=$8.4\times10^{-3}$) and *TP53* (h2g=0.44; se=0.22; *P*=$1.8\times10^{-2}$), as well as for the positive controls height (h2g=0.78; se=0.18; *P* =$3.0\times10^{-11}$) and weight (h2g=0.84; se=0.17; *P*=$1.8\times10^{-4}$). We also found indicative heritability for sigA (h2g=0.42; se=0.26; *P*=0.41), but no indication for sigB (h2g=0). These estimates were obtained using the INT-normalised outcome values, and similar trends were observed when using raw values (see **Extended Data Table 5**).

*ACE*

Finally, as a complementary test, we used ACE models, which is a classical approach for twin studies. ACE models partition the variance of a phenotype of interest (in this case the somatic mutation variables) into three components: additive genetic effects (A), common familial environment (C), and the environmental contribution unique to the individual (E). For a discussion of the underlying assumptions in ACE models see [92]. We relied on the *OpenMx* R package [93] and fitted different models: Saturated, ACE, AE, CE, and E. Likelihood ratio tests were conducted to compare nested models, and we report *P*-values for the comparison between ACE and CE models (**Extended Data Table 5**), i.e.

testing whether the addition of a genetic component provides a significantly better fit. To factor in potentially confounding covariates such as age, sex, pack-years and drink-years, we used regression residuals as input to the models, estimating the heritability of *NOTCH1*, *TP53*, signature A and B burdens, and, as positive controls, weight, height and BMI. Input data was either scaled to aid in the numerical optimization or transformed through RINT (**Extended Data Table 5**). With RINT, which we found to be more robust to outliers, we found significant heritability for *NOTCH1* (h2=0.53, CI95% 0.40-0.63; *P*-value=$1.2\times10^{-3}$), *TP53* (h2=0.49, CI95% 0.15-0.60; *P*-value=$7.2\times10^{-3}$), BMI (h2=0.78, CI95% 0.71-0.83; *P*-value=$5.12\times10^{-9}$), height (h2=0.76, CI95% 0.49-0.93; *P*-value=$6.31\times10^{-16}$), and weight (h2=0.80, CI95% 0.55-0.85; *P*-value=$1.18\times10^{-9}$). Contrary to the direct comparison of residuals, we found no significant heritability for signature A and B burdens. Heritability estimates for the three positive controls (BMI, height and weight) are in line with studies done on larger twin cohorts (e.g.[94])

#### *GWAS*

Given that we detected significant heritability for several somatic mutation phenotypes, we next performed a genome-wide association study (GWAS). To avoid confounding genetic effects with shared environmental effects given the twin relationships in our dataset, we used fastGWA [Jiang et al 2019 NG, [95]], a linear mixed model based on a sparse GRM to account for sample structure. We tested for association between the same set of phenotypes as in the h2 analysis and 7.66 million variants available, using the same samples and covariates, this time further conditioning on the type of zygosity. Using the standard threshold for genome-wide significance of $5\times10^{-8}$ for significance, we detected two associations, both with SigB: an intron variant in *FARP1* (rs145095522; *P*=2.92x10-8; MAF=14.7%), and another intron variant near *GPC6* (*P*=3.76x10-8; MAF=1.2%). It is unclear whether these associations are meaningful, particularly as they are only borderline significant and would not reach genome-wide significance after Bonferroni adjustment of the *P*-value cutoff for the 4 GWAS tests performed ($P>1.25\times10^{-8}$). The two significant SNPs are also single outlier SNPs in their respective loci. For completion, we report the list of loci identified with $P<5\text{x}10^{-6}$, after clumping with PLINK, in **Extended Data Table 5**.

Overall, the lack of clear significant associations is probably unsurprising given the limited sample size. Larger studies will be needed to address these questions in the future.
